# Exploring polygenic-environment and residual-environment interactions for depressive symptoms within the UK Biobank

**DOI:** 10.1101/2021.08.02.21261499

**Authors:** Alexandra C. Gillett, Bradley S. Jermy, S. Hong Lee, Oliver Pain, David M. Howard, Saskia P. Hagenaars, Ken B. Hanscombe, Jonathan R. I. Coleman, Cathryn M. Lewis

**Affiliations:** Social, Genetic and Developmental Psychiatry Centre, Institute of Psychiatry, Psychology and Neuroscience, King’s College London, London, United Kingdom; NIHR Maudsley Biomedical Research Centre, South London and Maudsley NHS Trust, London, SE5 8AF, UK; Australian Centre for Precision Health, UniSA Allied Health and Human Performance, University of South Australia, Adelaide, SA 5000, Australia; Division of Psychiatry, University of Edinburgh, Royal Edinburgh Hospital, Edinburgh, UK; Department of Medical and Molecular Genetics, Faculty of Life Sciences and Medicine, King’s College London

**Author notes:** **Corresponding author:** Dr. Alexandra Gillett, Social, Genetic and Developmental Psychiatry Centre, Institute of Psychiatry, Psychology and Neuroscience - PO80, De Crespigny Park, Denmark Hill, London, United Kingdom, SE5 8AF.

**Keywords:** genotype-environment interaction, residual-environment interaction, depressive symptoms, multivariate reaction norm model

## Abstract

Substantial advances have been made in identifying genetic contributions to depression, but little is known about how the effect of genes can be modulated by the environment, creating a gene-environment interaction. Using multivariate reaction norm models (MRNMs) within the UK Biobank (N=61294-91644), we investigate whether the polygenic and residual variation of depressive symptoms are modulated by 25 a-priori selected covariate traits: 12 environmental variables, 5 biomarkers and polygenic risk scores for 8 mental health disorders. MRNMs provide unbiased polygenic-covariate interaction estimates for a quantitative trait by controlling for outcome-covariate correlations and residual-covariate interactions. Of the 25 selected covariates, 11 significantly modulate depressive symptoms, but no single interaction explains a large proportion of phenotypic variation. Results are dominated by residual-covariate interactions, suggesting that covariate traits (including neuroticism, childhood trauma and BMI) typically interact with unmodelled variables, rather than a genome-wide polygenic score, to influence depressive symptoms. Only average sleep duration has a polygenic-covariate interaction explaining a demonstrably non-zero proportion of the variability in depressive symptoms. This effect is small, accounting for only 1.22% (95% CI [0.54,1.89]) of variation. The presence of an interaction highlights a specific focus for intervention, but the negative results here indicate a limited contribution from polygenic-environment interactions.

## Introduction

Major depressive disorder (MDD) is a common and debilitating mental disorder which is the second leading cause of years lived with disability worldwide (Vos, et al., 2020). It has a lifetime prevalence of 17.8% in global populations (Vos, et al., 2016). The core symptoms of depression are persistent low mood and anhedonia, with other diagnostic signs and symptoms including changes in cognition, appetite, or sleep, and feelings of fatigue and worthlessness. The heritability of MDD is lower than many other psychiatric disorders, estimated at between 30-40%, with higher values for severe cases (Kendall, et al., 2021). This lower heritability suggests that a substantial proportion of liability to depression is due to environmental risk factors.

Genome-wide association studies (GWAS) have made progress in identifying variants associated with MDD, with 178 loci now identified and SNP-heritability estimates ranging between 5.5% and 11.2%, depending on the depression definition used (Levey, et al., 2021). The difference between pedigree and SNP-heritability estimates may indicate a role for additional sources of genetic related variation, such as gene-environment (G-E) interactions. Identifying G-E interactions would provide insight into the biological mechanisms of depression, improve the accuracy of heritability estimates and path the way to individualised preventative healthcare (Hunter, 2005). Genetic studies have used a wide range of definitions of MDD, from diagnosis in clinical studies, to self-report of a diagnosis with depression, to reported presence of depressive symptoms. These criteria show a strong common genetic overlap, with pairwise genetic correlations of at least 0.7 for most MDD definitions (Jermy, et al., 2020; Levey, et al., 2021). We have previously shown that a continuous measure of depression, based on factor analysis of questionnaire responses in the general population effectively captures the polygenetic component of depression (Jermy, et al., 2020).

As noted above, a substantial component of the liability to depression arises from nongenetic factors. Stressful life events and exposure to trauma provide the strongest risks, with smoking, obesity, BMI and exercise also associated with depression (Luppino, et al., 2010; Gianfredi, et al., 2020; Coleman, et al., 2020). The correlation or interaction between these risk factors and the genetic predisposition to depression has been largely un-investigated. Performing studies to disentangle the genetic and environmental contributions to complex traits such as depression is challenging. The environmental variables potentially have a genetic component, and these traits may also be genetically correlated with depression (Wray, et al., 2018). Causation might be multi-directional, with depression risk increased as a consequence of a risk factor, or a risk factor being observed because a healthy lifestyle is more challenging to maintain during a depressive episode. Further, depression may influence the reporting of risk factor status, for example, retrospective reporting of trauma differs from prospective reporting (Baldwin, Reuben, Newbury, & Danese, 2019). These complexities make testing for G-E interactions and correlations challenging, as highlighted by studies investigating an interaction between polygenic risk scores (PRSs) for depression and reported childhood trauma. Early investigations identified an interaction (Peyrot, et al., 2014; Mullins, et al., 2016), but a larger study in the Psychiatric Genomics Consortium (PGC) found no evidence for departure from additive contribution to risk of depression (Peyrot, et al., 2018).

In this paper we model a continuous measure of depressive symptoms and explore genome-wide genotype-covariate (G-C) and residual-covariate (R-C) interactions for 25 covariate traits, including environmental risk factors. A significant G-C interaction means that the additive genetic component for symptoms of depression (G), which has been estimated internal to the data, varies with respect to a covariate trait (C) (Xuan, et al., 2020). This can be thought of as a polygenic-covariate interaction (Dahl, et al., 2020). A significant R-C interaction means that the variation observed in symptoms of depression is modulated by the covariate trait, but in a manner not specified by the model; hence it is a residual interaction. We analyse 17 measured traits in UK Biobank (UKB) including BMI and related body composition traits, exercise measures, smoking, neuroticism, sleep duration, childhood trauma, Townsend deprivation index (TDI) and biomarkers. We additionally analyse PRSs for eight psychiatric disorders. The interactions are modelled in a reaction norm analysis using mtg2 software (Lee & van der Werf, 2016) which tests whether individual differences in the genetic and residual effects are modulated by another risk factor. The multivariate reaction norm model (MRNM) uses covariance functions to model interactions between high-dimensional sets of genetic variants and environmental covariates whilst controlling for trait correlations. It is useful when it is not feasible to investigate interactions variant by variant due to dimensionality (Jarquin, et al., 2014), and has higher power compared to single variant interaction tests for polygenic traits (Dahl, et al., 2020). This statistical framework allows us to robustly investigate the role of these factors in modulating the polygenic, and residual, effects on depression symptoms.

## Methods

### UK Biobank (UKB)

Analysis was performed using the UKB, a health study of over 500,000 UK participants who were recruited in mid-life (40-69 years old) between 2006 and 2010 (Sudlow, et al., 2015). Detailed information on health and lifestyle are available from self-report at baseline, when biological samples for genetic analysis and biomarker testing were also taken. A follow up Mental Health Questionnaire (MHQ), completed online by 157,339 participants in 2016, collected information on a wide range of lifetime psychiatric diagnoses and current depression symptoms (Davis, et al., 2020).

#### Outcome of interest

An outcome trait measuring depression symptoms was derived from the MHQ assessment of depressive symptoms over the last two weeks, which are drawn from the PHQ-9 and correspond to the Diagnostic and Statistical Manual of Mental Disorders criteria for MDD. This trait, which we call depSympt, was constructed via a hierarchical model by Jermy et al. (2020) using MHQ depression-related symptom data. It summarises symptoms related to mood, anxiety, subjective well-being, psychomotor cognitive factors and neuro-vegetative factors. A summary of depSympt and its construction can be found in the Supplementary Materials (SM) section 1.1, with full information given in Jermy et al. (2020).

depSympt is strongly associated with lifetime MDD status, defined using the Composite International Diagnostic Interview Short Form, where it explains 11% of the variation in liability to MDD (Jermy, et al., 2020). On average, prevalent MDD cases have higher depSympt values compared to controls, showing that ever having had depression is associated with increased current depressive symptoms compared to never having had depression (Supplementary Figure 2). Permutation tests (used as depSympt is non-normal) showed highly significant differences between both mean and median depSympt values in MDD cases and controls (*p* < 1*E* − 315).

#### The covariate traits

25 covariate traits were selected, based on previous associations with MDD phenotypes and availability in UKB. These comprise 17 environmental and biomarker covariate traits. Body composition was represented by body mass index (BMI), waist-to-hip ratio, and waist circumference. Exercise variables used the metabolic equivalent task (MET) scores based on the International Physical Activity Questionnaire, which assesses the frequency, intensity and duration of exercise in three categories: walking, moderate exercise and vigorous exercise. Four variables of summed MET minutes per week were analysed: all activities (MET total), and the separate categories of walking (MET walk), moderate exercise (MET mod), and vigorous exercise (MET vig). Other covariate traits from the baseline assessment were Townsend deprivation index (TDI), average sleep duration (sleep), neuroticism score, and pack years of smoking (smoking). Five biomarkers of LDL cholesterol, HDL cholesterol, triglycerides, C-reactive protein (CRP) and vitamin D were analysed. All biomarkers except for LDL were log transformed (see SM section 1.3). From the MHQ, a continuous variable summarising reported childhood trauma was analysed (Pitharouli, et al., 2021). All covariates were from the baseline UKB assessment, except reported childhood trauma, which was collected in the MHQ.

PRSs for the following eight mental health traits were also selected as covariates: major depressive disorder (MDD) (Wray, et al., 2018), bipolar disorder (BIP) (Sklar, et al., 2011), schizophrenia (Ripke, et al., 2014), anxiety (Otowa, et al., 2016), attention deficit hyperactivity disorder (ADHD) (Demontis, et al., 2019), autism spectrum disorder (ASD) (Anney, et al., 2017), obsessive compulsive disorder (OCD) (Arnold, et al., 2018) and anorexia nervosa (Watson, et al., 2019). Construction of the scores is described in SM section 1.2, and details of the GWAS summary statistics are given in Supplementary Table 5.

For the 17 environmental traits, the G-C and the R-C interactions are gene-environment and residual-environment interactions respectively. For the eight PRSs, the G-C interactions represent a narrow form of gene-gene interaction which captures variation in depressive symptoms attributable to an interaction between the PRS and the residual additive genetic component of depression symptoms after removing the fixed effect of the PRS.

#### Genetic data

Autosomal genotype data underwent a centralised quality control procedure described by Bycroft et al. (2018) prior to imputation. We then selected HapMap3 single nucleotide polymorphisms (SNPs) from the imputed UKB genetic data (Ni, et al., 2019; Xuan, et al., 2020), and further removed variants with a minor allele frequency < 0.01, an information score (used to index the quality of genotype imputation) < 0.7 and completeness < 95% (Coleman, et al., 2016).

Quality control for participants followed procedures detailed by (Coleman, et al., 2020). Briefly, analysis was limited to unrelated individuals of European ancestry who had completed the online MHQ, had a call rate of >98% for genotyped SNPs and for whom genetic sex matched self-reported sex. Additionally, individuals were removed for unusual levels of missingness or heterozygosity where recommended by the UKB core analysis team, or if they had withdrawn consent for analysis. After quality control, 126,522 participants were retained. This reduced to 119,692 after omitting individuals missing the outcome trait, depSympt (Jermy, et al., 2020). From the 1,118,287 SNPs retained, genetic relationship matrices (GRMs) were created using Plink version 1.9 for use in the interaction models (Yang, Lee, Goddard, & Visscher, 2011; Chang, et al., 2015).

### Statistical Analysis

Interaction analysis was performed using a mixed effects model called the multivariate reaction norm model (MRNM) (Ni, et al., 2019), which includes two interaction types (polygenic and residual) as random effects, and adjusts for genetic and residual correlations between the outcome and covariate traits. In the sections that follow we provide a broad overview of the MRNM and detail its application in this work, with Figure 1 summarising our approach within the UKB.

**Figure 1.**
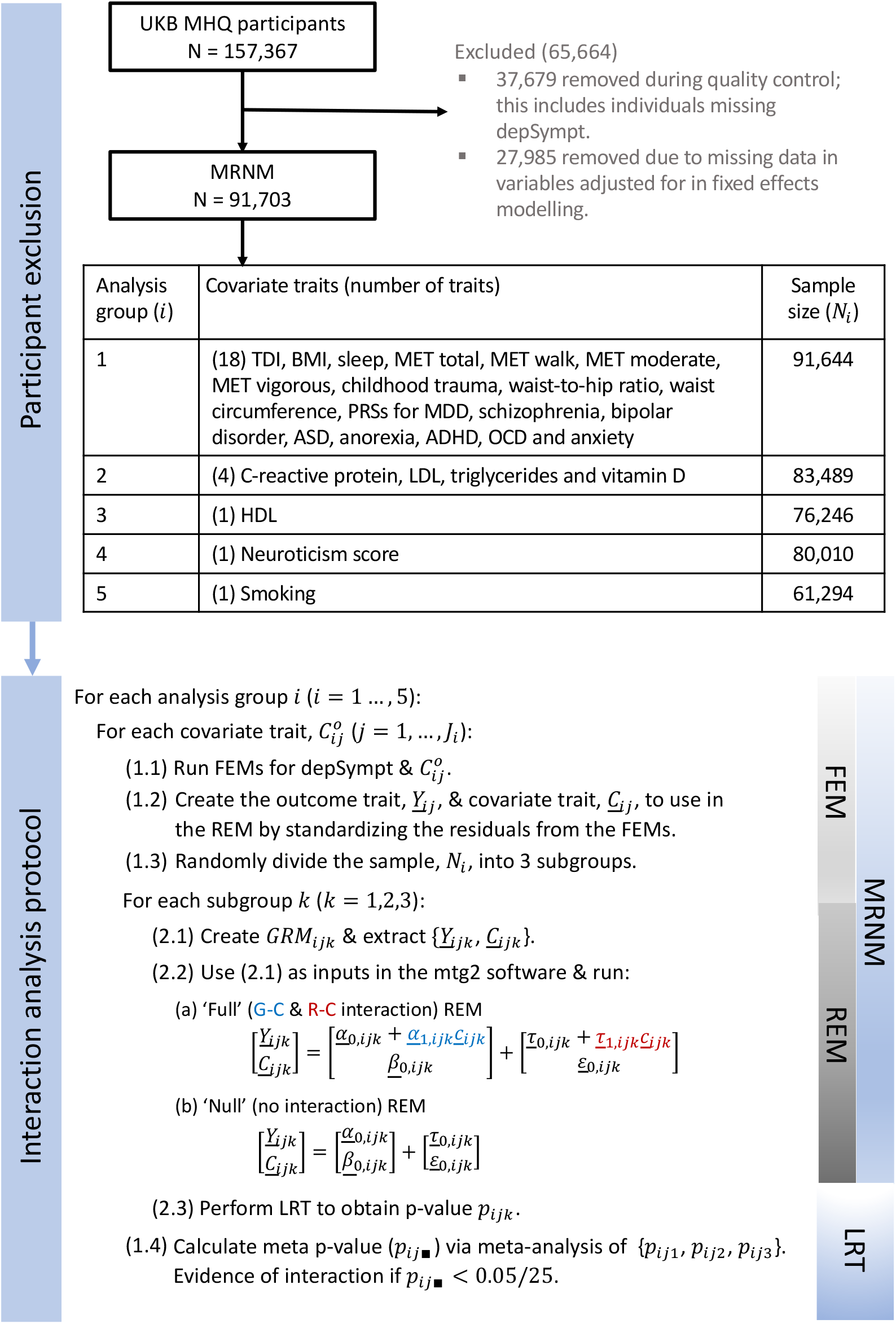
Methods flowchart for UK Biobank: participant exclusion, analysis groupings and scheme for interaction analysis. MRNM = multivariate reaction norm model. FEM = fixed effects model. REM = random effects model. G-C = genotype-covariate. R-C = residual-covariate. LRT = likelihood ratio test. N_ik_ = sample size for analysis group i, subgroup k. <u>C</u>_i1_ (<u>C</u>_i1k_) = vector of length N_i_ (N_i1k_) containing the j^th^ standardised residual covariate trait from analysis group i (and subgroup k). <u>Y</u>_i1_ (<u>Y</u>_i1k_) = vector of length N_i_ (N_ik_) containing the standardised residual outcome trait for interaction analysis with <u>C</u>_i1_ (<u>C</u>_i1k_). <u>α</u>_0,i1k_ (<u>β</u>_0,i1k_) = random effects vector of length N_ik_ representing the contribution to the outcome (covariate) trait for each individual from the homogeneous polygenic component. <u>τ</u>_0,i1k_ (<u>ε</u>_0,i1k_)) = random effects vector of length N_ik_ representing the contribution to the outcome (covariate) trait for each individual from the homogeneous residual component). <u>α</u>_(,i1k_ (<u>τ</u>_(,i1k_) = random effects vector of length N_ik_ representing the contribution to the outcome trait for each individual due to G-C (R-C) interaction.

#### Model overview

A reaction norm (RN) is a genotype-specific function describing the relationship between an outcome and a covariate trait. Interactions are indicated by non-parallel RNs which produce a relationship between the variability of outcome and the covariate within a population. The MRNM looks for evidence of interactions by estimating the trend in outcome variability across the covariate trait, and decomposing this into polygenic and residual sources using estimated genetic similarities from genome-wide SNP data within a random effects model (Schaeffer, 2004; Jarquin, et al., 2014; Ni, et al., 2019). Genetic similarity here is defined using the GRM.

The MRNM has two modelling stages. Firstly, in the fixed effects model, linear regression is used to estimate the expected value of a trait using a set of variables (fixed effects) selected for inclusion (e.g. genotype batch to adjust for possible confounding). This is done for both the outcome trait (here, depSympt) and the covariate traits (see SM section 1.3 for details). Secondly, using a multivariate random effects model, the standardised residual variation in outcome (*Y*) and the covariate trait (*C*), not explained by their respective fixed effects models, is partitioned into genetic and residual random components. For the outcome trait these genetic and residual random effects can be a function of *C*, allowing heterogeneity of the polygenic and residual variance components for *Y* across *C*, thereby incorporating genotype-covariate (G-C) and residual-covariate (R-C) interactions into the model. This random effects model can be written as:

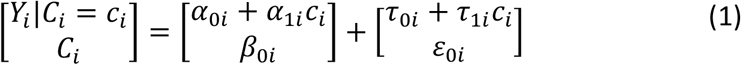

where, for an individual *i*: 1. *α*_0*i*_ (*β*_0*i*_) is a random effect defining the random polygenic intercept for *Y*_*i*_ (*C*_*i*_), 2. *τ*_0*i*_ (*ε*_0*i*_) is a random effect defining the random residual intercept for w*Y*_*i*_ (*C*_*i*_), 3. *α*_1*i*_ is a random effect capturing the interaction of the polygenic component for *Y*_*i*_ with *C*_*i*_ (the G-C interaction), and 4. *τ*_1*i*_ is a random effect capturing the trend in the phenotypic variability of *Y*_*i*_ across *C*_*i*_ that is not explained by the measured genetic variables (the R-C interaction, which can be thought of as covariate-specific noise). These random effects are random variables, characterised by a population multivariate normal distribution with mean zero, and a covariance matrix requiring estimation. The MRNM is therefore parameterised by estimating the covariance matrix between the random effects, which represent sources of (co)variation for *Y* and *C* within the population. Genetic (residual) correlation between the two traits is included in the model by estimating the covariance parameters between the genetic (residual) random effects for *Y* and for *C*. This is an advantage of the MRNM because not accounting for trait correlations can lead to an inflation in the strength of the interactions (Ni, et al., 2019).

Using the MRNM in Equation (1), the marginalised variance for *Y*_*i*_ is:

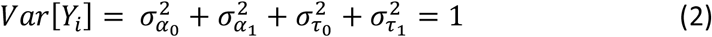

for all individuals in the population. 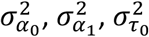 and 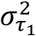 therefore measure the importance of the effect they capture (homogeneous polygenic component, G-C interaction, homogeneous residual component and R-C interaction) in explaining the variability of outcome in the population.

#### Phenotype adjustment

Interactions are assessed for each covariate trait in turn. In the first modelling stage of the MRNM, we adjust depSympt and the covariate trait for fixed effects, using a linear model. Fixed effects included demographic variables (age, sex, and assessment centre) and population structure using the first 15 principal components. We also adjust for stressful and traumatic events in adulthood, due to their likely impact on depressive symptoms. SM section 1.3 details the fixed effects variables used for each depSympt-covariate trait combination.

Except for principal components, continuous fixed effects variables were allowed to have a non-linear relationship with depSympt and the covariate traits by using fractional polynomials (FPs) (Royston & Altman, 1994). We used the mfp package (Benner & Ambler, 2015) within a generalised linear model stats::glm (R Core Team, 2020).

After fixed effects adjustment, all traits are standardised to allow comparison of their relative importance in explaining the variability in outcome across interaction models. For depSympt, we can recover the size of an interaction effect on the original, rather than residualised, scale and use the MRNMs to provide an estimate of the proportion of variability in depSympt explained by G-C and R-C interactions.

Covariate traits were analysed in five groups, each with a different sample size based on the missingness of covariate traits (see Figure 1). This addressed the trade-off between maximising sample size for each depSympt-covariate trait interaction analysis and the computational burden of constructing GRMs for each analysis. The fixed effects models are built using all available data within an analysis group, with sample sizes ranging from 61,294 to 91,644.

#### Identifying interactions

For each (fixed effects adjusted and standardised) covariate trait, we use MRNMs (Ni, et al., 2019) to evaluate evidence of interactions explaining a non-zero proportion of the variability in depSympt. Following the approach of Xuan, et al. (2020), we run a full MRNM, with both G-C and R-C interactions included as random effects, and a null MRNM with no interactions. A likelihood ratio test (LRT) is then used to compare these models, with a significant LRT Bonferroni-corrected p-value (*p* < 0.05/25 = 0.002) providing evidence that interactions explain a non-zero proportion of outcome variance.

This approach assesses the evidence for an overall interaction effect. Simulation studies have shown there is low power to disentangle G-C and R-C interactions in nested MRNM comparisons (Xuan, et al., 2020), with biased G-C interaction estimates to be expected if un-modelled R-C interactions are present (Dahl, et al., 2020). In contrast, the full MRNM produced unbiased estimates of G-C and R-C variance components (Ni, et al., 2019; Xuan, et al., 2020). For significant covariate traits, we can therefore use variance component estimates from the full model, with 95% confidence intervals, to identify which interaction type, polygenic and/ or residual, explain the variability in depSympt.

MRNMs are computationally demanding. Therefore, to perform interaction analysis for a given covariate trait at biobank scale we randomly divided the available UKB participants into three subgroups. MRNMs were run, using the mtg2 package (Lee & van der Werf, 2016), and compared within each subgroup. Results were then meta-analysed using Fisher’s method (Evangelou & Ioannidis, 2013), as described in SM section 1.5. Fixed effects adjustments and post-modelling analysis, including creating graphics, was performed using R version 4.0.4. A Bonferroni correction was applied to adjust for multiple testing (giving a significance threshold of *p* = 0.05/25 = 0.002). A sensitivity analysis was performed by re-fitting the MRNMs using a rank-based inverse normal transformation (RINT) of depSympt. Applying this transformation can control the type I error rate when the assumption of normality is violated (Ni, et al., 2019) and loss of signal indicates spurious interaction effects in the untransformed model (Xuan, et al., 2020). Simulations have shown that parameter estimates from the full model, without applying the RINT, remain unbiased when the normality assumption is violated (Xuan, et al., 2020). The MRNM without transforming depSympt was therefore used for variance component estimation.

## Results

Prior to interaction analysis, linear regression models for the outcome trait, depSympt, with each covariate trait in turn were run (Supplementary Table 8). With the exception of LDL and the PRS for OCD, all covariate traits considered have a statistically significant main effect (*p* < 0.05/25 = 0.002), providing evidence that expected current symptoms of depression vary with these covariate traits. The effect sizes vary widely, ranging from 0.014% of the variability in depSympt explained by the ASD PRS to 21.55% of the variability in depSympt explained by neuroticism score. When fractional polynomials (FPs) are considered, which allow covariate traits to have a non-linear relationship with the expected value of depSympt, the effect of LDL on expected depSympt also becomes statistically significant, although the proportion of variability in depSympt explained is low (0.04%). The proportion of phenotypic variability in depSympt explained by each covariate trait does not greatly change between the main effects and the FP linear models, with average sleep duration having the largest absolute increase from 0.87% in the main effects model, to 2.24% in the FP model (Supplementary Tables 9 and 10).

Distribution plots of the outcome variable summarising depressive symptoms, depSympt, and each covariate trait (before and after fixed effects adjustment) can be found in Supplementary Figures 3 - 27, with Supplementary Table 7 presenting the distribution characteristics for the unadjusted traits. Prior to fixed effects adjustment, the distribution of standardised depSympt is not normally distributed, with evidence of some positive skew (skewness = 0.47). Some deviations from normality are still present after fixed effects adjustment, with a median skewness of 0.34, however, we note that deviations from normality for depSympt after fixed effects adjustment does not mean that the normality assumption of the MRNM is violated since this applies to the distribution of outcome conditional on the fixed *and* the random effects models. Some covariate traits, for example MET total, are highly skewed even after fixed effects adjustment, however, our primary focus is on the variance components for the outcome trait, for which we performed a sensitivity analysis, re-fitting the MRNMs using the RINT for depSympt to control the type I error rate if the assumption of normality is violated.

For each of the 25 covariate traits considered, MRNMs with and without interactions were run for depSympt, and evidence of G-C and/ or R-C interactions was assessed using LRTs. 11 of the 25 covariate traits had p-values below the Bonferroni-corrected significance level, which are presented in Table 1. These 11 traits remained significant in the RINT-based sensitivity analysis (Supplementary Table 13), used to check significant results were not due to normality violations, providing evidence for an interaction effect between depSympt and the following variables: neuroticism, childhood trauma, average sleep duration, BMI, waist circumference, smoking, waist-to-hip ratio, TDI, summed MET minutes for all activities, for walking and for moderate activities. These results show that, even after adjusting for fixed effects, individual-level differences in these covariate traits contribute to variation in depSympt.

**Table 1.** Percentage of variation in depSympt (†adjusted for age, sex, batch and PCs) attributable to the genotype-covariate (G-C) interaction and the residual-covariate (R-C) interaction for covariate traits with significant interaction effects, showing p-value for comparison of the full model to null model, with significance set at α = 0.05/25 = 0.002.

| Proportion of variability in depSympt<sup>†</sup> attributable to: |  |  |  |  |  |
| --- | --- | --- | --- | --- | --- |
| Covariate | p-value | G-C interaction (%) |  | R-C interaction (%) |  |
|  |  | Estimate | 95% CI | Estimate | 95% CI |
| Neuroticism | 5.06E-139 | -0.15 | [-0.76, 0.46] | 2.58 | [ 1.86, 3.30] |
| Childhood trauma | 2.59E-058 | 0.59 | [-0.14, 1.32] | 2.98 | [ 2.18, 3.77] |
| Sleep | 1.97E-041 | 1.22 | [ 0.54, 1.89] | 2.52 | [ 1.78, 3.27] |
| BMI | 6.36E-018 | -0.23 | [-0.86, 0.41] | 1.39 | [ 0.68, 2.09] |
| Waist circumference | 6.15E-016 | -0.15 | [-0.78, 0.48] | 1.48 | [ 0.78, 2.19] |
| Smoking | 2.49E-010 | 0.47 | [-0.52, 1.46] | 1.57 | [ 0.51, 2.63] |
| Waist-to-hip ratio | 4.49E-009 | -0.33 | [-0.95, 0.29] | 1.03 | [ 0.34, 1.73] |
| MET total | 1.92E-007 | 0.23 | [-0.42, 0.87] | 0.53 | [-0.17, 1.24] |
| MET walk | 1.13E-005 | 0.10 | [-0.55, 0.74] | 1.18 | [ 0.45, 1.92] |
| MET mod | 5.73E-004 | -0.26 | [-0.87, 0.35] | -0.08 | [-0.78, 0.61] |
| TDI | 1.96E-003 | -0.19 | [-0.81, 0.42] | 1.67 | [ 0.97, 2.38] |

To assess if these covariate traits modulate the polygenic component and/ or the residual component of depression symptoms (the G-C and R-C interaction respectively), we use a measure of interaction strength: the proportion of variability in depSympt explained by an interaction effect. This measure is plotted as a %, with 95% CIs, for: (a) the G-C interaction and (b) the R-C interaction in Figure 2. When a 95% CI includes zero, we cannot be certain that the interaction term explains any of the variability in depSympt.

**Figure 2.**
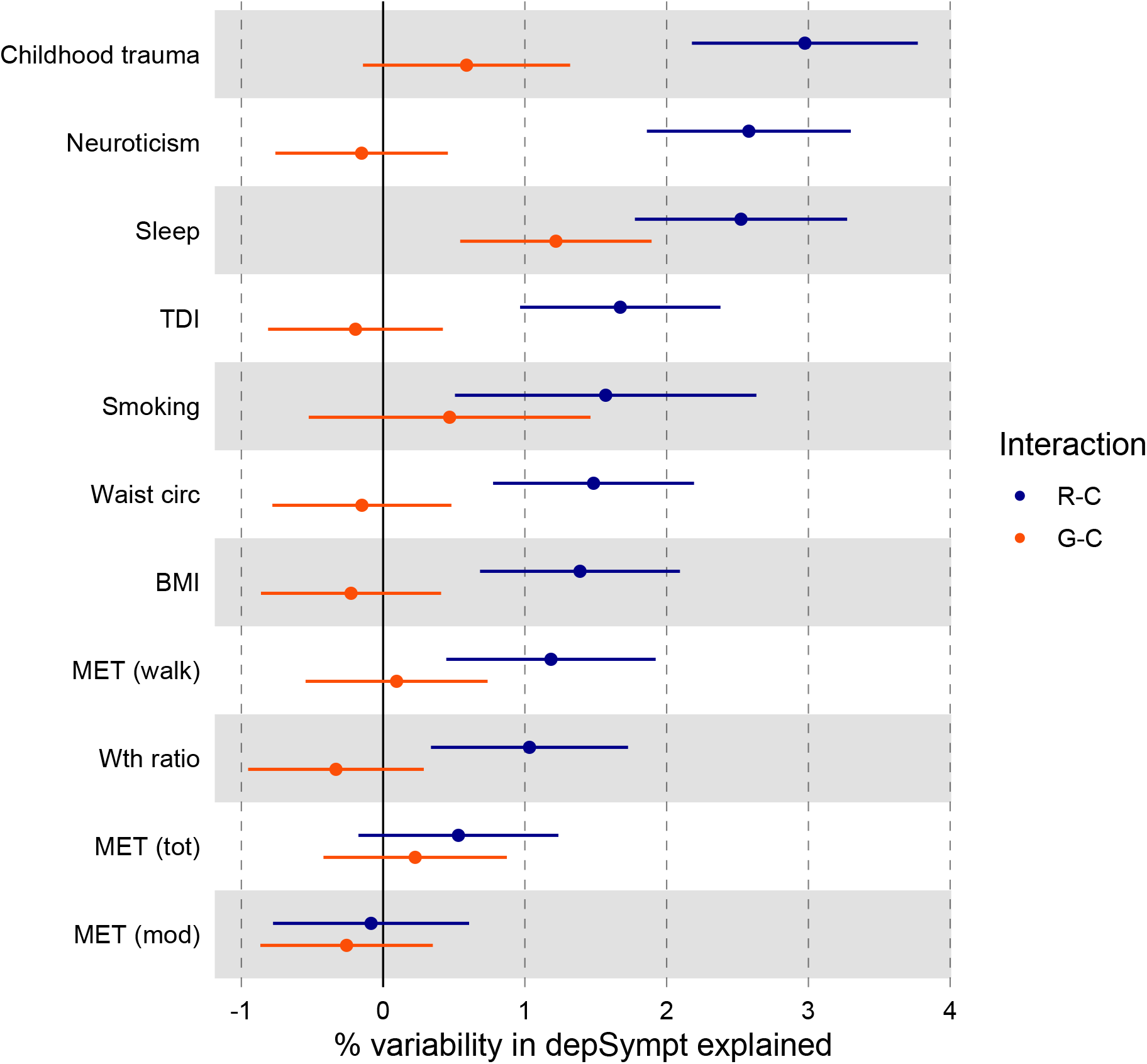
The percentage of variation in depSympt (adjusted for age, sex, batch and PCs) attributable to the R-C (residual-covariate) interaction (blue) and the G-C (genotype-covariate) interaction (red) with 95% confidence intervals.

Nine of the eleven significant covariate traits had 95% CIs for the percentage of variability in depSympt attributable to R-C interactions that excluded zero, with point estimates ranging from 1.03% (95% CI: 0.34-1.73) for waist to hip ratio, to 2.98% (95% CI: 2.18-3.77) for childhood trauma. These results show that a small, but significant, proportion of the residual variability in depSympt is modulated by the following covariate traits: neuroticism, childhood trauma, average sleep duration, BMI, waist circumference, smoking, waist-to-hip ratio, summed MET minutes per week walking and TDI.

For the G-C interactions, only average sleep duration had a 95% CI that excluded zero (Figure 2). This polygenic-sleep interaction is estimated to explain 1.22% of the variability in depSympt (95% CI: 0.54-1.89). Figure 3 plots the relationship between the variance components (polygenic, residual and total) for depSympt and fixed effects adjusted average sleep duration, as estimated by the full MRNM. It shows a U-shaped relationship between the polygenic variance component and sleep, suggesting a larger polygenic contribution to depression symptoms for individuals getting far more or less sleep than expected compared to those in central sleep duration percentiles (see Supplementary Figure 46).

**Figure 3.**
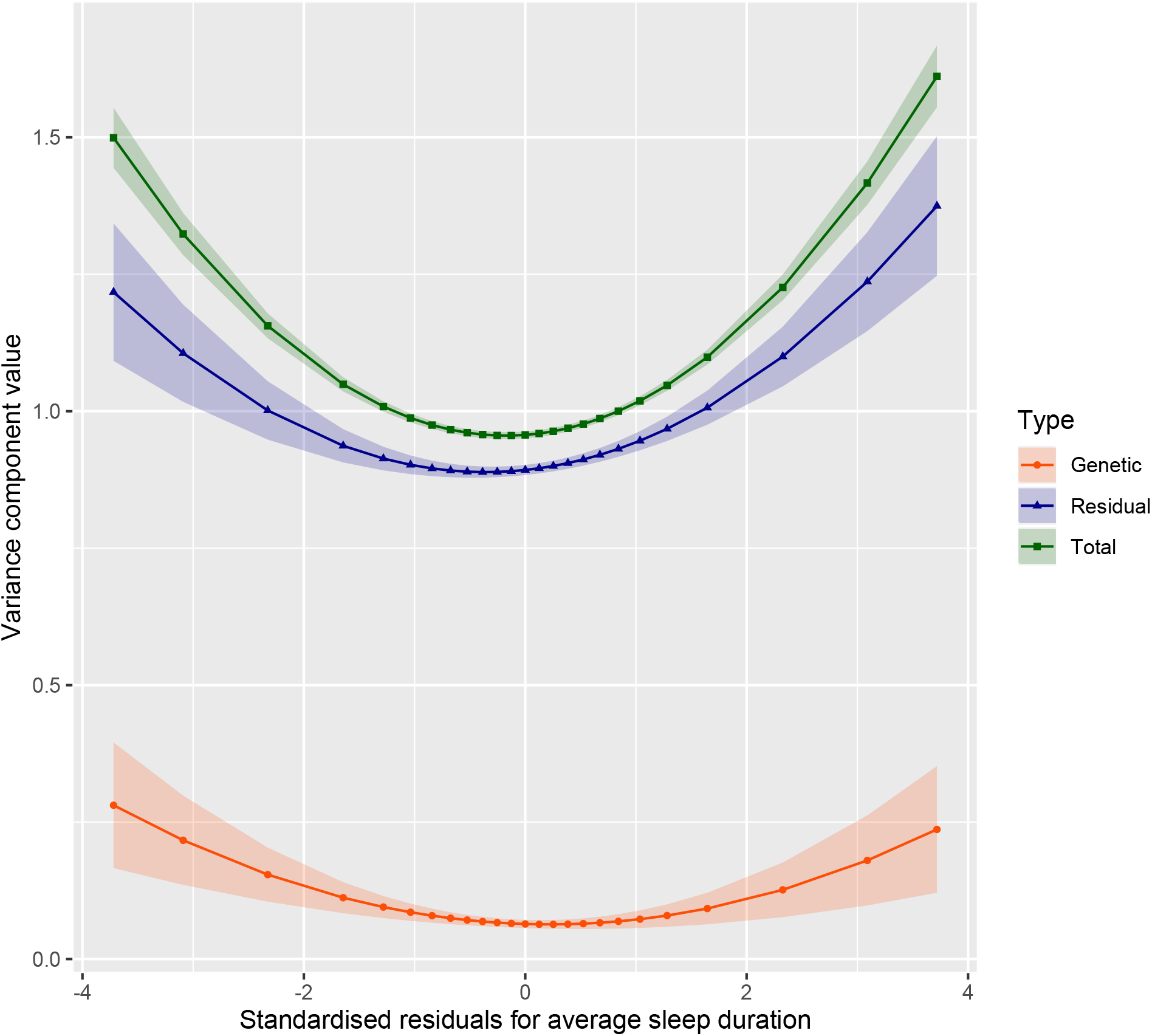
Variance components for standardised residual depSympt by standardised residual average sleep duration, with 95% confidence intervals (presented as coloured bands). See SM section 1.5.3 for variance component and standard error equations.

For the 11 covariates with significant interactions, R-C and G-C interactions account for only a small proportion of the variation in depSympt, and significant results are primarily driven by R-C interactions, not G-C interactions. These results imply that covariate traits exert more influence on depressive symptoms through the modulation of residual (unmodelled) effects compared to polygenic effects.

We note the presence of negative variance component estimates, which are possible within random effects models including the MRNM (Ni, et al., 2019; Xuan, et al., 2020). These values arise as the algorithm estimates this parameter freely, without any constraint that the variance must be positive. All negative variance component estimates have 95% CIs that cross 0, indicating that they are under-estimates of a null interaction effect.

Whilst interactions involving the classic depression environmental risk factors were significant – including with childhood trauma, BMI, exercise measures and smoking – none of the PRSs for psychiatric disorders nor the biomarkers showed significant results for the G-C or the R-C interaction. The MDD PRS was close to the significance threshold (p = 2.63E − 03 > 0.002) and was below the threshold for the RINT-based analysis (p = 7.72E − 04 < 0.002), but the overall impact of both the G-C and R-C interactions, as measured by the proportion of variability in depSympt they explain, have 95% CIs overlapping with zero, in line with the null finding.

For two of the eleven covariate traits, MET total and MET moderate, where the interaction model provided a better fit to the data compared to the null model, the 95% confidence intervals for the proportion of variance explained overlap with 0 for both the R-C and the G-C interactions. These results imply that while there is evidence for a trend in the variability of depSympt across these covariate traits, we are unable to confidently disentangle the source of this interaction into genetic or residual components.

## Discussion

G-E interactions in depression may give insights into its etiology, highlighting both the biological mechanism and identifying environmental risk factors whose role in depression is moderated by genetics. The low heritability for depression compared to many other psychiatric disorders, and the prominent role of environmental risk factors through trauma and stress, make depression a natural target for exploring G-E interactions. Previous G-E interaction studies in depression using genome-wide data, performing SNP-by-environment tests, have identified few significant and replicated results either in European ancestries (Arnau-Soler, et al., 2019) or in Hispanic, African American and Hans Chinese populations (Dunn, et al., 2016; Peterson, et al., 2018). Increasing sample sizes may yield more significant SNP-by-environment interactions, but the highly polygenic nature of depression makes searching for polygenic-environment interactions appealing. Investigations of G-E interactions using depression PRSs have demonstrated null or conflicting results (Kendall, et al., 2021), and these PRS-environment interaction analyses have not modelled a residual trend in the variability of outcome, which can bias G-E interaction estimates (Dahl, et al., 2020). Additionally, the SNP effects used to construct a PRS are estimated assuming an additive genetic model. PRSs are therefore currently designed with the expectation that they do not vary across an environmental gradient. To assess whether the polygenic component for a trait is modified by the environment, a model which can create a polygenic score allowing for this possibility should be employed.

Furthermore, for a complex trait like depression, we would expect the genetic space to map to the phenotypic space through the environment, making the outcome a complex interplay between genes and environment (Assary, Vincent, Machlitt-Northen, Keers, & Pluess, 2020). Since environmental traits are often complex traits themselves, part of this interplay may include a genetic correlation between environment and depression, i.e., BMI, smoking and exercise measures have a positive genetic correlation with depression (Wray, et al., 2018). Not accounting for G-E correlation whilst investigating G-E interaction can lead to biased variance component estimates (Purcell, 2002), and have been shown to inflate the significance of interaction results (Ni, et al., 2019). Interaction analyses using PRSs do adjust for correlation through a main effect, however, they do not allow for residual correlations between traits nor offer the opportunity to investigate G-E interactions in the *presence* of G-E correlation. In this paper we have used MRNMs (Ni, et al., 2019) within the UKB to identify genome-wide genotype-covariate (G-C) and residual-covariate (R-C) interactions for depressive symptoms whilst controlling for residual trait correlations, including genetic correlation. The G-C and R-C interactions allow the polygenic variance component and the residual variance component for depressive symptoms respectively to vary across a continuous covariate trait. The MRNM extends existing polygenic-environment interaction approaches from categorical environmental traits to continuous ones, providing a route to avoid the pitfalls associated with the arbitrary categorising of continuous traits (Altman & Royston, 2006; Naggara, et al., 2011).

We included 25 continuous covariate traits, which covered childhood trauma, body composition, physical activity, smoking and PRSs for eight mental health disorders. For each covariate in turn, MRNMs with and without interactions were run and compared using LRTs. These models jointly test the presence of G-C and/or R-C interaction effects. The contribution of each interaction is then extracted from the variance component estimates, summarised as the proportion of the outcome variability an interaction effect explains.

MRNMs for 11 of the 25 covariates found evidence for some interaction effect. Across these 11 covariates, the variability in depressive symptoms attributable to the R-C interaction effect tended to be substantially larger than that attributable to the G-C effect. Only one covariate, average sleep duration, had a G-C variance component estimate where the confidence interval excluded zero. The significant p-values observed in the LRTs between the null and interaction models are therefore likely to be driven by the R-C interactions. For nine covariate traits, the proportion of variability in depressive symptoms attributable to the R-C interaction had a confidence interval which did not include zero, but the proportion of variance accounted for was small, with the residual-’childhood trauma’ interaction explaining the largest percentage of phenotypic variation at 3.0%, decreasing to 1.0% for the residual-’waist-to-hip ratio’ interaction.

These R-C interactions can be interpreted as non-random noise, where the covariate traits explain additional variation in depSympt not captured explicitly in the model. Several possible extensions to the model may better explain the role of these covariate traits in depressive symptoms. Firstly, these variables may have a non-linear relationship with symptoms of depression not captured by fractional polynomials (FPs); refinement of the non-linear fixed effects model would resolve this e.g. fitting splines. Secondly, the covariates may interact with each other, or with additionally un-modelled environmental variables, to influence depressive symptoms. Finally, the covariates may interact with genetic variants not captured by genome-wide genotyping and imputation (such as rare variants, repeats, or structural variation), or with other omics-type data (such as the transcriptome).

Individually these R-C interactions are small, but cumulatively they could explain a large proportion of the variability in depressive symptoms. Random noise is not useful for prediction and further research to explain the residual heteroscedasticity is warranted. A potential route could be incorporating an environmental similarity matrix into the MRNM and looking for ‘exposome’ effects by utilising shared environmental information (Xuan, et al., 2020).

A further consideration is that the R-C interaction effect can capture deviations from normality in the conditional outcome trait; an effect not necessarily indicative of an interaction, rather driven by the mean-variance relationship of non-normal distributions (Young, Wauthier, & Donnelly, 2018). If this were true, then exploring the significant R-C interactions may not yield useful results. However, possible explanations for the significant R-C interactions, such as environment-environment interactions, have been reported in the depression literature (Hullam, et al., 2019; Morrissey & Kinderman, 2020), indicating that further exploration via multivariate approaches will improve the accuracy of depression models and reveal sets of relevant risk factors unlikely to be identified via univariate methods.

Our primary interest in these models was to assess evidence for G-C interactions for depressive symptoms. Only average sleep duration had an estimate of the proportion of phenotypic variability explained by a G-C interaction with a 95% confidence interval not overlapping with zero. A small, but statistically significant, proportion of the variability in current depressive symptoms is therefore attributable to a genome-wide G-E interaction with average sleep duration measured at the UKB baseline assessment, 5-10 years earlier than the assessment of depSympt. Our results imply that the optimal sleep duration to minimise depressive symptoms can vary by genetic profile, but this modification of the polygenic variation for depSympt using historic sleep patterns is limited, with the estimate of variation attributable to this interaction being low at 1.2%. Other covariate traits that had non-zero estimates for R-C interaction variance component had much lower G-C interaction variance components, ranging from 0.59% (for childhood trauma), to estimates that were below zero (indicating a zero effect).

This is not the first study to identify a significant gene-sleep interaction for depression. A twin study by Watson, et al. (2014) found that the genetic contribution to depressive symptoms was significantly higher for both short (<7 hours per night) and long (≥ 9 hours per night) sleep durations compared to the average (7-8.9 hours per night)-a trend that we also observed (Supplementary Figure 46). Additionally, there is evidence for a complex bidirectional relationship between sleep and depression involving variables/ biomarkers such as circadian rhythms (Kronfeld-Schor & Einat, 2012; Khan, et al., 2018), stress (Leggett, Burgard, & Zivin, 2016; Palagini, et al., 2019), melatonin (Rahman, Marcu, Kayumov, & Shapiro, 2010), serotonin (van Dalfsen & Markus, 2019), dopamine (Finan & Smith, 2013; Boland, Goldschmied, Wakschal, Nusslock, & Gehrman, 2020), and their respective genes. Future work investigating gene-sleep interactions for depression could utilise these previously highlighted genes within genomic partitioning analysis.

An initially surprising result is that this MRNM analysis does not support a non-zero G-C interaction effect for childhood trauma, despite G-C interactions of childhood trauma with depression previously having been identified (Peyrot, et al., 2014; Mullins, et al., 2016; Shen, et al., 2020). In addition to methodological and dataset differences, this study used a recently developed, composite measure of childhood trauma (Pitharouli, et al., 2021). The null G-C interaction result here, in contrast with previous significant results for other measures of childhood trauma, may suggest that G-C interactions for differing types of childhood trauma should be investigated as separate covariates and not as a continuous weighted aggregate.

Although estimated interaction effects for symptoms of depression are small, the MRNM has been able to identify strong lifestyle modulation effects on cardiovascular traits. Xuan et al. (2020) used the MRNM and 22 lifestyle traits to explore interactions for 23 cardiovascular traits, finding evidence of lifestyle modulation for 42% of the outcome-covariate trait pairings. Sizeable G-C and R-C interactions were observed suggesting the need for personalised lifestyle interventions to reduce the risk of cardiovascular disease. The largest G-C interaction explained ∼10% of (fixed effects adjusted) phenotypic variability and was for the modulation of the polygenic effect on HDL cholesterol level by physical activity. The largest R-C interaction effect, explaining ∼20% of phenotypic variability, was for the modulation of the nongenetic component for white blood cell count by smoking.

Our study has limitations. The environmental risk factors analysed here were selected based on a literature review, but no systematic review or meta-analysis was performed and other variables with equally compelling rationale for inclusion may not have been considered. Similarly, PRSs for traits that have a genetic correlation with depression were included, together with biomarkers that have been widely tested for association with depression (CRP, vitamin D) but we cannot exclude interaction with other biomarkers. Additionally, the modifying effect of covariate traits on depressive symptoms were considered in separate models. A joint model for interactions may provide a better fit, however, the computational burden and the required sample size for robust parameter estimation will increase. All statistical models require assumptions about variables to be analysed. We analysed a continuous measure of depression, as required by the MRNM, and chose to use a composite measure previously constructed and validated as highly correlated with MDD diagnosis. Other options would have been to take raw scores for numbers of depression symptoms reported, or the ordinal measure of presence/absence of the two core depression symptoms (each scored 0, 1, 2), as analysed in other genome-wide association studies (Turley, et al., 2018; Levey, et al., 2021). The outcome trait depSympt, extracted from a hierarchical model, was chosen as it accounts for correlation between reported depression symptoms, and is continuous. The UKB depressive symptoms, from which the depSympt variable was constructed, represents a snapshot of participants mental health over the two weeks prior to completing the MHQ. This does not account for historical mental health, or capture trends of mental health, and the potential dynamic nature of G-E interactions, over time. Our study analysed only European ancestries in UKB and used only PRSs from GWASs of European ancestry. The MRNM provides a flexible and broad modelling framework, but model fitting is computationally intensive, particularly given the large sample sizes available in UKB. Meta-analysis across three subgroups was required to make this computational feasible. Ni et al. (2019) showed that using meta-analysis across subgroups reduced the power to identify non-zero interaction variance components compared to using the complete sample, and analysis with the full available UKB sample might yield more significant G-C/ R-C interactions. The MRNM used here estimates the polygenic-covariate interaction and, as an aggregate genome-wide measure, does not provide accessible information about the contribution from individual variants, genes, or pathways (Assary, Vincent, Machlitt-Northen, Keers, & Pluess, 2020), although the model could be extended for use within genomic partitioning.

In summary, the MRNM provides a flexible, if computationally intensive, framework to comprehensively model genetic and environmental contributions to complex traits. For depression, these models show significant R-C interactions, potentially highlighting unmodelled relationships between nongenetic contributions to depressive symptoms. Evidence for a G-C interaction was only found at one covariate (average sleep duration), suggesting that interactions between polygenic scores and the explored environmental risk variables do not play a major role in the etiology of depressive symptoms, and therefore, personalised lifestyle interventions based on SNP profiles are not required.

## Data Availability

Individual-level data for UK Biobank must be applied for via the Access Management System (https://www.ukbiobank.ac.uk/enable-your-research/apply-for-access).

## URLS

- LDSC HapMap 3 SNP-list: https://data.broadinstitute.org/alkesgroup/LDSCORE/w_hm3.snplist.bz2
- mtg2 software: https://sites.google.com/site/honglee0707/mtg2

## Acknowledgements

This paper represents independent research funded by the UK Medical Research Council (MR/N015746/1), and the National Institute for Health Research (NIHR) Biomedical Research Centre at South London and Maudsley NHS Foundation Trust and King’s College London. The authors acknowledge use of the research computing facility at King’s College London, Rosalind (https://rosalind.kcl.ac.uk), which is delivered in partnership with the NIHR Maudsley BRC, and part-funded by capital equipment grants from the Maudsley Charity (award 980) and Guy’s & St. Thomas’ Charity (TR130505). The views expressed are those of the authors and not necessarily those of the NHS, the NIHR or the Department of Health and Social Care. Saskia P. Hagenaars was supported by the Medical Research Council (MR/S0151132). David M. Howard is supported by a Sir Henry Wellcome Postdoctoral Fellowship (Reference 213674/Z/18/Z) and a 2018 NARSAD Young Investigator Grant from the Brain & Behaviour Research Foundation (Ref: 27404).

UKB: This research was conducted under UK Biobank application 18177.

