## Supplementary Materials for "Exploring polygenic-environment and residual-environment interactions for depressive symptoms within the UK Biobank"

27 Histograms of standardised pack-years smoking as a proportion of age . . 67
28 Histograms of standardised CRP and log-CRP . . . . . . . . . . . . . . . 68
29 Histograms of standardised Triglycerides and log-Triglycerides . . . . . . . 68
30 Histograms of standardised Vitamin D and log-Vitamin D . . . . . . . . . 69
31 Histograms of standardised LDL and log-LDL . . . . . . . . . . . . . . . . 69
32 Histograms of standardised HDL and log-HDL . . . . . . . . . . . . . . . 70
33 Forest-plot of the proportion of variation in depSympt attributable to: (a) a genotype-covariate interaction, and (b) a residual-covariate interaction, with 95% confidence intervals, for all 3 subgroups & the meta-analysis. . . 72
34 Forest-plot of the proportion of variation in depSympt attributable to: (a) a genotype-covariate interaction, and (b) a residual-covariate interaction, with 95% confidence intervals, for all 3 subgroups & the meta-analysis. All covariate traits. . . . . . . . . . . . . . . . . . . . . . . . . . . . . 73
35 Variance components for $Y$ (residualised depSympt) against residualised and standardised neuroticism score. . . . . . . . . . . . . . . . . . . . . . 74
36 Variance components for $Y$ (residualised depSympt) against residualised and standardised childhood trauma summary variable. . . . . . . . . . . 75
37 Variance components for $Y$ (residualised depSympt) against residualised and standardised average sleep duration. . . . . . . . . . . . . . . . . . . 76
38 Variance components for $Y$ (residualised depSympt) against residualised and standardised BMI. . . . . . . . . . . . . . . . . . . . . . . . . . . . 77
39 Variance components for $Y$ (residualised depSympt) against residualised and standardised waist circumference. . . . . . . . . . . . . . . . . . . . 78
40 Variance components for $Y$ (residualised depSympt) against residualised and standardised smoking. . . . . . . . . . . . . . . . . . . . . . . . . . . 79

41 Variance components for $Y$ (residualised depSympt) against residualised and standardised waist to hip ratio. . . . . . . . . . . . . . . . . . . . . . . . 80
42 Variance components for $Y$ (residualised depSympt) against residualised and standardised MET total. . . . . . . . . . . . . . . . . . . . . . . . . . 81
43 Variance components for $Y$ (residualised depSympt) against residualised and standardised MET walk. . . . . . . . . . . . . . . . . . . . . . . . . . 82
44 Variance components for $Y$ (residualised depSympt) against residualised and standardised MET moderate. . . . . . . . . . . . . . . . . . . . . . . 83
45 Variance components for $Y$ (residualised depSympt) against residualised and standardised TDI. . . . . . . . . . . . . . . . . . . . . . . . . . . . . 84
46 Proportion of the total variance component for $Y$ (residualised depSympt) against residualised and standardised average sleep duration. . . . . . . . 85
47 Plots of the variance components for normalised depSympt across normalised MET (total) . . . . . . . . . . . . . . . . . . . . . . . . . . . . . . 86
48 Plots of the variance components for normalised depSympt across normalised MET (moderate) . . . . . . . . . . . . . . . . . . . . . . . . . . . . 87

### 1 Supplementary Methods

#### 1.1 depSympt: understanding the outcome trait

depSympt was created by Jermy et al. [2020]. Using depression-related symptom data from the Mental Health Questionnaire (MHQ) within the UK Biobank (UKB), the authors performed a factor analysis to identify latent continuous factors that could be driving the observed symptoms. A hierarchical model with five first-order factors and one second-order factor was identified. Please see Table (1) for the inputted symptom data, and Figure (1) for a visualisation of the identified hierarchical model.

The first-order factors can be labelled by the group of symptoms that they capture: 1. Mood (capturing depressed thoughts, anhedonia and suicidal thoughts), 2. Anxiety (capturing symptoms related to anxiety, nervousness, worry, foreboding and restlessness), 3. Subjective well-being (related to general wellbeing, belief in meaningfulness of own life and suicidal thoughts), 4. Psychomotor Cognitive factor (capturing symptoms for impaired concentration, restlessness and psychomotor retardation or agitation), and, 5. Neuro-vegetative factor (capturing changes in appetite, energy and sleep).

The second order factor, which we call the depSympt, can be thought of as a continuous depression score involved in driving all of the identified first-order factors. As such, it is highly correlated with the five first-order factors, with correlations ranging between 0.73 and 0.96 within the MHQ sample used by Jermy et al. [2020] ($n = 148,957$), and ranging between 0.84 and $\approx 1$ (see Tables (2) and (3)) within the reduced sample with genetic data available used in this work ($n = 119,690$).

Due to these high correlations, we selected the depSympt to be the outcome trait when investigating genotype-covariate (G-C) and residual-covariate (R-C) interactions for a depressive symptom trait. A low depSympt score is associated with having low severity or no depression-related symptoms at the time of taking the MHQ. Conversely, a high depSympt score is associated with having an increased number of symptoms, with an increased severity level. For details please see Table (4), which provides mean depSympt across severity levels for each of the 15 symptoms included in the final factor analysis model.

Table (3) shows that 11.06% of the variability in liability to (prevalent) depression is attributable to depSympt (this is the highest of all of the created latent factors). Figure (2) presents a density plot for depSympt within the available UKB study population, grouped by MDD status, demonstrating that the average depSympt value for MDD cases is larger than that for controls (0.35 compared to -0.18). Therefore, although depSympt is a continuous summary variable capturing current depressive symptoms, it is also associated with being a prevalent MDD case. Interactions identified within this study would warrant investigation using case-control depression phenotypes.

Table 1: The 18 original symptom variables selected from UKB to be used in the factor analysis which created depSympt. Table taken from the Supplementary Materials from Jermy et al. [2020].

| Field | Symptom Class | Symptom | Question |
|---|---|---|---|
| 20510 | Depressive Symptoms | Depressed mood | Over the last 2 weeks, how often have you been bothered by any of the following problems? Feeling down, depressed, or hopeless |
| 20514 | Depressive Symptoms | Anhedonia | Over the last 2 weeks, how often have you been bothered by any of the following problems? Little interest or pleasure in doing things |
| 20511 | Depressive Symptoms | Appetite loss or gain | Over the last 2 weeks, how often have you been bothered by any of the following problems? Poor appetite or overeating |
| 20517 | Depressive Symptoms | Insomnia or hypersomnia | Over the last 2 weeks, how often have you been bothered by any of the following problems? Trouble falling or staying asleep, or sleeping too much |

Continued on next page

**Table 1 – continued from previous page**

| Field | Symptom Class | Symptom | Question |
|---|---|---|---|
| 20518 | Depressive Symptoms | Psychomotor agitation or retardation | Over the last 2 weeks, how often have you been bothered by any of the following problems? Moving or speaking so slowly that other people could have noticed? Or the opposite- being so fidgety or restless that you have been moving around a lot more than usual |
| 20519 | Depressive Symptoms | Fatigue or loss of energy | Over the last 2 weeks, how often have you been bothered by any of the following problems? Feeling tired or having little energy |
| 20507* | Depressive Symptoms | Feelings of inadequacy | Over the last 2 weeks, how often have you been bothered by any of the following problems? Feeling bad about yourself or that you are a failure or have let yourself or your family down |
| 20508 | Depressive Symptoms | Impaired ability to think, concentrate | Over the last 2 weeks, how often have you been bothered by any of the following problems? Trouble concentrating on things, such as reading the newspaper or watching television |
| 20513 | Depressive Symptoms | Recurrent thoughts of death or suicide ideation, plan for committing suicide | Over the last 2 weeks, how often have you been bothered by any of the following problems? Thoughts that you would be better off dead or of hurting yourself in some way |
| 20506 | Anxiety Symptoms | Nervous, anxious or on edge | Over the last 2 weeks, how often have you been bothered by any of the following problems? Feeling nervous, anxious or on edge |
| 20509 | Anxiety Symptoms | Uncontrollable worrying | Over the last 2 weeks, how often have you been bothered by any of the following problems? Not being able to stop or |
| | | | Continued on next page |

**Table 1 – continued from previous page**

| Field | Symptom Class | Symptom | Question |
|---|---|---|---|
| | | | control worrying |
| 20515* | Anxiety Symptoms | Trouble relaxing | Over the last 2 weeks, how often have you been bothered by any of the following problems? Trouble relaxing |
| 20505* | Anxiety Symptoms | Irritable | Over the last 2 weeks, how often have you been bothered by any of the following problems? Becoming easily annoyed or irritable |
| 20520 | Anxiety Symptoms | Worrying about different things | Over the last 2 weeks, how often have you been bothered by any of the following problems? Worrying too much about different things |
| 20512 | Anxiety Symptoms | Foreboding | Over the last 2 weeks, how often have you been bothered by any of the following problems? Feeling afraid as if something awful might happen |
| 20516 | Anxiety | Restlessness | Over the last 2 weeks, how often have you been bothered by any of the following problems? Being so restless that it is hard to sit still |
| 20458 | Happiness and subjective well-being | General Happiness | In general, how happy are you? |
| 20460 | Happiness and subjective well-being | Belief that own life is meaningful | To what extent do you feel your life to be meaningful? |

* 3 symptom variables excluded from the final factor analysis model

Table 4: The relationship between depSympt and the current depressive symptoms variables used in its creation by Jermy et al. [2020].

| Field | Symptom | depSympt mean | depSympt sd | p-value |
|---|---|---|---|---|

Continued on next page

**Table 4 – continued from previous page**

| Field | Symptom | depSympt mean | depSympt sd | p-value |
|---|---|---|---|---|
| 20458 | General happiness | | | < 2.2e-16 |
| | Extremely happy | -1.17 | 0.75 | |
| | Very happy | -0.39 | 0.68 | |
| | Moderately happy | 0.51 | 0.74 | |
| | Moderately unhappy | 1.64 | 0.71 | |
| | Very unhappy | 2.21 | 0.91 | |
| | Extremely unhappy | 2.80 | 0.97 | |
| 20460 | Life feels meaningful | | | < 2.2e-16 |
| | Not at all | 1.33 | 1.31 | |
| | A little | 1.17 | 0.95 | |
| | A moderate amount | 0.52 | 0.85 | |
| | Very much | -0.26 | 0.81 | |
| | An extreme amount | -0.68 | 0.94 | |
| 20506 | Nervousness/ anxiety | | | < 2.2e-16 |
| | Not at all | -0.37 | 0.80 | |
| | Several days | 0.81 | 0.70 | |
| | > 1/2 the days | 1.56 | 0.80 | |
| | Nearly every day | 1.99 | 0.94 | |
| 20508 | Trouble concentrating | | | < 2.2e-16 |
| | Not at all | -0.27 | 0.81 | |
| | Several days | 1.10 | 0.62 | |
| | > 1/2 the days | 1.89 | 0.68 | |
| | Nearly every day | 2.31 | 0.89 | |
| 20509 | Uncontrolled worrying | | | < 2.2e-16 |
| | Not at all | -0.33 | 0.80 | |
| | Several days | 0.93 | 0.67 | |
| | > 1/2 the days | 1.61 | 0.74 | |
| | Nearly every day | 2.03 | 0.89 | |
| 20510 | Feelings of depression | | | < 2.2e-16 |
| | Not at all | -0.38 | 0.71 | |
| | Several days | 1.20 | 0.45 | |
| | > 1/2 the days | 2.14 | 0.43 | |
| | Nearly every day | 2.77 | 0.58 | |
| 20511 | Under or over eating | | | < 2.2e-16 |
| | Not at all | -0.24 | 0.85 | |
| | Several days | 0.89 | 0.72 | |
| | > 1/2 the days | 1.50 | 0.79 | |
| | Nearly every day | 1.91 | 0.96 | |
| 20512 | Feelings of foreboding | | | < 2.2e-16 |
| | Not at all | -0.22 | 0.87 | |

Continued on next page

**Table 4 – continued from previous page**

| Field | Symptom | depSympt mean | depSympt sd | p-value |
|---|---|---|---|---|
| | Several days | 0.95 | 0.74 | |
| | > 1/2 the days | 1.62 | 0.83 | |
| | Nearly every day | 2.04 | 0.95 | |
| 20513 | Suicidal/ self-harming thoughts | | | < 2.2e-16 |
| | Not at all | -0.08 | 0.92 | |
| | Several days | 1.75 | 0.65 | |
| | > 1/2 the days | 2.51 | 0.63 | |
| | Nearly every day | 2.99 | 0.76 | |
| 20514 | Anhedonia | | | < 2.2e-16 |
| | Not at all | -0.32 | 0.75 | |
| | Several days | 1.26 | 0.46 | |
| | > 1/2 the days | 2.04 | 0.52 | |
| | Nearly every day | 2.52 | 0.79 | |
| 20516 | Restlessness | | | < 2.2e-16 |
| | Not at all | -0.15 | 0.91 | |
| | Several days | 1.01 | 0.77 | |
| | > 1/2 the days | 1.70 | 0.91 | |
| | Nearly every day | 1.80 | 1.11 | |
| 20517 | Sleep problems | | | < 2.2e-16 |
| | Not at all | -0.55 | 0.79 | |
| | Several days | 0.37 | 0.74 | |
| | > 1/2 the days | 0.88 | 0.85 | |
| | Nearly every day | 1.21 | 1.02 | |
| 20518 | Movement and/or speaking changes | | | < 2.2e-16 |
| | Not at all | -0.09 | 0.93 | |
| | Several days | 1.44 | 0.72 | |
| | > 1/2 the days | 2.19 | 0.82 | |
| | Nearly every day | 2.27 | 1.12 | |
| 20519 | Fatigue | | | < 2.2e-16 |
| | Not at all | -0.66 | 0.68 | |
| | Several days | 0.46 | 0.66 | |
| | > 1/2 the days | 1.23 | 0.73 | |
| | Nearly every day | 1.66 | 0.92 | |
| 20520 | Changes in worry | | | < 2.2e-16 |
| | Not at all | -0.42 | 0.78 | |
| | Several days | 0.75 | 0.69 | |
| | > 1/2 the days | 1.53 | 0.77 | |
| | Nearly every day | 1.96 | 0.91 | |

sd = standard deviation. P-value is from a likelihood ratio test comparing generalised linear models for depSympt with and without the symptom included.

Continued on next page

**Table 4 – continued from previous page**

| Field | Symptom | depSympt mean | depSympt sd | p-value |
|---|---|---|---|---|
| Symptom variables are current symptoms at the time of taking the MHQ. | | | | |

Table 2: Proportion of variation in liability to depression* explained by six latent depression symptom scores [Jermy et al., 2020], and the correlation of these scores with depSympt. (*depression here is defined using data fields `20446` and `20441`. A case will answer yes to at least one of the following: 1 (`20446`). 'Ever had prolonged feelings of sadness or depression?', and/or, 2 (`20441`). 'Ever had prolonged loss of interest in normal activities?'. $p(\text{case} \mid \text{MHQ responder} + \text{within sample}) = 0.5610$; calculated using sample size $n = 119,690$.

| Covariate | Proportion of variation in liability to depression explained (%) | Correlation with depSympt |
|---:|:---:|:---:|
| depSympt | 11.06 | 1.000 |
| Depression | 10.72 | 0.995 |
| Anxiety | 9.56 | 0.883 |
| Subjective wellbeing | 8.07 | 0.842 |
| Psychomotor cognitive | 10.44 | 0.976 |
| Neurovegetative | 10.16 | 0.955 |

Table 3: Correlation matrix for the six latent depression symptom scores of Jermy et al. [2020] ($n = 119,690$).

| | depSympt | Depression | Anxiety | Wellbeing[a] | Psychomotor[b] | Neurovegetative |
|---:|---:|---:|---:|---:|---:|---:|
| depSympt | 1.000 | 0.995 | 0.883 | 0.842 | 0.976 | 0.955 |
| Depression | 0.995 | 1.000 | 0.863 | 0.830 | 0.961 | 0.939 |
| Anxiety | 0.883 | 0.863 | 1.000 | 0.699 | 0.849 | 0.808 |
| Wellbeing[a] | 0.842 | 0.830 | 0.699 | 1.000 | 0.798 | 0.747 |
| Psychomotor[b] | 0.976 | 0.961 | 0.849 | 0.798 | 1.000 | 0.927 |
| Neurovegetative | 0.955 | 0.939 | 0.808 | 0.747 | 0.927 | 1.000 |

depSympt is our selected outcome trait. All depression scores are highly correlated with depSympt.

a: Subjective wellbeing factor of depressive symptoms, defined in main text.

b: Psychomotor cognitive factor of depressive symptoms, defined in main text.

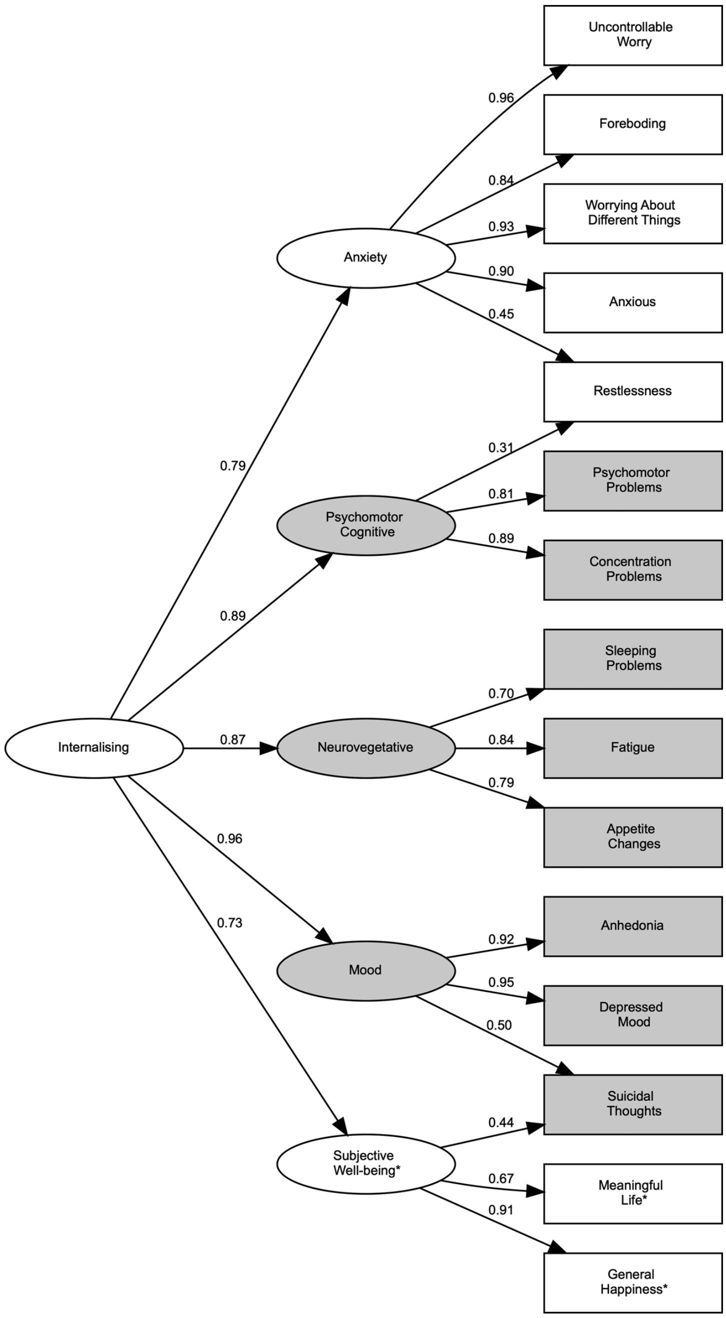

Figure 1: Visualisation for the model of depression symptom scores from Jermy et al. [2020]; exact copy of Figure 2. Note: ‘internalising’ factor in this plot is called depSympt in this work. Original caption reads: Factor model used to derive the dimensional phenotypes. As is customary in structural equation modelling graphs, circles are factors and squares are the self-reported symptoms. Shaded areas relate to either core MDD symptoms or factors containing a majority of MDD symptoms. Arrows pointing from either one factor to a symptom or a factor to another factor represent the factor loadings. *The items ‘General Happiness’ and ‘Meaningful Life’ have been reverse coded such that they explore ‘general unhappiness’ or ‘lack of meaning in one’s life’. Subjective well-being, therefore, also corresponds to a ‘subjective lack of well-being’. Nomenclature has been retained for the brevity of the labelling.

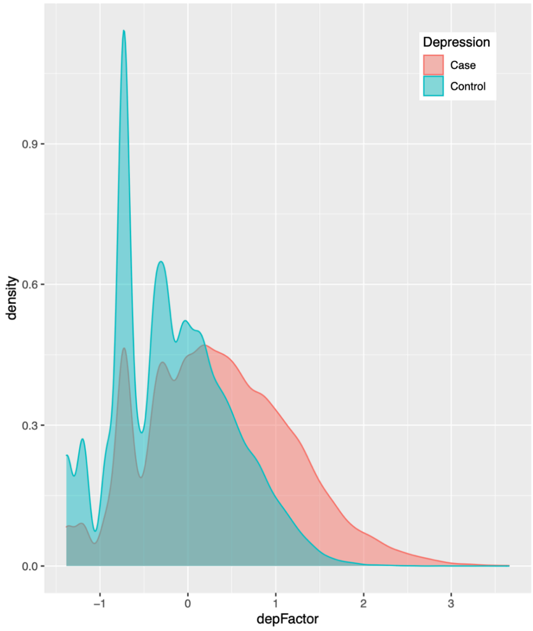

| | | depFactor | | |
|---|---|---|---|---|
| | N | Mean | SD | Median |
| MDD status | | | | |
| Case | 67,144 | 0.35 | 0.84 | 0.30 |
| Control | 52,545 | -0.18 | 0.66 | -0.25 |
| Overall | 119,689 | 0.12 | 0.81 | 0.06 |

Figure 2: depSympt density plot by major depressive disorder (MDD) status with descriptive statistics. Permutation-based hypothesis tests were performed for all listed depSympt descriptive statistics, testing the null of case-control equivalence. 100, 000 permutations were used. Empirical p-values for mean, median and standard deviation (SD) were all 0, meaning under the null no case-control differences as extreme as those observed occurred from 100, 000 samples.

#### 1.2 Polygenic risk scores

Polygenic risk scores (PRS) were calculated within the UKB dataset using PRSice v.2 [Choi and O'Reilly, 2019] for all available UKB participants who passed quality control as outlined in the main text, and with information available for the relevant mental health disorder. PRSice computes PRSs in a target sample (here the UKB cohort) by calculating the weighted sum of trait-associated alleles using summary statistics from GWAS discovery dataset(s). SNPs in linkage disequilibrium ($r^2 \geq 0.1$ [250-kb window]) were removed. We used the default average option that calculates the ratio between the PRS and the number of alleles included in each individual and scores were standardised (mean= 0, SD= 1). Where possible, the p-value threshold used was that found to be the most predictive in the study which originally generated the summary statistics. If PRSs were not generated, or at least not generated in a way requiring a p-value threshold, then PRSs were calculated using a p-value threshold equal to 0.5. Table 5 details the genome-wide association study, or meta-analysis, from which the summary statistics used in PRS generation where taken for each of the 8 included mental health disorders.

Table 5: References for summary statistics used in polygenic risk score creation.

| | | | Number of... | |
|---|---|---|---|---|
| Disorder | Summary statistics | p-value threshold | Cases | Controls |
| ADHD | Demontis et al. [2019] | 0.10 | 19,099 | 34,194 |
| Anorexia | Watson et al. [2019][a] | 0.50 | 16,991 | 56,059 |
| Anxiety | Otowa et al. [2016] | 0.20 | 7,016 | 14,745 |
| ASD | Anney et al. [2017] | 0.50 | 6,197 | 7,377 |
| Bipolar | Sklar et al. [2011] | 0.50 | 7,481 | 9,250 |
| MDD | Wray et al. [2018][b,c] | 0.05 | 45,591 | 97,674 |
| OCD | Arnold et al. [2018] | 0.01 | 2,688 | 7,037 |
| Schizophrenia | Ripke et al. [2014] | 0.05 | 33,640 | 43,456 |

*a* A small number of UKB participants are included.

*b* Used version available without UKB included.

*c* Used MDD PRS created **without** summary statistics from 23andMe.

European only samples were used in the selected summary statistics for all mental health disorders.

#### 1.3 Phenotype adjustment

All traits are adjusted for: genotype batch (data field 22000), assessment centre at which participant consented (data field 54), year of birth (data field 34), sex (data field 31), age (at interview) (data field 21003), principal components 1 to 15, and 11 variables from the MHQ relating to traumatic and stressful events occurring in adulthood or that are not captured by the childhood trauma summary variable; 'Been in a confiding relationship as an adult' (data field 20522), 'Physical violence by partner or ex (adult)' (data field

20523), 'Belittlement by partner or ex (adult)' (data field 20521), 'Sexual interference partner or ex (adult)' (data field 20524), 'Able to pay rent/mortgage as an adult' (data field 20525), 'Victim of sexual assault' (data field 20531), 'Victim of physically violent crime' (data field 20529), 'Been in serious accident believed to be life threatening?' (data field 20526), 'Witnessed sudden violent death' (data field 20530), 'Diagnosed with life-threatening illness' (data field 20528) and 'Been involved in combat or in a war-zone' (data field 20527).

Table 6 describes the additional adjustments made for depSympt during interaction analysis with each covariate trait in turn. The covariate is adjusted for all the same variables as depSympt, except for itself.

Average sleep duration is known to have a non-linear relationship with depression symptoms, with both too little and too much sleep being symptoms of depression. It is possible that other continuous traits also have a non-linear relationship with depSympt. Therefore, with the exception of the principal components, all continuous fixed effects variables used are allowed to have a non-linear relationship with depSympt and the covariate traits by using fractional polynomials (FPs) [Royston and Altman, 1994]. To do this we used the `R` package `mfp` [Benner and Ambler, 2015] within a generalised linear model (`stats::glm` [R Core Team, 2020]). When specified, this package explores the relationship between an outcome and a continuous covariate by testing for suitable (power and log based) transformations of the covariate that best explain the relationship between this variable and the outcome. We allow up to two fractional polynomial terms (transformations of each continuous covariate) to be included. It is possible for the `mfp` package to select no relationship between a variable and the outcome. If this occurs we still include the untransformed variable as a linear term in the final fixed effects model. See Benner and Ambler [2015] for full details on which power transformations are tested for, and how, when using fractional polynomials.

Prior to transformation via fractional polynomials, all biomarkers except LDL were log-transformed. Log-transforming biomarkers is typically done and after inspecting the distribution plots of the untransformed and log-transformed biomarkers (see Supplementary Figures 28 - 32) we concluded that only LDL had a distribution obviously closer to normality on the untransformed scale.

Table 6: Additional variables used in fixed effects adjustment of depSympt for all interaction analyses (defined by the covariate trait).

| Covariate trait | Additional variables for depSympt fixed effects models |
|---|---|
| BMI | BMI, TDI, sleep, childhood trauma, MET (total) |
| TDI | BMI, TDI, sleep, childhood trauma, MET (total) |
| Sleep | BMI, TDI, sleep, childhood trauma, MET (total) |
| Childhood trauma | BMI, TDI, sleep, childhood trauma, MET (total) |
| MET (total) | BMI, TDI, sleep, childhood trauma, MET (total) |
| MET (walk) | BMI, TDI, sleep, childhood trauma, MET (walk) |
| MET (mod) | BMI, TDI, sleep, childhood trauma, MET (mod) |
| MET (vig) | BMI, TDI, sleep, childhood trauma, MET (vig) |
| Waist circumference | Waist circumference, TDI, sleep, childhood trauma, MET (total) |
| Waist to hip ratio | Waist to hip ratio, TDI, sleep, childhood trauma, MET (total) |
| MDD PRS | MDD PRS, BMI, TDI, sleep, childhood trauma, MET (total) |
| Scz PRS | Scz PRS, BMI, TDI, sleep, childhood trauma, MET (total) |
| BIP PRS | BIP PRS, BMI, TDI, sleep, childhood trauma, MET (total) |
| ASD PRS | ASD PRS, , BMI, TDI, sleep, childhood trauma, MET (total) |
| Anorexia PRS | Anorexia PRS, BMI, TDI, sleep, childhood trauma, MET (total) |
| Anxiety PRS | Anxiety PRS, BMI, TDI, sleep, childhood trauma, MET (total) |
| ADHD PRS | ADHD PRS, BMI, TDI, sleep, childhood trauma, MET (total) |
| OCD PRS | OCD PRS, BMI, TDI, sleep, childhood trauma, MET (total) |
| log-CRP | log-CRP, LDL, log-triglycerides, log-vitamin D, BMI, TDI, sleep, childhood trauma, MET (total) |
| LDL | log-CRP, LDL, log-triglycerides, log-vitamin D, BMI, TDI, sleep, childhood trauma, MET (total) |
| log-Triglycerides | log-CRP, LDL, log-triglycerides, log-vitamin D, BMI, TDI, sleep, childhood trauma, MET (total) |
| log-Vitamin D | log-CRP, LDL, log-triglycerides, log-vitamin D, BMI, TDI, sleep, childhood trauma, MET (total) |
| log-HDL | log-HDL, log-CRP, LDL, log-triglycerides, log-vitamin D, BMI, TDI, sleep, childhood trauma, MET (total) |
| Neuroticism | Neuroticism, BMI, TDI, sleep, childhood trauma, MET (total) |
| Smoking | Smoking, BMI, TDI, sleep, childhood trauma, MET (total) |

Sleep = average sleep duration. MET (total) = Summed MET minutes per week all activities. MET (walk) = Summed MET minutes per week walking. MET (mod) = Summed MET minutes per week moderate. MET (vig) = Summed MET minutes per week vigorous. PRS = polygenic risk score. MDD = major depressive disorder. Scz = schizophrenia. BIP = bipolar. ADHD = attention deficit hyperactivity disorder. ASD = autism spectrum disorder. OCD = obsessive compulsive disorder. CRP = C-reactive protein.

#### 1.4 Multivariate reaction norm model

##### 1.4.1 Model introduction

Developed within studies of ecology and agriculture, the reaction norm (RN) is a function characterising phenotypic plasticity; that is, how the observed phenotype of a given genotype (individual) changes when moving along an environmental gradient. Non-parallel RNs indicate the presence of genotype-environment interactions. Population properties can be studied using a collection, or bundle, of RNs via a RN model (RNM). RNMs estimate: 1. the average outcome trait value for a given covariate trait value (the estimated trend between trait and environment regardless of genotype via a fixed effects model), and 2. the residual outcome trait variability for each environmental value allowing investigation of genotype-environment interactions via a random effects model (non-parallel RNs due to the presence of gene-environment interactions will produce heterogeneity in outcome variance across the environmental gradient). RNMs are therefore a type of mixed effects model capturing average trend via the fixed effects model and residual heterogeneity via the random effects model.

RNs can be obtained experimentally within many plant and animal studies. This is not generally possible for human studies; we do not typically observe an individuals phenotypic response to varying levels of an environmental exposure, whilst controlling for confounders. Instead, a single point on each individuals RN is typically available (we observe one outcome value and one covariate trait for each individual in the study population). However, a RNM can still be constructed using estimated genetic similarities from genome-wide SNP data within a random regression model [Schaeffer, 2004, Jarquin et al., 2014, Ni et al., 2019].

In this approach, after adjusting for average trends in the outcome trait across all genotypes using a linear regression model (the fixed effects model), the variance of the outcome trait ($Y$) is decomposed into genetic and residual components. The genetic component captures the proportion of variability in $Y$ attributable to the measured genetic variables. This will be the SNP heritability, which is the correlation between the estimated genetic sharing (defined by the genetic relationship matrix) and phenotypic sharing [Hall and Bush, 2016]. In the RNM these variance components are further decomposed such that they are functions of the covariate trait ($C$), allowing the SNP heritability and the residual variance component for outcome $Y$ to vary with respect to $C$, thereby incorporating a genome-wide genotype-covariate (G-C) and residual-covariate (R-C) interaction.

Like in the RNMs applied to animal/plant studies, RNMs within human GWAS are looking at the average trend and the trend in outcome variability across a covariate. In controlled experiments we can be certain that the cause of the trend in phenotypic variability is due to G-C interactions. In observational data, where we cannot control for other sources of variation within the study design, this is not certain. Therefore, we need to estimate what proportion of this trend is attributable to genetic and non-genetic sources by using a measure of genetic similarity.

In the MRNM the correlation between the outcome and covariate trait is modelled. This is done by incorporating a second random effects model for the covariate trait, $C$. The term multivariate therefore refers to two random effects models being considered jointly; one for $Y$ and one for $C$. As before we: 1. adjust $C$ for average trends using a fixed effects model, and, 2. decompose the residual variation into genetic and residual components. Unlike for outcome trait $Y$ we do not further decompose these random components, and are therefore estimating the SNP heritability and residual variance component for $C$. We will discuss in the next section, which mathematically defines the model, how using a random effects models for both $Y$ and $C$ can estimate and control for genetic and residual correlations between the two traits.

##### 1.4.2 Model definition

Focusing on the random effects model, because this is where the interactions are modelled, we assume that $Y$ ($C$) refers to the outcome (covariate) trait that has been pre-adjusted for fixed effects using a linear model, and then standardised. For completeness, that is for each individual $i$:

$$Y_i = \frac{Y_i^o - E[Y_i^o|\underline{X}_i = \underline{x}_i, C_i^o = c_i^o]}{\sqrt{Var[Y_i^o|\underline{X}_i = \underline{x}_i, C_i^o = c_i^o]}} \sim N(0,1) \tag{1}$$

and:

$$C_i = \frac{C_i^o - E[C_i^o|\underline{X}_i = \underline{x}_i]}{\sqrt{Var[C_i^o|\underline{X}_i = \underline{x}_i]}} \sim N(0,1) \tag{2}$$

where:

- $Y_i^o$ ($C_i^o$) is the original outcome (covariate) trait, prior to fixed effects adjustment, for individual $i$,
- $\underline{X}_i$ is a vector of random variables selected for inclusion in the fixed effects model for both $Y$ and $C$,
- $\underline{x}_i$ is a vector of observed variables for individual $i$,
- $E[Y_i^o|\underline{X}_i = \underline{x}_i, C_i^o = c_i^o]$ is the estimated value of $Y_i^o$ from a linear model including predictors $\underline{X}_i$ and $C_i^o$,
- $Var[Y_i^o|\underline{X}_i = \underline{x}_i, C_i^o = c_i^o]$ is the residual variation from this fixed effects model,
- $E[C_i^o|\underline{X}_i = \underline{x}_i]$ is the estimated value of $C_i^o$ from a linear model including predictors $\underline{X}_i$, and,
- $Var[C_i^o|\underline{X}_i = \underline{x}_i]$ is the residual variation from this fixed effects model.

Why do we use standardised traits? In this paper we have multiple covariate traits, whose G-C and R-C interactions with respect to the outcome trait are explored in separate MRNMs. Standardising the covariate traits allows us to compare their relative importance in explaining the variability in the outcome trait across models. Standardising the outcome trait allows the comparison of the impact of covariate traits across outcome traits. Here, we only have one outcome trait (depSympt), but: 1. results from the model using the standardised outcome trait allows comparison across studies, and, 2. the proportion of the variability in raw depSympt attributable to changes in the standard deviation of the covariate trait can be obtained from a model using standardised depSympt as the outcome (see Supplementary Section 1.3 for details).

For each individual $i$ in a sample of size $N$, we define the (random effects part of the) MRNM as:

$$\begin{bmatrix} Y_i|C_i = c_i \\ C_i \end{bmatrix} = \begin{bmatrix} \alpha_{0i} + \alpha_{1i}c_i \\ \beta_{0i} \end{bmatrix} + \begin{bmatrix} \tau_{0i} + \tau_{1i}c_i \\ \epsilon_{0i} \end{bmatrix}$$

where:

- $\alpha_{0i} \sim N(0, \sigma^2_{\alpha_0})$ is the random effect coefficient describing the random genetic intercept for $Y_i$. It is a random variable for the main genetic effect for individual $i$, describing the relationship between the measured genetic variables for this individual and their standardised deviation from the expected outcome trait value, which does not change with $C_i$.

- $\alpha_{1i} \sim N(0, \sigma^2_{\alpha_1})$ is the random effect coefficient describing a random genetic slope across $C_i$ for individual $i$. This is the G-C interaction term, and is a random variable describing the relationship between the measured genetic variables for this individual and their standardised deviation from the expected outcome trait value, which can vary across $C_i$.

- $\tau_{0i} \sim N(0, \sigma^2_{\tau_0})$ is the random effect coefficient describing the residual random intercept for $Y_i$. It is a random variable describing the standardised residual deviation from the expected value for this individual, that is not accounted for by the measured genetic variables and which does not vary with $C_i$.

- $\tau_{1i} \sim N(0, \sigma^2_{\tau_1})$ is the random effect coefficient describing the residual random slope for individual $i$. It is a random variable describing the standardised residual deviation from the expected value for individual $i$, that is not accounted for by the measured genetic variables but which does vary with $C_i$. This is the R-C interaction.

- $\beta_{0i} \sim N(0, \sigma^2_{\beta_0})$ is the random effect coefficient describing the random genetic intercept for $C_i$. It is a random variable describing the relationship between the measured genetic variables for individual $i$ and their standardised deviation for the covariate trait. The population distribution parameter $\sigma^2_{\beta_0}$ is the SNP heritability for the covariate trait given the fixed effects model.

- $\epsilon_{0i} \sim N(0, \sigma^2_{\epsilon_0})$ is the random effect coefficient describing the residual random intercept for $C_i$. It is a random variable describing the standardised deviation of the covariate trait for individual $i$ that is not explained by their measured genetic variables.

Here, unlike in a fixed effects model, each individual in the sample has his or her own set of random variables (random effects) to capture heterogeneity. Although each individual has their own random effects, these are assumed to follow the same *population* distribution. Here this distribution is assumed to be multivariate normal with mean equal to zero. Therefore, to parameterise the random effects model, we need to estimate the unknown variance-covariance parameters for the random effects. We shall now show this explicitly by writing the model within the study population. The above random effects model can be written in matrix form for the complete sample as follows:

$$\begin{bmatrix} \underline{Y}|\underline{C} = \underline{c} \\ \underline{C} \end{bmatrix} = \begin{bmatrix} \underline{\alpha}_0 + \underline{\alpha}_1 \underline{c} \\ \underline{\beta}_0 \end{bmatrix} + \begin{bmatrix} \underline{\tau}_0 + \underline{\tau}_1 \underline{c} \\ \underline{\epsilon}_0 \end{bmatrix}$$

such that:

$$\begin{bmatrix} \underline{Y}|\underline{C} = \underline{c} \\ \underline{C} \end{bmatrix} \sim N\left( \begin{bmatrix} \underline{0} \\ \underline{0} \end{bmatrix}, \Sigma \right)$$

where:

$$\Sigma = \begin{bmatrix} \Sigma_{\underline{Y}} & \Sigma_{\underline{Y},\underline{C}} \\ \Sigma_{\underline{C},\underline{Y}} & \Sigma_{\underline{C}} \end{bmatrix}$$

As you can see, for a random effects model, estimating the unknown population model parameters are contained within the covariance matrix $\Sigma$, which defines the variance-covariance of the conditional outcome trait and the covariate trait for the $N$ individuals in the study population.

***Functions defining the covariance matrix***

*1. Defining $\Sigma_{\underline{Y}}$*

$\Sigma_{\underline{Y}}$ is the variance-covariance matrix for $\underline{Y}|\{\underline{C} = \underline{c}\}$, describing the covariance between the standardised residual outcome trait for all individuals, where the $i^{th}$ row and $j^{th}$ column is defined as:

$$\begin{aligned} \Sigma_{\underline{Y}}(i,j) &= Cov[Y_i|\{C_i = c_i\}, Y_j|\{C_j = c_j\}] \\ &= \underline{A}(i,j)\sigma_{g_{Y,i},g_{Y,j}} + \underline{I}(i,j)\sigma_{e_{Y,i},e_{Y,j}} \end{aligned}$$

where $\sigma_{g_{Y,i},g_{Y,j}}$ is the covariance between the random genetic components of trait $Y$ for two values of the covariate trait denoted by $c_i$ and $c_j$ which is:

$$\sigma_{g_{Y,i},g_{Y,j}} = \sigma^2_{\alpha_0} + (c_i + c_j)\sigma_{\alpha_0,\alpha_1} + c_i c_j \sigma^2_{\alpha_1}$$

Similarly, $\sigma_{e_{Y,i},e_{Y,j}}$ is the covariance between the random residual components of trait $Y$ for two values of the covariate trait denoted by $c_i$ and $c_j$ which is:

$$\sigma_{e_{Y,i},e_{Y,j}} = \sigma^2_{\tau_0} + (c_i + c_j)\sigma_{\tau_0,\tau_1} + c_i c_j \sigma^2_{\tau_1}$$

$\underline{A}(i,j)$ the value contained within the $i^{th}$ row and $j^{th}$ column of the GRM, corresponding to the average (measured) genetic sharing between individuals $i$ and $j$. $\underline{A}(i,i) = 1$. $\underline{I}(i,j)$ is the value in the $i^{th}$ row and $j^{th}$ column of the identity matrix. This will be 0 when $i \neq j$, meaning we assume there is no environmental sharing between individuals in this model.

We note that the above leads to a conditional variance estimate for trait $Y_i$ of:

$$Var[Y_i|C_i = c_i] = \sigma^2_{\alpha_0} + 2c_i\sigma_{\alpha_0,\alpha_1} + c_i^2\sigma^2_{\alpha_1} + \sigma^2_{\tau_0} + 2c_i\sigma_{\tau_0,\tau_1} + c_i^2\sigma^2_{\tau_1}$$

and, assuming $E[C_i] = 0$ and $Var[C_i] = 1$, an unconditional variance estimate of:

$$Var[Y_i] = \sigma^2_{\alpha_0} + \sigma^2_{\alpha_1} + \sigma^2_{\tau_0} + \sigma^2_{\tau_1}$$

for all individuals in the population.

Since $Var[Y_i] = 1$, the variance estimates for $\sigma^2_{\alpha_0}$, $\sigma^2_{\alpha_1}$, $\sigma^2_{\tau_0}$ and $\sigma^2_{\tau_1}$ represent a measure of the importance for that variance component (main genetic, G-C, main residual and R-C) in explaining the variability of the outcome trait in the population.

*2. Defining* $\Sigma_{\underline{Y},\underline{C}}$

$\Sigma_{\underline{Y},\underline{C}} = \Sigma^T_{\underline{C},\underline{Y}}$ is the covariance matrix for $\underline{Y}|\{\underline{C} = \underline{c}\}$ and $\underline{\text{C}}$, where the $i^{th}$ row and $j^{th}$ column is defined as:

$$\begin{aligned}
\Sigma_{\underline{Y},\underline{C}}(i,j) &= Cov[Y_i|\{C_i = c_i\}, C_j] \\
&= \underline{A}(i,j)\Big(\sigma_{\alpha_0,\beta_0} + c_i\sigma_{\alpha_1,\beta_0}\Big) + \underline{I}(i,j)\Big(\sigma_{\tau_0,\epsilon_0} + c_i\sigma_{\tau_1,\epsilon_0}\Big)
\end{aligned}$$

$\Sigma_{\underline{Y},\underline{C}}(i,j)$ contains the covariance between the conditional outcome trait for individual $i$ (given we observe their covariate trait) and the covariate trait for individual $j$. It is the sum of the genetic and the residual covariances between the traits when $i = j$. It is assumed that when individual $i \neq j$, $\Sigma_{\underline{Y},\underline{C}}(i,j)$ is equal to the genetic covariance between the traits only. That is, residual variation is explained by independent variables for each individual, and there are no un-modelled correlations, within the environment for example, between individuals that explain any covariation between the outcome of one individual and the covariate of another (we note that there is a modelled relationship between the variance of $Y_i$ and $C_i$).

The genetic covariance is a function of the measured trait value ($c_i$), the measured genetic sharing and the population random effects covariances $\sigma_{\alpha_0,\beta_0}$ and $\sigma_{\alpha_1,\beta_0}$ (which are to be estimated). $\sigma_{\alpha_0,\beta_0}$ defines how the main genetic effect for $Y|C = c$ covaries with the main genetic effect of $C$, and $\sigma_{\alpha_1,\beta_0}$ defines how the G-C interaction effect for $Y|C = c$ covaries with the main genetic effect of $C$.

The residual covariance, only used within an individual, is a function of the measured trait value ($c_i$) and the population random effects covariances $\sigma_{\tau 0,\epsilon 0}$ and $\sigma_{\tau 1,\epsilon 0}$ (which are to be estimated). $\sigma_{\tau 0,\epsilon 0}$ defines how the main residual random effect for $Y|C = c$ covaries with the main residual effect of $C$, and $\sigma_{\tau 1,\epsilon 0}$ defines how the R-C interaction random effect for $Y|C = c$ covaries with the main genetic effect of $C$, but only within an individual (not between individuals). Estimating these parameters allows for residual covariance between outcome and the covariate trait that is otherwise un-modelled. If these covariances were not included, genetic interaction variances may be inflated.

*3. Defining* $\Sigma_{\underline{C}}$

$\Sigma_{\underline{C}}$ is the variance-covariance matrix for $\underline{C}$, where the $i^{th}$ row and $j^{th}$ column is defined as:

$$\begin{aligned}\Sigma_{\underline{C}}(i,j) &= Cov[C_i, C_j] \\ &= \underline{A}(i,j)\sigma^2_{\beta_0} + \underline{I}(i,j)\sigma^2_{\epsilon 0}\end{aligned}$$

These variance-covariance matrices defining $\Sigma$ can also be written in matrix form. Please see Ni et al. [2019], the methods paper which first described the MRNM within human GWAS, for details of this. For information about the (restricted) maximum likelihood estimation process for these covariance parameters please see Lee and van der Werf [2016], which describes the `mtg2` software package and its corresponding manual found here: `https://sites.google.com/site/honglee0707/mtg2`.

Here, we just note that the MRNM is parameterised by estimating the following covariance matrix between the random effects (RE), which represent sources of (co)variation for $\underline{Y}|\underline{C} = \underline{c}$ and $\underline{C}$:

$$\Sigma_{RE} = \begin{bmatrix} \sigma^2_{\alpha 0} & \sigma_{\alpha 0,\alpha 1} & \sigma_{\alpha 0,\tau 0} & \sigma_{\alpha 0,\tau 1} & \sigma_{\alpha 0,\beta 0} & \sigma_{\alpha 0,\epsilon 0} \\ \sigma_{\alpha 0,\alpha 1} & \sigma^2_{\alpha 1} & \sigma_{\alpha 1,\tau 0} & \sigma_{\alpha 1,\tau 1} & \sigma_{\alpha 1,\beta 0} & \sigma_{\alpha 1,\epsilon 0} \\ \sigma_{\alpha 0,\tau 0} & \sigma_{\alpha 1,\tau 0} & \sigma^2_{\tau 0} & \sigma_{\tau 0,\tau 1} & \sigma_{\tau 0,\beta 0} & \sigma_{\tau 0,\epsilon 0} \\ \sigma_{\alpha 0,\tau 1} & \sigma_{\alpha 1,\tau 1} & \sigma_{\tau 0,\tau 1} & \sigma^2_{\tau 1} & \sigma_{\tau 1,\beta 0} & \sigma_{\tau 1,\epsilon 0} \\ \sigma_{\alpha 0,\beta 0} & \sigma_{\alpha 1,\beta 0} & \sigma_{\tau 0,\beta 0} & \sigma_{\tau 1,\beta 0} & \sigma^2_{\beta 0} & \sigma_{\beta 0,\epsilon 0} \\ \sigma_{\alpha 0,\epsilon 0} & \sigma_{\alpha 1,\epsilon 0} & \sigma_{\tau 0,\epsilon 0} & \sigma_{\tau 1,\epsilon 0} & \sigma_{\beta 0,\epsilon 0} & \sigma^2_{\epsilon 0} \end{bmatrix} = \begin{bmatrix} \sigma^2_{\alpha 0} & \sigma_{\alpha 0,\alpha 1} & 0 & 0 & \sigma_{\alpha 0,\beta 0} & 0 \\ \sigma_{\alpha 0,\alpha 1} & \sigma^2_{\alpha 1} & 0 & 0 & \sigma_{\alpha 1,\beta 0} & 0 \\ 0 & 0 & \sigma^2_{\tau 0} & \sigma_{\tau 0,\tau 1} & 0 & \sigma_{\tau 0,\epsilon 0} \\ 0 & 0 & \sigma_{\tau 0,\tau 1} & \sigma^2_{\tau 1} & 0 & \sigma_{\tau 1,\epsilon 0} \\ \sigma_{\alpha 0,\beta 0} & \sigma_{\alpha 1,\beta 0} & 0 & 0 & \sigma^2_{\beta 0} & 0 \\ 0 & 0 & \sigma_{\tau 0,\epsilon 0} & \sigma_{\tau 1,\epsilon 0} & 0 & \sigma^2_{\epsilon 0} \end{bmatrix}$$

Although some of these model parameters are variances, and so should be $> 0$, the algorithm estimating these parameters does not know that they should be constrained. Negative variance estimates are therefore possible. Typically this is just an underestimation of a small, or zero, variance. We therefore calculate confidence intervals for variance parameters. A variance estimate with a 95% confidence interval that overlaps with zero indicates a lack of confidence that the random effect it corresponds to is useful in explaining phenotypic variation.

***The variance-covariance random effects parameters***

In this work some covariance parameters are assumed to be zero, as indicated in the definition of $\Sigma_{RE}$ above. In particular, within each trait and between traits, we assume that the genetic random effects are uncorrelated with the residual random effects.

$\sigma^2_{\beta_0}$ ($= Var[\beta_0]$) is the SNP heritability for the standardised covariate trait, $C$. $\sigma^2_{\epsilon_0}$ is the proportion of variability in $C$ not captured by the measured genetic variables; it is residual.

Using the above MRNM, the (measured) genetic and residual variance components for $Y_i$ are a function of $C_i$, such that:

$$Var[Y_i|C_i = c_i] = V_{G_i|C_i=c_i} + V_{R_i|C_i=c_i} \tag{3}$$

$$\begin{aligned} V_{G_i|C_i=c_i} &= \sigma^2_{\alpha_0} + 2c_i\sigma_{\alpha_0,\alpha_1} + c_i^2\sigma^2_{\alpha_1} \\ V_{R_i|C_i=c_i} &= \sigma^2_{\tau_0} + 2c_i\sigma_{\tau_0,\tau_1} + c_i^2\sigma^2_{\tau_1} \end{aligned}$$

for all individuals in the population. $V_{G_i|C_i=c_i}$ ($V_{R_i|C_i=c_i}$) is the genetic (residual) variance component for $Y_i|\{C_i = c_i\}$. The MRNM in Equation (3) specifies that the phenotypic variability in $Y_i$ is a degree 2 polynomial function, which will equal $\sigma^2_{\alpha_0} + \sigma^2_{\tau_0}$ when $c_i = 0$ (the mean). Therefore, $\sigma^2_{\alpha_0}$ can be thought of as the polygenic variance component of $Y$ for individuals with the covariate trait equal to the expected from the fixed effects model. $\sigma^2_{\alpha_1}$ and $\sigma_{\alpha_0,\alpha_1}$ determine the change in $V_{G_i|C_i=c_i}$ for different values of $C_i$ with larger (absolute) values for these variance-covariance model parameters indicating a larger differences in the polygenic variance component of $Y$ for larger deviations in the covariate trait from its mean.

$\sigma^2_{\alpha_1}$ in particular is an important measure for strength of the G-C interaction, highlighted by its role in the unconditional variance of $Y_i$ defined as:

$$Var[Y_i] = V_{G_i} + V_{R_i} = 1$$

$$\begin{aligned} V_{G_i} &= \sigma^2_{\alpha_0} + \sigma^2_{\alpha_1} \\ V_{R_i} &= \sigma^2_{\tau_0} + \sigma^2_{\tau_1} \end{aligned}$$

$\sigma^2_{\alpha_1}$ is part of the SNP heritability for $Y$ determined by variation in $C$. The larger the $\sigma^2_{\alpha_1}$ value, the more important the G-C interaction is in explaining the variability observed in the standardised outcome trait.

Similarly, $V_{R_i|C_i=c_i}$ is the residual variance component for $Y_i|\{C_i = c_i\}$, which will equal $\sigma^2_{\tau_0}$ when $C = 0$. The variance-covariance parameters $\sigma^2_{\tau_1}$ and $\sigma_{\tau_0,\tau_1}$ determine the change in $V_{R_i|C_i=c_i}$ for changes in $C_i$, with $\sigma^2_{\tau_1}$ being a measure of the importance of the R-C interaction in explaining the variability in $Y_i$.

Correlation between $Y_i$ and $C_i$ is incorporated into the model by allowing non-zero covariance parameters between the genetic components of the 2 traits ($\sigma_{\alpha_0,\beta_0}$ and $\sigma_{\alpha_1,\beta_0}$), and the residual components of the two traits ($\sigma_{\tau_0,\epsilon_0}$ and $\sigma_{\tau_1,\epsilon_0}$). This is an important advantage of the MRNM because not accounting for genetic and residual correlations between traits could lead to an inflation in the strength of the interactions [Ni et al., 2019].

#### 1.5 Meta-analysis methods

##### 1.5.1 Likelihood ratio test

Let $p_{jk}$ be the likelihood ratio p-value for the $j^{th}$ covariate trait ($j = 1, 2, ..., 25$) and the $k^{th}$ subset of data ($k = 1, 2, 3$), when testing the hypothesis:

$$H_0: \ \sigma^2_{\alpha_{1jk}} = \sigma^2_{\tau_{1jk}} = \sigma_{\alpha_{0jk},\alpha_{1jk}} = \sigma_{\tau_{0jk},\tau_{1jk}} = \sigma_{\alpha_{1jk},\beta_{0jk}} = \sigma_{\tau_{1jk},\epsilon_{0jk}} = 0$$
$$H_1: \ \text{otherwise}$$

Using Fishers method [Evangelou and Ioannidis, 2013], the combined/ meta test statistic for covariate trait $j$ is:

$$\chi^2_j = -2\sum_{k=1}^{K} log(p_{jk})$$

where $K = 3$, and is the number of datasets to combine, and degrees of freedom, $df$, equals $2K$. The meta p-value is then calculated using the $\chi^2$ distribution. An example of the `R` code is:

```
pchisq(q = ts_j, df=6, lower.tail=F)
```

where `ts_j` equals $\chi^2_j$ above.

##### 1.5.2 Variance-covariance model parameters: estimates and standard errors

The random effects part of the MRNM is parameterised by variance-covariance parameters that are estimated in each data subset, along with standard errors. To combine these parameter estimates and standard errors to obtain meta-analysed estimates we used the following method. Taking $\sigma^2_{\alpha_{0j}}$ as an example, where $j$ denotes the $j^{th}$ covariate trait, we use:

$$\hat{\sigma}^2_{\alpha_{0j}} = \frac{\sum_{k=1}^{K} \hat{\sigma}^2_{\alpha_{0jk}} SE(\hat{\sigma}^2_{\alpha_{0jk}})^{-1}}{\sum_{k=1}^{K} SE(\hat{\sigma}^2_{\alpha_{0jk}})^{-1}}$$

to obtain the meta-analysed model parameter estimate, and:

$$SE(\hat{\sigma}_{\alpha_{0j}}) = \frac{1}{\sum_{k=1}^{K} SE(\hat{\sigma}^2_{\alpha_{0jk}})^{-1}}$$

to obtain the meta-analysed standard error for the model parameter. This is a fixed effects meta-analysis method used by Ni et al. [2019]. It provides a weighted mean of the parameter estimates giving more weight to data subsets with smaller standard error estimates [Hedges and Vevea, 1998]. Wald confidence intervals are presented (i.e. using a normal distribution) as is done in meta-analysis packages such as `R::metafor` [Viechtbauer, 2010].

##### 1.5.3 Genetic, residual and total variance components for standardised residual depSympt as a function of the covariate trait: estimate and SEs

Assume that we have adjusted both depSympt and the $j^{th}$ covariate trait for their respective fixed effects model, and then standardised, such that for an individual $i$:

- the outcome trait we are considering is $Y_i$ as defined in Equation (1), and,
- the covariate trait we are considering is $C_{ij}$ as defined in Equation (2).

For a given individual $i$, it is useful to understand the estimated relationship between expected variability in standardised residual depSympt ($Y_i$) and the standardised residual covariate trait ($C_{ij}$), and to break this relationship into the genetic variance component and the residual variance component, as well as obtain standard errors (SEs) for these variance components across $C_{ij}$. Plotting these relationships (as we have done in Figure 3 presented within the main text for average sleep duration, or for the other covariates considered in Supplementary Figures 35 - **??**), allows researchers to better understand the MRNM output, including the relative contributions to total variation from the genetic component compared to the residual, how this changes across the covariate trait and if confidence intervals overlap with each other, or with 0.

Focusing on the part of the MRNM considered here which defined $Y_i$, we recall that:

$$Y_i|\{C_{ij} = c_{ij}\} = \alpha_{0i} + \alpha_{1i}c_{ij} + \tau_{0i} + \tau_{1i}c_{ij}$$

where the random effects follow a multivariate normal distribution, with zero-mean and covariances between genetic and nongenetic random effects fixed at 0. Using this we can write the following equation for the variance of $Y_i|\{C_{ij} = c_{ij}\}$:

$$\begin{aligned} V_{Y_i|C_{ij}=c_{ij}} &= Var[Y_i|C_{ij} = c_{ij}] \\ &= E[(Y_i|C_{ij} = c_{ij})^2] - (E[Y_i|C_{ij} = c_{ij}])^2 \\ &= E[(Y_i|C_{ij} = c_{ij})^2] \\ &= V_{G_i|C_{ij}=c_{ij}} + V_{R_i|C_{ij}=c_{ij}} \end{aligned}$$

where:

$$V_{G_i|C_{ij}=c_{ij}} = \sigma^2_{\alpha_0} + 2c_{ij}\sigma_{\alpha_0,\alpha_1} + c^2_{ij}\sigma^2_{\alpha_1}$$

is the genetic variance component for conditional $Y_i$, and:

$$V_{R_i|C_{ij}=c_{ij}} = \sigma^2_{\tau_0} + 2c_{ij}\sigma_{\tau_0,\tau_1} + c^2_{ij}\sigma^2_{\tau_1}$$

is the residual variance component for conditional $Y_i$ (as defined in the MRNM definition section). We note the above is not considering correlation between individuals under-study, and rather focuses within a given individual. An estimate for the total, genetic and residual variance components will be obtained using: $[\hat{\sigma}^2_{\alpha_0}, \hat{\sigma}_{\alpha_0,\alpha_1}, \hat{\sigma}^2_{\alpha_1}, \hat{\sigma}^2_{\tau_0}, \hat{\sigma}_{\tau_0,\tau_1}, \hat{\sigma}^2_{\tau_1}]$, which are variance component estimates outputted by the MRNM (mtg2 package [Lee and van der Werf, 2016]).

Additionally, the inverse Fisher information matrix is outputted by mtg2. This matrix is used to estimate the standard errors of the estimated variance components. Extracting the elements from this matrix relating to the variance components required, and using the delta method, the standard error for the genetic variance component for $Y_i$ given $C_{ij} = c_{ij}$ is given by:

$$SE(\hat{V}_{G_i|C_{ij}=c_{ij}}) = \nabla^T_{V_{G_i|C_{ij}=c_{ij}}} \hat{\Sigma}_{V_{G|C_j=c_j}} \nabla_{V_{G_i|C_{ij}=c_{ij}}}$$

where:

$$\nabla_{V_{G_i|C_{ij}=c_{ij}}} = \begin{bmatrix} 1 \\ 2c_{ij} \\ c^2_{ij} \end{bmatrix}$$

and:

$$\hat{\Sigma}_{V_{G|C_j=c_j}} = \begin{bmatrix} Var[\hat{\sigma}^2_{\alpha_0}] & Cov[\hat{\sigma}^2_{\alpha_0}, \hat{\sigma}_{\alpha_0,\alpha_1}] & Cov[\hat{\sigma}^2_{\alpha_0}, \hat{\sigma}^2_{\alpha_1}] \\ Cov[\hat{\sigma}_{\alpha_0,\alpha_1}, \hat{\sigma}^2_{\alpha_0}] & Var[\hat{\sigma}_{\alpha_0,\alpha_1}] & Cov[\hat{\sigma}_{\alpha_0,\alpha_1}, \hat{\sigma}^2_{\alpha_1}] \\ Cov[\hat{\sigma}^2_{\alpha_1}, \hat{\sigma}^2_{\alpha_0}] & Cov[\hat{\sigma}^2_{\alpha_1}, \hat{\sigma}_{\alpha_0,\alpha_1}] & Var[\hat{\sigma}^2_{\alpha_1}] \end{bmatrix}$$

Similarly, the standard error for the residual variance component for $Y_i$ given $C_{ij} = c_{ij}$ is given by:

$$SE(\hat{V}_{R_i|C_{ij}=c_{ij}}) = \nabla^T_{V_{R_i|C_{ij}=c_{ij}}} \hat{\Sigma}_{V_{R|C_j=c_j}} \nabla_{V_{R_i|C_{ij}=c_{ij}}}$$

where:

$$\nabla_{V_{R_i|C_{ij}=c_{ij}}} = \begin{bmatrix} 1 \\ 2c_{ij} \\ c^2_{ij} \end{bmatrix}$$

and:

$$\hat{\Sigma}_{V_{R|C_j=c_j}} = \begin{bmatrix} Var[\hat{\sigma}^2_{\tau_0}] & Cov[\hat{\sigma}^2_{\tau_0}, \hat{\sigma}_{\tau_0,\tau_1}] & Cov[\hat{\sigma}^2_{\tau_0}, \hat{\sigma}^2_{\tau_1}] \\ Cov[\hat{\sigma}_{\tau_0,\tau_1}, \hat{\sigma}^2_{\tau_0}] & Var[\hat{\sigma}_{\tau_0,\tau_1}] & Cov[\hat{\sigma}_{\tau_0,\tau_1}, \hat{\sigma}^2_{\tau_1}] \\ Cov[\hat{\sigma}^2_{\tau_1}, \hat{\sigma}^2_{\tau_0}] & Cov[\hat{\sigma}^2_{\tau_1}, \hat{\sigma}_{\tau_0,\tau_1}] & Var[\hat{\sigma}^2_{\tau_1}] \end{bmatrix}$$

The standard error for the total variability for $Y_i$ given $C_{ij} = c_{ij}$ is given by:

$$SE(\hat{V}_{Y_i|C_{ij}=c_{ij}}) = \nabla^T_{V_{Y_i|C_{ij}=c_{ij}}} \hat{\Sigma}_{V_{Y|C_j=c_j}} \nabla_{V_{Y_i|C_{ij}=c_{ij}}}$$

where:

$$\nabla_{V_{Y_i|C_{ij}=c_{ij}}} = \begin{bmatrix} 1 \\ 2c_{ij} \\ c_{ij}^2 \\ 1 \\ 2c_{ij} \\ c_{ij}^2 \end{bmatrix}$$

and:

$$\hat{\Sigma}_{V_{Y|C_j=c_j}} = \begin{bmatrix} \hat{\Sigma}_{V_{G|C_j=c_j}} & \hat{\Sigma}_{V_{G|C_j=c_j}, V_{R|C_j=c_j}} \\ \hat{\Sigma}^T_{V_{G|C_j=c_j}, V_{R|C_j=c_j}} & \hat{\Sigma}_{V_{R|C_j=c_j}} \end{bmatrix}$$

with $\hat{\Sigma}_{V_{G|C_j=c_j}}$ and $\hat{\Sigma}_{V_{R|C_j=c_j}}$ as defined above, and:

$$\hat{\Sigma}_{V_{G|C_j=c_j}, V_{R|C_j=c_j}} = \begin{bmatrix} Cov[\hat{\sigma}^2_{\alpha_0}, \hat{\sigma}^2_{\tau_0}] & Cov[\hat{\sigma}^2_{\alpha_0}, \hat{\sigma}_{\tau_0,\tau_1}] & Cov[\hat{\sigma}^2_{\alpha_0}, \hat{\sigma}^2_{\tau_1}] \\ Cov[\hat{\sigma}_{\alpha_0,\alpha_1}, \hat{\sigma}^2_{\tau_0}] & Cov[\hat{\sigma}_{\alpha_0,\alpha_1}, \hat{\sigma}_{\tau_0,\tau_1}] & Cov[\hat{\sigma}_{\alpha_0,\alpha_1}, \hat{\sigma}^2_{\tau_1}] \\ Cov[\hat{\sigma}^2_{\alpha_1}, \hat{\sigma}^2_{\tau_0}] & Cov[\hat{\sigma}^2_{\alpha_1}, \hat{\sigma}_{\tau_0,\tau_1}] & Cov[\hat{\sigma}^2_{\alpha_1}, \hat{\sigma}^2_{\tau_1}] \end{bmatrix}$$

As previously noted, $\hat{\Sigma}_{V_{G|C_j=c_j}}$, $\hat{\Sigma}_{V_{R|C_j=c_j}}$ and $\hat{\Sigma}_{V_{G|C_j=c_j}, V_{R|C_j=c_j}}$ can be extracted from the inverse Fisher information outputted by mtg2.

Due to the large sample size available within the UK Biobank, in this analysis we split our study sample in three subgroups and ran MRNMs within each group. Therefore we estimated $V_{G_i|C_{ij}=c_{ij}}$, $V_{R_i|C_{ij}=c_{ij}}$ and $V_{Y_i|C_{ij}=c_{ij}}$ and obtained their respective SEs, over a range of $c_{ij}$ values, within each subgroup and then meta-analysed the results.

##### 1.5.4 Proportion of variability in depSympt attributable to $C_j$: estimates and standard errors

Heritability estimates for an outcome are typically present with/ after adjustment for age, sex, batch effects and PCs. To ensure the proportion of variability in depSympt attributable the considered interaction effects are comparable to heritability estimates, and other variance components estimates in the literature, we transform our interaction variance component estimates to this scale to. These estimates are presented in Table 1 and Figure 2 in the main text. Here we provide the method for obtaining these estimates from MRNM/mtg2 outputs.

Recall that for an individual $i$, the standardised residual outcome used in the random effects model of the MRNM is defined by:

$$Y_i = \frac{Y_i^o - E[Y_i^o|\underline{X}_i = \underline{x}_i, C_i^o = c_i^o]}{\sqrt{Var[Y_i^o|\underline{X}_i = \underline{x}_i, C_i^o = c_i^o]}} \sim N(0,1)$$

and:

$$Y_i = \alpha_{0i} + \alpha_{1i}C_i + \tau_{0i} + \tau_{1i}C_i$$

where:

- $Y_i^o$ is the unoriginal depSympt random variable,
- $\underline{X}_i$ ($\underline{x}_i$) is a random (observed) vector for variables contained in the fixed effects model,
- $C_i^o$ ($c_i^o$) is the unadjusted random (observed) covariate trait,
- $E[Y_i^o|\underline{X}_i = \underline{x}_i, C_i^o = c_i^o]$ is the expected value for depSympt from the fixed effects model adjusting for $\{\underline{X}_i = \underline{x}_i, C_i^o = c_i^o\}$,
- $Var[Y_i^o|\underline{X}_i = \underline{x}_i, C_i^o = c_i^o]$ is the variance of the residual random variable in the above mentioned fixed effects model,
- $C_i \sim N(0, 1)$ is the standardised residual covariate trait random variable, after fixed effects adjustment, and,
- $\{\alpha_{0i}, \alpha_{1i}, \tau_{0i}, \tau_{1i}\}$ is an individual-specific random effect corresponding to a main genetic effect, and G-C interaction effect, a main residual effect and a R-C effect respectively.

Let us change notation to:

$$E[Y_i^o|\underline{X}_i = \underline{x}_i, C_i^o = c_i^o] = \mu_{Y_i^o|\underline{X}_i=\underline{x}_i, C_i^o=c_i^o}$$

and:

$$Var[Y_i^o|\underline{X}_i = \underline{x}_i, C_i^o = c_i^o] = \sigma^2_{Y^o|\underline{X}=\underline{x}, C^o=c^o}$$

then, re-arranging the above to make $Y_i^o$ the subject of the formula gives:

$$\begin{aligned} Y_i^o &= \mu_{Y_i^o|\underline{X}_i=\underline{x}_i, C_i^o=c_i^o} + \sigma_{Y^o|\underline{X}=\underline{x}, C^o=c^o} Y_i \\ &= \mu_{Y_i^o|\underline{X}_i=\underline{x}_i, C_i^o=c_i^o} + \sigma_{Y^o|\underline{X}=\underline{x}, C^o=c^o} \big(\alpha_{0i} + \alpha_{1i}C_i + \tau_{0i} + \tau_{1i}C_i\big) \end{aligned}$$

Let the variables used within fixed effects adjustment be split into 2 groups:

$$\underline{X}_i = \begin{bmatrix} \underline{X}_{core,i} \\ \underline{X}_{other,i} \end{bmatrix}$$

where age, sex, batch effects and PCs for individual $i$ are contained within $\underline{X}_{core,i}$. Then we can write the fixed effects model as:

$$\mu_{Y_i^o|\underline{X}_i=\underline{x}_i, C_i^o=c_i^o} = \beta_0 + \underline{\beta}^T_{core}\underline{x}_{core,i} + \underline{\beta}^T_{other}\underline{x}_{other,i} + \beta_C c_i^o$$

Then:

$$Y_i^o = \beta_0 + \underline{\beta}^T_{core}\underline{x}_{core,i} + \underline{\beta}^T_{other}\underline{x}_{other,i} + \beta_C c_i^o + \sigma_{Y^o|\underline{X}=\underline{x}, C^o=c^o} \big(\alpha_{0i} + \alpha_{1i}C_i + \tau_{0i} + \tau_{1i}C_i\big)$$

Note, $Y_i^o$ here is technically $Y_i^o|\{\underline{X}_i = \underline{x}_i, C_i^o = c_i^o\}$, and therefore $Y_i^o|\{\underline{X}_{core,i} = \underline{x}_{core,i}\}$ can be approximated by:

$$\begin{aligned} Y_i^o &= \beta_0^{'} + \underline{\beta}_{core}^T \underline{x}_{core,i} + \underline{\beta}_{other}^T \underline{X}_{other,i} + \beta_C C_i^o + \sigma_{Y^o|\underline{X}=\underline{x},C^o=c^o}\big(\alpha_{0i} + \alpha_{1i}C_i + \tau_{0i} + \tau_{1i}C_i\big) \\ &\approx \beta_0^{'} + \underline{\beta}_{core}^T \underline{x}_{core,i} + \epsilon_i \end{aligned}$$

where $\beta_0^{'}$ is an updated intercept term defined as:

$$\beta_0^{'} = \beta_0 + \underline{\beta}_{other}^T E[\underline{X}_{other,i}]$$

and $\epsilon_i$ is the residual random variable from the fixed effects model for depSympt only adjusting for the core variables (age, sex, batch effects and PCs), such that:

$$\begin{aligned} \epsilon_i &\approx \underline{\beta}_{other}^T \big(\underline{X}_{other,i} - E[\underline{X}_{other,i}]\big) + \beta_C C_i^o + \sigma_{Y^o|\underline{X}=\underline{x},C^o=c^o}\big(\alpha_{0i} + \alpha_{1i}C_i + \tau_{0i} + \tau_{1i}C_i\big) \\ &\sim N(0, \sigma^2_{Y^o|\underline{X}_{core}=\underline{x}_{core}}) \end{aligned}$$

This means that:

$$\begin{aligned} &Var[Y_i^o|\underline{X}_{core,i} = \underline{x}_{core,i}] \\ &= \sigma^2_{Y^o|\underline{X}_{core}=\underline{x}_{core}} \\ &\approx Var[\underline{\beta}_{other}^T \big(\underline{X}_{other,i} - E[\underline{X}_{other,i}]\big)] + \beta_C^2 + \sigma^2_{Y^o|\underline{X}=\underline{x},C^o=c^o}\big(\sigma^2_{\alpha_0} + \sigma^2_{\alpha_1} + \sigma^2_{\tau_0} + \sigma^2_{\tau_1}\big) \end{aligned}$$

and therefore, the proportion of variability in depSympt (adjusted for age, sex, batch effects and PCs) attributable to:

- the main genetic effect $= \frac{\sigma^2_{Y^o|\underline{X}=\underline{x},C^o=c^o}}{\sigma^2_{Y^o|\underline{X}_{core}=\underline{x}_{core}}}\sigma^2_{\alpha_0}$,
- the G-C interaction effect $= \frac{\sigma^2_{Y^o|\underline{X}=\underline{x},C^o=c^o}}{\sigma^2_{Y^o|\underline{X}_{core}=\underline{x}_{core}}}\sigma^2_{\alpha_1}$,
- the main residual effect $= \frac{\sigma^2_{Y^o|\underline{X}=\underline{x},C^o=c^o}}{\sigma^2_{Y^o|\underline{X}_{core}=\underline{x}_{core}}}\sigma^2_{\tau_0}$, and,
- the R-C interaction effect $= \frac{\sigma^2_{Y^o|\underline{X}=\underline{x},C^o=c^o}}{\sigma^2_{Y^o|\underline{X}_{core}=\underline{x}_{core}}}\sigma^2_{\tau_1}$.

These are approximations, since the derivation ignores correlation between $C$ and $C^o$, but they do provide estimates on a scale more akin to those typically seen in the literature. The maths looks messy/ complex, but the routine is simple:

- store the estimate for $\sigma^2_{Y^o|\underline{X}=\underline{x},C^o=c^o}$; the variance estimate for the residual noise from the fixed effects model adjusting for $\underline{X} = \underline{x}, C^o = c^o$,
- store the estimate for $\sigma^2_{Y^o|\underline{X}_{core}=\underline{x}_{core}}$; the variance estimate for the residual noise from the fixed effects model adjusting for $\underline{X}_{core} = \underline{x}_{core}$,
- extract the required variance component estimate from the mtg2 output, and,

- input into the above equations.

Standard errors are calculated using the SE of the variance component on the scale, for example, for the G-C interaction effect:

$$SE\Big(\frac{\sigma^2_{Y^o|\underline{X}=\underline{x},C^o=c^o}}{\sigma^2_{Y^o|\underline{X}_{core}=\underline{x}_{core}}}\sigma^2_{\alpha_1}\Big) = \frac{\sigma^2_{Y^o|\underline{X}=\underline{x},C^o=c^o}}{\sigma^2_{Y^o|\underline{X}_{core}=\underline{x}_{core}}}SE\big(\sigma^2_{\alpha_1}\big)$$

where $SE\big(\sigma^2_{\alpha_1}\big)$ is part of the output from mtg2. Again, these need to be calculated within each subgroup within our analysis and then meta-analysed to obtain presented results.

### 2 Supplementary Tables

#### 2.1 Covariate traits

Table 7: Characteristics of depSympt and the covariate traits in available UK Biobank sample ($N = 119,690$). Note: all biomarkers are on their untransformed scale.

| | Missing data (%) | Mean | SD | Median |
|---|---|---|---|---|
| depSympt | 0.00 | 0.12 | 0.81 | 0.06 |
| Neuroticism | 15.27 | 3.82 | 3.14 | 3.00 |
| Childhood trauma | 1.85 | 1.21 | 1.93 | 0.00 |
| Sleep | 0.21 | 7.18 | 0.97 | 7.00 |
| BMI | 0.19 | 26.74 | 4.53 | 26.06 |
| Waist circ | 0.11 | 88.46 | 13.10 | 88.00 |
| Smoking | 32.59 | 0.19 | 0.34 | 0.00 |
| WTH ratio | 0.12 | 0.86 | 0.09 | 0.86 |
| MET total | 13.94 | 2425.87 | 2373.38 | 1693.00 |
| MET walk | 13.94 | 951.04 | 992.99 | 594.00 |
| MET mod | 13.94 | 837.75 | 1120.50 | 400.00 |
| TDI | 0.12 | -1.78 | 2.79 | -2.49 |
| MET vig | 13.94 | 637.08 | 1030.63 | 240.00 |
| LDL | 5.00 | 3.58 | 0.84 | 3.54 |
| Triglycerides | 4.88 | 1.66 | 0.97 | 1.41 |
| Vitamin D | 8.77 | 49.93 | 20.65 | 48.40 |
| CRP | 4.98 | 2.25 | 3.95 | 1.14 |
| HDL | 13.00 | 1.49 | 0.38 | 1.44 |
| MDD PRS | 0.00 | -4.31E-03 | 1.74E-04 | -4.31E-03 |
| Anorexia PRS | 0.00 | -1.02E-02 | 9.60E-05 | -1.02E-02 |
| Anxiety PRS | 0.00 | -3.83E-03 | 3.25E-04 | -3.83E-03 |
| ADHD PRS | 0.00 | -2.48E-03 | 1.88E-04 | -2.48E-03 |
| BIP PRS | 0.00 | 7.41E-04 | 1.76E-04 | 7.40E-04 |
| OCD PRS | 0.00 | -1.53E-02 | 1.57E-03 | -1.53E-02 |
| ASD PRS | 0.00 | 1.61E-04 | 1.46E-04 | 1.61E-04 |
| Scz PRS | 0.00 | -2.95E-03 | 2.18E-04 | -2.94E-03 |

Table 8: Linear fixed effects model summary.

| | % phenotypic variance explained | Beta | SE | p-value |
|---|---|---|---|---|
| Neuroticism | 21.55 | 0.46 | 2.78E-03 | 0.000E+00 |
| Childhood trauma | 5.20 | 0.23 | 2.80E-03 | 0.000E+00 |
| Sleep | 0.87 | -0.09 | 2.84E-03 | 7.927E-228 |
| BMI | 1.05 | 0.10 | 2.86E-03 | 3.162E-277 |
| Waist circ | 1.23 | 0.13 | 3.26E-03 | 0.000E+00 |
| Smoking | 1.54 | 0.13 | 3.59E-03 | 1.311E-274 |
| WTH ratio | 0.75 | 0.12 | 3.88E-03 | 3.283E-198 |
| MET total | 0.16 | -0.04 | 3.07E-03 | 9.692E-39 |
| MET walk | 0.08 | -0.03 | 3.07E-03 | 3.132E-19 |
| MET mod | 0.04 | -0.02 | 3.09E-03 | 5.054E-10 |
| TDI | 0.72 | 0.08 | 2.87E-03 | 8.502E-190 |
| MDD PRS | 0.31 | 0.05 | 2.86E-03 | 3.893E-82 |
| MET vig | 0.21 | -0.05 | 3.08E-03 | 1.119E-48 |
| Anorexia PRS | 0.07 | 0.03 | 2.91E-03 | 5.491E-20 |
| LDL | 0.00 | -0.01 | 2.94E-03 | 7.361E-02 |
| Anxiety PRS | 0.02 | 0.02 | 2.85E-03 | 3.933E-08 |
| log-Triglycerides | 0.40 | 0.06 | 3.01E-03 | 3.915E-102 |
| ADHD PRS | 0.06 | 0.02 | 2.85E-03 | 3.634E-17 |
| log-Vitamin D | 0.51 | -0.07 | 2.99E-03 | 1.554E-122 |
| log-CRP | 0.22 | 0.05 | 2.92E-03 | 1.955E-56 |
| BIP PRS | 0.02 | 0.02 | 2.89E-03 | 7.801E-08 |
| log-HDL | 0.38 | -0.07 | 3.39E-03 | 1.913E-87 |
| OCD PRS | 0.00 | 0.00 | 2.85E-03 | 7.726E-01 |
| ASD PRS | 0.01 | 0.01 | 2.85E-03 | 2.452E-05 |
| Scz PRS | 0.10 | 0.03 | 2.93E-03 | 8.415E-29 |

Output from a linear model for (standardised) depSympt against each standardised covariate in turn (main effect only). All models adjust for: sex, age, genotype batch and principal components 1-15

Only LDL and OCD polygenic risk score have a p-value $> 0.05/25$

Table 9: Comparison of the proportion of variability in depSympt explained by each covariate trait: main effects only versus fractional polynomials

| | Main effects only | | Fractional polynomials | |
|---|---|---|---|---|
| | % of variance explained | p-value | % of variance explained | p-value |
| Neuroticism | 21.55 | 0.000E+00 | 21.61 | 0.000E+00 |
| Childhood trauma | 5.20 | 0.000E+00 | 5.38 | 0.000E+00 |
| Sleep | 0.87 | 7.927E-228 | 2.24 | 0.000E+00 |
| BMI | 1.05 | 3.162E-277 | 1.25 | 1.355E-315 |
| Waist circ | 1.23 | 0.000E+00 | 1.30 | 0.000E+00 |
| Smoking | 1.54 | 1.311E-274 | 1.57 | 3.261E-275 |
| WTH ratio | 0.75 | 3.283E-198 | 0.79 | 1.207E-200 |
| MET total | 0.16 | 9.692E-39 | 0.58 | 9.833E-126 |
| MET walk | 0.08 | 3.132E-19 | 0.28 | 1.965E-62 |
| MET mod | 0.04 | 5.054E-10 | 0.30 | 2.254E-66 |
| TDI | 0.72 | 8.502E-190 | 0.74 | 4.715E-187 |
| MDD PRS | 0.31 | 3.893E-82 | 0.31 | 1.559E-79 |
| MET vig | 0.21 | 1.119E-48 | 0.68 | 5.247E-148 |
| Anorexia PRS | 0.07 | 5.491E-20 | 0.07 | 2.068E-19 |
| LDL† | 0.00 | 7.361E-02 | 0.04 | 1.868E-10 |
| Anxiety PRS | 0.02 | 3.933E-08 | 0.02 | 6.379E-08 |
| log-Triglycerides | 0.40 | 3.915E-102 | 0.49 | 2.793E-117 |
| ADHD PRS | 0.06 | 3.634E-17 | 0.06 | 1.118E-16 |
| log-Vitamin D | 0.51 | 1.554E-122 | 0.58 | 1.056E-133 |
| log-CRP | 0.22 | 1.955E-56 | 0.49 | 4.365E-117 |
| BIP PRS | 0.02 | 7.801E-08 | 0.02 | 1.239E-07 |
| log-HDL | 0.38 | 1.913E-87 | 0.47 | 1.143E-104 |
| OCD PRS‡ | 0.00 | 7.726E-01 | | |
| ASD PRS | 0.01 | 2.452E-05 | 0.01 | 3.277E-05 |
| Scz PRS | 0.10 | 8.415E-29 | 0.10 | 6.005E-28 |

Main effects only = a linear model for (standardised) depSympt against each standardised covariate in turn. Fractional polynomial = a linear model for (standardised) depSympt against each standardised covariate in turn allowing for transformations of the covariate traits using fractional polynomials (FPs). All models adjust for: sex, age, genotype batch and principal components 1-15. Note: the FP fixed effects models are not those used in the final interaction analysis, which adjust for more fixed effects.

† The effect of LDL on depSympt is non-significant in the main effects model ($p > 0.05/25$) but significant in the FP model

‡ The effect of OCD PRS on depSympt is non-significant in both models (it was excluded by the fractional polynomial model).

Note: The FP model presented in the main results allowed a non-linear relationship between age and depSympt. Here the FP model adjusted for age as a main effect only. Differences in % of depSympt variance explained for these FP models are due to this.

Table 10: Fractional polynomial fixed effects model summary.

| | % phenotypic variance explained | Beta | SE | p-value |
|---|---|---|---|---|
| **Neuroticism** | 21.61 | | | **0.00E+00** |
| I((X + 1.6)^0.5) | | 0.92 | 1.19E-02 | 0.00E+00 |
| I((X + 1.6)^3) | | 0.01 | 3.18E-04 | 4.89E-122 |
| **Childhood trauma** | 5.38 | | | **0.00E+00** |
| log((X + 1.2)) | | 0.28 | 6.95E-03 | 0.00E+00 |
| I((X + 1.2)^2) | | 0.01 | 8.75E-04 | 1.08E-45 |
| **Sleep** | 2.24 | | | **0.00E+00** |
| I(((X + 7.5)/10)^2) | | -2.10 | 4.03E-02 | 0.00E+00 |
| I(((X + 7.5)/10)^2*log(((X + 7.5)/10))) | | 6.52 | 1.46E-01 | 0.00E+00 |
| **BMI** | 1.25 | | | **1.35E-315** |
| log(((X + 3.3)/10)) | | -1.65 | 8.53E-02 | 6.93E-83 |
| I(((X + 3.3)/10)^0.5) | | 6.84 | 2.96E-01 | 1.65E-117 |
| **Waist circumference** | 1.30 | | | **0.00E+00** |
| I(((X + 3.6)/10)^1) | | -0.15 | 1.59E-01 | 3.38E-01 |
| I(((X + 3.6)/10)^2) | | 1.80 | 1.99E-01 | 1.25E-19 |
| **Smoking** | 1.57 | | | **3.26E-275** |
| I((X + 0.6)^-2) | | -0.00 | 3.92E-06 | 3.63E-06 |
| I((X + 0.6)^1) | | 0.11 | 5.07E-03 | 5.87E-106 |
| **Waist-to-hip ratio** | 0.79 | | | **1.21E-200** |
| I(((X + 4.7)/10)^3) | | 1.07 | 5.35E-02 | 1.30E-88 |
| I(((X + 4.7)/10)^3*log(((X + 4.7)/10))) | | -1.71 | 1.40E-01 | 4.22E-34 |
| **MET tot** | 0.58 | | | **9.83E-126** |
| I((X + 1.1)^-0.5) | | 0.15 | 7.02E-03 | 1.28E-94 |
| I((X + 1.1)^1) | | 0.02 | 4.35E-03 | 5.19E-08 |
| **MET walk** | 0.28 | | | **1.96E-62** |
| I((X + 1)^-2) | | 0.00 | 3.81E-05 | 2.06E-20 |
| log((X + 1)) | | -0.02 | 3.69E-03 | 6.21E-10 |
| **MET mod** | 0.30 | | | **2.25E-66** |
| I((X + 0.8)^-0.5) | | 0.06 | 4.30E-03 | 4.60E-48 |
| I((X + 0.8)^0.5) | | 0.07 | 1.11E-02 | 4.80E-09 |
| **TDI** | 0.74 | | | **4.71E-187** |
| I((X + 1.7)^1) | | 0.03 | 1.03E-02 | 7.98E-04 |
| I((X + 1.7)^2) | | 0.01 | 2.25E-03 | 4.34E-07 |
| **MDD PRS** | 0.31 | | | **1.56E-79** |
| I(((X + 4.5)/10)^1) | | 0.55 | 2.86E-02 | 3.89E-82 |
| **MET vig** | 0.68 | | | **5.25E-148** |
| log((X + 0.7)) | | -0.09 | 4.12E-03 | 7.11E-108 |

Continued on next page

**Table 10 – continued from previous page**

| | % phenotypic variance explained | Beta | SE | p-value |
|---|---|---|---|---|
| I((X + 0.7)^1) | | 0.05 | 5.18E-03 | 1.45E-19 |
| **Anorexia PRS** | 0.07 | | | **2.07E-19** |
| I(((X + 4.3)/10)^1) | | 0.27 | 2.91E-02 | 5.49E-20 |
| **LDL** | 0.04 | | | **1.87E-10** |
| I(((X + 3.3)/10)^0.5) | | -0.76 | 1.13E-01 | 1.20E-11 |
| I(((X + 3.3)/10)^0.5*log(((X + 3.3)/10))) | | 0.84 | 1.33E-01 | 2.75E-10 |
| **Anxiety PRS** | 0.02 | | | **6.38E-08** |
| I(((X + 4.2)/10)^1) | | 0.16 | 2.85E-02 | 3.93E-08 |
| **log-Triglycerides** | 0.49 | | | **2.79E-117** |
| I((X + 1.5)^-2) | | 0.00 | 4.59E-04 | 4.57E-04 |
| log((X + 1.5)) | | 0.12 | 5.16E-03 | 2.34E-121 |
| **ADHD PRS** | 0.06 | | | **1.12E-16** |
| I(((X + 4.5)/10)^1) | | 0.24 | 2.85E-02 | 3.63E-17 |
| **log-Vitamin D** | 0.58 | | | **1.06E-133** |
| I((X + 2)^1) | | -0.21 | 1.53E-02 | 2.45E-41 |
| I((X + 2)^1*log((X + 2))) | | 0.08 | 8.72E-03 | 1.62E-19 |
| **log-C-Reactive Protein** | 0.49 | | | **4.37E-117** |
| I((X + 0.6)^0.5) | | 0.34 | 1.94E-02 | 3.72E-68 |
| I((X + 0.6)^1) | | -0.07 | 7.52E-03 | 2.86E-23 |
| **BIP PRS** | 0.02 | | | **1.24E-07** |
| I(((X + 4.9)/10)^1) | | 0.16 | 2.89E-02 | 7.80E-08 |
| **log-HDL** | 0.47 | | | **1.14E-104** |
| I(((X + 3.3)/10)^1) | | -0.62 | 3.43E-02 | 1.84E-72 |
| I(((X + 3.3)/10)^1*log(((X + 3.3)/10))) | | 1.52 | 1.50E-01 | 4.26E-24 |
| **OCD PRS** | | | | |
| **ASD PRS** | 0.01 | | | **3.28E-05** |
| I(((X + 4.2)/10)^1) | | 0.12 | 2.85E-02 | 2.45E-05 |
| **Scz PRS** | 0.10 | | | **6.00E-28** |
| I(((X + 4.2)/10)^1) | | 0.33 | 2.93E-02 | 8.41E-29 |

Output from a linear model for depSympt against each covariate in turn (fractional polynomials used).
All models adjust for: sex, age, genotype batch and principal components 1-15.
Fractional polynomial transformations provided. X = standardised covariate trait.
Two p-value types presented: 1. Wald p-values corresponding to each beta estimate, and,
2. global LRT p-values (bold font) testing for inclusion of all selected FP transformations.
OCD PRS was excluded during FP modelling. All other traits had a global $p < 0.05/25$.

#### 2.2 Fractional polynomial interaction models

Table 11: Likelihood ratio test statistics and p-values comparing the full versus the null multivariate reaction norm models.

| Covariate | Test statistic (meta) | p-value (meta) |
|---|---|---|
| **Neuroticism** | 658.69 | 5.06E-139 |
| **Childhood trauma** | 283.66 | 2.59E-58 |
| **Sleep** | 204.63 | 1.97E-41 |
| **BMI** | 93.26 | 6.36E-18 |
| **Waist circ** | 83.70 | 6.15E-16 |
| **Smoking** | 56.34 | 2.49E-10 |
| **WTH ratio** | 50.10 | 4.49E-09 |
| **MET total** | 41.90 | 1.92E-07 |
| **MET walk** | 32.83 | 1.13E-05 |
| **MET mod** | 23.78 | 5.73E-04 |
| **TDI** | 20.84 | 1.96E-03 |
| MDD PRS | 20.12 | 2.63E-03 |
| MET vig | 18.09 | 6.00E-03 |
| Anorexia PRS | 14.72 | 2.26E-02 |
| LDL | 12.57 | 5.04E-02 |
| Anxiety PRS | 12.08 | 6.01E-02 |
| log-Triglycerides | 11.32 | 7.90E-02 |
| ADHD PRS | 8.76 | 1.88E-01 |
| log-Vitamin D | 5.84 | 4.42E-01 |
| log-CRP | 4.69 | 5.85E-01 |
| BIP PRS | 4.20 | 6.50E-01 |
| log-HDL | 3.52 | 7.42E-01 |
| OCD PRS | 2.87 | 8.25E-01 |
| ASD PRS | 2.67 | 8.49E-01 |
| Scz PRS | 1.33 | 9.70E-01 |

full model = both G-C and R-C interactions.
null model = no interactions.
Results are ordered by p-value.
Significance is set at $\alpha = 0.05/25 = 0.002$.
Significance results highlighted in bold.

Table 12: LRT statistics and p-values: all covariates, all subgroups

| | Test statistic | | | | p-value | | | |
|---|---|---|---|---|---|---|---|---|
| | Subgroup 1 | Subgroup 2 | Subgroup 3 | Meta | Subgroup 1 | Subgroup 2 | Subgroup 3 | Meta |
| Neu | 243.85 | 220.81 | 247.27 | 658.69 | 8.46E-50 | 6.99E-45 | 1.57E-50 | 5.06E-139 |
| Childhood trauma | 125.13 | 123.41 | 78.93 | 283.66 | 1.36E-24 | 3.13E-24 | 5.94E-15 | 2.59E-58 |
| Sleep | 73.51 | 75.32 | 96.36 | 204.63 | 7.77E-14 | 3.29E-14 | 1.44E-18 | 1.97E-41 |
| BMI | 44.99 | 32.50 | 48.56 | 93.26 | 4.71E-08 | 1.31E-05 | 9.11E-09 | 6.36E-18 |
| Waist circ | 40.47 | 31.14 | 43.93 | 83.70 | 3.68E-07 | 2.38E-05 | 7.64E-08 | 6.15E-16 |
| Smoking | 17.24 | 57.04 | 6.34 | 56.34 | 8.45E-03 | 1.79E-10 | 3.86E-01 | 2.49E-10 |
| WTH atio | 21.09 | 17.93 | 37.92 | 50.10 | 1.77E-03 | 6.41E-03 | 1.17E-06 | 4.49E-09 |
| MET total | 12.20 | 18.42 | 36.11 | 41.90 | 5.76E-02 | 5.27E-03 | 2.62E-06 | 1.92E-07 |
| MET walk | 9.17 | 13.73 | 32.38 | 32.83 | 1.64E-01 | 3.28E-02 | 1.38E-05 | 1.13E-05 |
| MET mod | 9.69 | 11.21 | 23.65 | 23.78 | 1.38E-01 | 8.21E-02 | 6.05E-04 | 5.73E-04 |
| TDI | 19.34 | 9.65 | 12.16 | 20.84 | 3.62E-03 | 1.40E-01 | 5.85E-02 | 1.96E-03 |
| MDD PRS | 17.28 | 9.40 | 13.65 | 20.12 | 8.30E-03 | 1.52E-01 | 3.38E-02 | 2.63E-03 |
| MET vig | 7.03 | 14.65 | 15.62 | 18.09 | 3.18E-01 | 2.32E-02 | 1.60E-02 | 6.00E-03 |
| Anor. PRS | 6.67 | 12.07 | 13.97 | 14.72 | 3.53E-01 | 6.04E-02 | 2.99E-02 | 2.26E-02 |
| LDL | 18.37 | 5.81 | 3.21 | 12.57 | 5.37E-03 | 4.44E-01 | 7.82E-01 | 5.04E-02 |
| Anx. PRS | 13.36 | 5.09 | 10.14 | 12.08 | 3.76E-02 | 5.32E-01 | 1.19E-01 | 6.01E-02 |
| log-Tri | 16.73 | 2.76 | 6.19 | 11.32 | 1.03E-02 | 8.39E-01 | 4.02E-01 | 7.90E-02 |
| ADHD PRS | 8.42 | 9.08 | 6.65 | 8.76 | 2.09E-01 | 1.69E-01 | 3.54E-01 | 1.88E-01 |
| log-Vit D | 9.07 | 5.07 | 4.61 | 5.84 | 1.70E-01 | 5.35E-01 | 5.94E-01 | 4.42E-01 |
| log-CRP | 5.50 | 6.09 | 5.49 | 4.69 | 4.82E-01 | 4.13E-01 | 4.83E-01 | 5.85E-01 |
| BIP PRS | 6.20 | 4.93 | 4.93 | 4.20 | 4.01E-01 | 5.53E-01 | 5.52E-01 | 6.50E-01 |
| log-HDL | 4.33 | 7.11 | 2.44 | 3.52 | 6.33E-01 | 3.11E-01 | 8.75E-01 | 7.42E-01 |
| OCD PRS | 3.64 | 3.88 | 5.56 | 2.87 | 7.26E-01 | 6.93E-01 | 4.74E-01 | 8.25E-01 |
| ASD PRS | 3.51 | 6.37 | 1.92 | 2.67 | 7.43E-01 | 3.83E-01 | 9.27E-01 | 8.49E-01 |
| Scz PRS | 1.64 | 4.50 | 2.31 | 1.33 | 9.49E-01 | 6.09E-01 | 8.89E-01 | 9.70E-01 |

Meta p-values and test statistics were calculated using Fishers method. See SM Section 1.5 for details.

Table 13: Likelihood ratio test statistics and p-values when comparing models with and without interactions, using untransformed depSympt and RINT depSympt (sensitivity analysis).

| | Untransformed depSympt | | RINT depSympt | |
|---|---|---|---|---|
| | Test statistic | p-value | Test statistic | p-value |
| **Neuroticism** | 658.69 | 5.06E-139 | 740.15 | 1.31E-156 |
| **Childhood trauma** | 283.66 | 2.59E-58 | 343.97 | 3.03E-71 |
| **Sleep** | 204.63 | 1.97E-41 | 228.33 | 1.74E-46 |
| **BMI** | 93.26 | 6.36E-18 | 123.71 | 2.71E-24 |
| **Waist circ** | 83.70 | 6.15E-16 | 101.84 | 1.03E-19 |
| **Smoking** | 56.34 | 2.49E-10 | 66.63 | 2.00E-12 |
| **WTH ratio** | 50.10 | 4.49E-09 | 61.82 | 1.92E-11 |
| **MET total** | 41.90 | 1.92E-07 | 36.04 | 2.71E-06 |
| **MET walk** | 32.83 | 1.13E-05 | 33.57 | 8.16E-06 |
| **MET mod** | 23.78 | 5.73E-04 | 21.13 | 1.74E-03 |
| **TDI** | 20.84 | 1.96E-03 | 27.35 | 1.25E-04 |
| MDD PRS | 20.12 | 2.63E-03 | 23.07 | 7.72E-04 |
| MET vig | 18.09 | 6.00E-03 | 14.88 | 2.12E-02 |
| Anorexia PRS | 14.72 | 2.26E-02 | 13.93 | 3.04E-02 |
| LDL | 12.57 | 5.04E-02 | 10.41 | 1.08E-01 |
| Anxiety PRS | 12.08 | 6.01E-02 | 10.73 | 9.72E-02 |
| log-Tri | 11.32 | 7.90E-02 | 13.69 | 3.33E-02 |
| ADHD PRS | 8.76 | 1.88E-01 | 9.51 | 1.47E-01 |
| log-Vitamin D | 5.84 | 4.42E-01 | 7.61 | 2.68E-01 |
| log-CRP | 4.69 | 5.85E-01 | 4.61 | 5.95E-01 |
| BIP PRS | 4.20 | 6.50E-01 | 4.67 | 5.87E-01 |
| log-HDL | 3.52 | 7.42E-01 | 2.64 | 8.53E-01 |
| OCD PRS | 2.87 | 8.25E-01 | 2.54 | 8.63E-01 |
| ASD PRS | 2.67 | 8.49E-01 | 3.37 | 7.61E-01 |
| Scz PRS | 1.33 | 9.70E-01 | 1.55 | 9.56E-01 |

RINT = rank-based inverse normal transformation. Covariates highlighted in bold had p-values $< 0.05/25$ when using untransformed depSympt as the outcome. Covariates above the line had p-values $< 0.05/25$ when using RINT depSympt as the outcome. Differences are minor including the PRS for MDD being significant when using RINT depSympt, but not for untransformed depSympt.

Table 14: Percentage of variability in depSympt attributable to: 1. the fixed effects, 2. the G-C interaction and 3. the R-C interaction, for each covariate trait.

| Covariate | Fixed effect | G-C interaction | | R-C interaction | |
|---|---|---|---|---|---|
| | | Estimate | 95% CI | Estimate | 95% CI |
| Neuroticism | 15.15 | -0.15 | [-0.76, 0.46] | 2.58 | [ 1.86, 3.30] |
| Childhood trauma | 1.51 | 0.59 | [-0.14, 1.32] | 2.98 | [ 2.18, 3.77] |
| Sleep | 1.17 | 1.22 | [ 0.54, 1.89] | 2.52 | [ 1.78, 3.27] |
| BMI | 0.37 | -0.23 | [-0.86, 0.41] | 1.39 | [ 0.68, 2.09] |
| Waist circ | 0.40 | -0.15 | [-0.78, 0.48] | 1.48 | [ 0.78, 2.19] |
| Smoking | 0.18 | 0.47 | [-0.52, 1.46] | 1.57 | [ 0.51, 2.63] |
| WTH ratio | 0.23 | -0.33 | [-0.95, 0.29] | 1.03 | [ 0.34, 1.73] |
| MET tot | 0.32 | 0.23 | [-0.42, 0.87] | 0.53 | [-0.17, 1.24] |
| MET walk | 0.16 | 0.10 | [-0.55, 0.74] | 1.18 | [ 0.45, 1.92] |
| MET mod | 0.15 | -0.26 | [-0.87, 0.35] | -0.08 | [-0.78, 0.61] |
| TDI | 0.05 | -0.19 | [-0.81, 0.42] | 1.67 | [ 0.97, 2.38] |
| MDD PRS | 0.12 | -0.30 | [-0.88, 0.29] | 0.16 | [-0.51, 0.83] |
| MET vig | 0.35 | 0.51 | [-0.16, 1.17] | 0.26 | [-0.44, 0.95] |
| Anorexia PRS | 0.05 | 0.01 | [-0.58, 0.59] | -0.01 | [-0.68, 0.66] |
| LDL | 0.01 | -0.23 | [-0.89, 0.44] | 0.89 | [ 0.14, 1.65] |
| Anxiety PRS | 0.01 | -0.55 | [-1.15, 0.04] | 0.92 | [ 0.23, 1.60] |
| log-Tri | 0.08 | 0.45 | [-0.22, 1.12] | 0.38 | [-0.37, 1.13] |
| ADHD PRS | 0.00 | -0.43 | [-1.03, 0.17] | 0.33 | [-0.35, 1.01] |
| log-Vitamin D | 0.10 | -0.17 | [-0.83, 0.49] | 1.11 | [ 0.35, 1.87] |
| log-CRP | 0.00 | 0.85 | [ 0.19, 1.51] | -0.63 | [-1.35, 0.10] |
| BIP PRS | 0.01 | -0.43 | [-1.01, 0.14] | 0.52 | [-0.14, 1.18] |
| log-HDL | 0.00 | -0.62 | [-1.33, 0.08] | 1.05 | [ 0.26, 1.84] |
| OCD PRS | -0.00 | 0.20 | [-0.39, 0.80] | -0.51 | [-1.18, 0.16] |
| ASD PRS | 0.01 | 0.35 | [-0.25, 0.94] | -0.47 | [-1.15, 0.21] |
| Scz PRS | 0.02 | -0.00 | [-0.59, 0.59] | -0.14 | [-0.81, 0.54] |

FPs used in the fixed effects model. Table is ordered by p-value.

Please see Table 11 for p-values.

Table 15: Percentage of variability in *residual* depSympt attributable to genotype-covariate (G-C) and residual-covariate (R-C) interactions.

| | G-C interaction | | R-C interaction | |
|---|---|---|---|---|
| Covariate | Estimate | 95% CI | Estimate | 95% CI |
| Neuroticism | -0.21 | [-1.06, 0.64] | 3.61 | [ 2.60, 4.61] |
| Childhood trauma | 0.68 | [-0.16, 1.52] | 3.43 | [ 2.51, 4.35] |
| Sleep | 1.41 | [ 0.63, 2.19] | 2.92 | [ 2.05, 3.78] |
| BMI | -0.26 | [-1.00, 0.47] | 1.60 | [ 0.79, 2.42] |
| Waist circ | -0.17 | [-0.90, 0.56] | 1.72 | [ 0.90, 2.53] |
| Smoking | 0.55 | [-0.61, 1.71] | 1.84 | [ 0.59, 3.08] |
| WTH ratio | -0.38 | [-1.10, 0.33] | 1.19 | [ 0.39, 1.99] |
| MET total | 0.26 | [-0.49, 1.01] | 0.61 | [-0.20, 1.43] |
| MET walk | 0.11 | [-0.63, 0.85] | 1.37 | [ 0.51, 2.22] |
| MET mod | -0.30 | [-1.00, 0.40] | -0.10 | [-0.89, 0.70] |
| TDI | -0.23 | [-0.94, 0.49] | 1.93 | [ 1.12, 2.75] |
| MDD PRS | -0.35 | [-1.02, 0.33] | 0.18 | [-0.59, 0.96] |
| MET vig | 0.58 | [-0.18, 1.35] | 0.30 | [-0.51, 1.10] |
| Anorexia PRS | 0.01 | [-0.67, 0.68] | -0.01 | [-0.79, 0.76] |
| LDL | -0.26 | [-1.03, 0.50] | 1.03 | [ 0.16, 1.90] |
| Anxiety PRS | -0.64 | [-1.32, 0.05] | 1.06 | [ 0.27, 1.85] |
| log-Tri | 0.52 | [-0.26, 1.30] | 0.44 | [-0.43, 1.31] |
| ADHD PRS | -0.50 | [-1.19, 0.19] | 0.39 | [-0.40, 1.17] |
| log-Vitamin D | -0.20 | [-0.96, 0.57] | 1.28 | [ 0.41, 2.16] |
| log-CRP | 0.99 | [ 0.22, 1.75] | -0.72 | [-1.56, 0.11] |
| BIP PRS | -0.51 | [-1.17, 0.16] | 0.60 | [-0.17, 1.37] |
| log-HDL | -0.72 | [-1.54, 0.10] | 1.22 | [ 0.30, 2.13] |
| OCD PRS | 0.23 | [-0.45, 0.92] | -0.59 | [-1.37, 0.18] |
| ASD PRS | 0.40 | [-0.29, 1.09] | -0.54 | [-1.32, 0.24] |
| Scz PRS | -0.00 | [-0.69, 0.68] | -0.16 | [-0.94, 0.62] |

FPs used in the fixed effects model. Table is ordered by p-value.
Please see Table 11 for p-values.

### 3 Supplementary Figures

#### 3.1 Covariate traits distributions

##### 3.1.1 Covariates in Analysis group 1

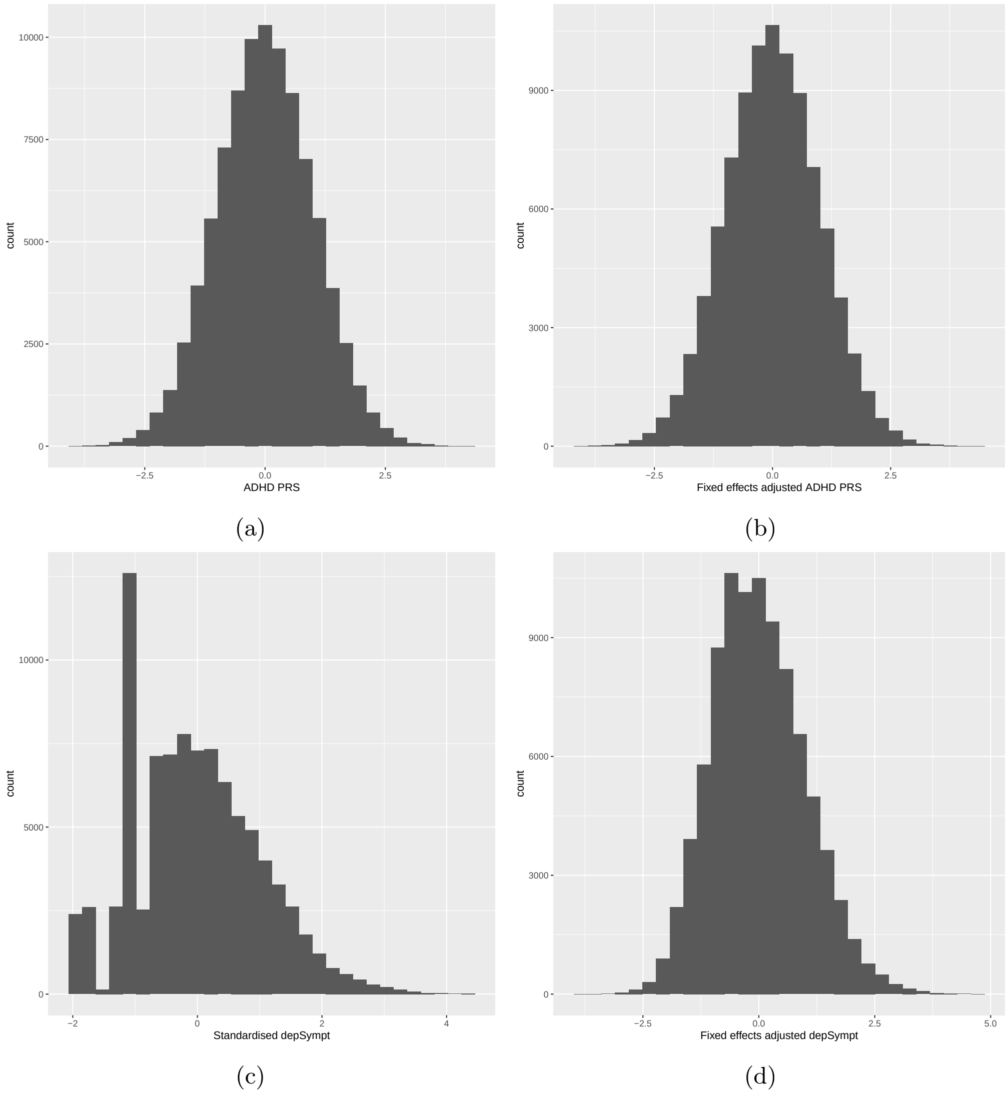

Figure 3: Histogram of standardised: (a) ADHD PRS, (b) ADHD PRS post fixed effects adjustment, (c) depSympt (Analysis group 1) and (d) depSympt post fixed effects adjustment (for use in interaction analysis with ADHD PRS), in the available UK Biobank study population.

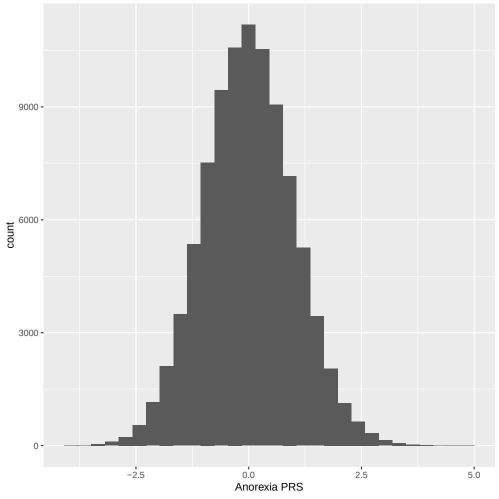

(a)

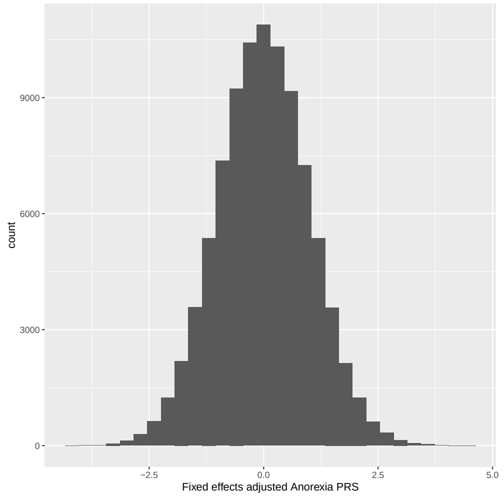

(b)

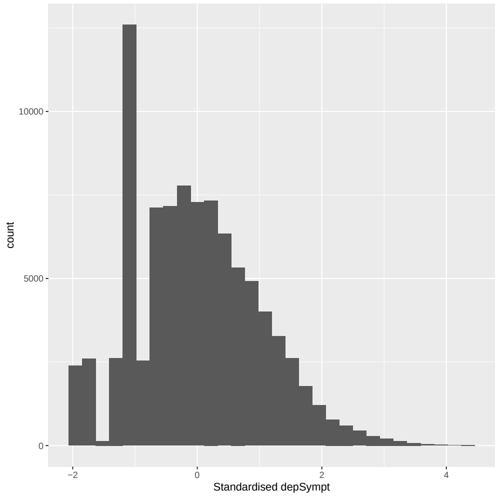

(c)

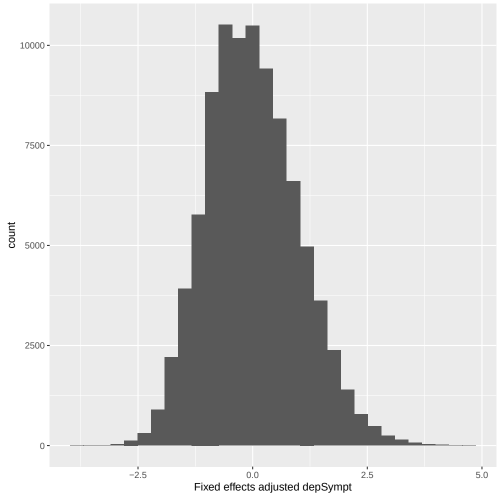

(d)

Figure 4: Histogram of standardised: (a) anorexia PRS, (b) anorexia PRS post fixed effects adjustment, (c) depSympt (Analysis group 1) and (d) depSympt post fixed effects adjustment (for use in interaction analysis with anorexia PRS), in the available UK Biobank study population.

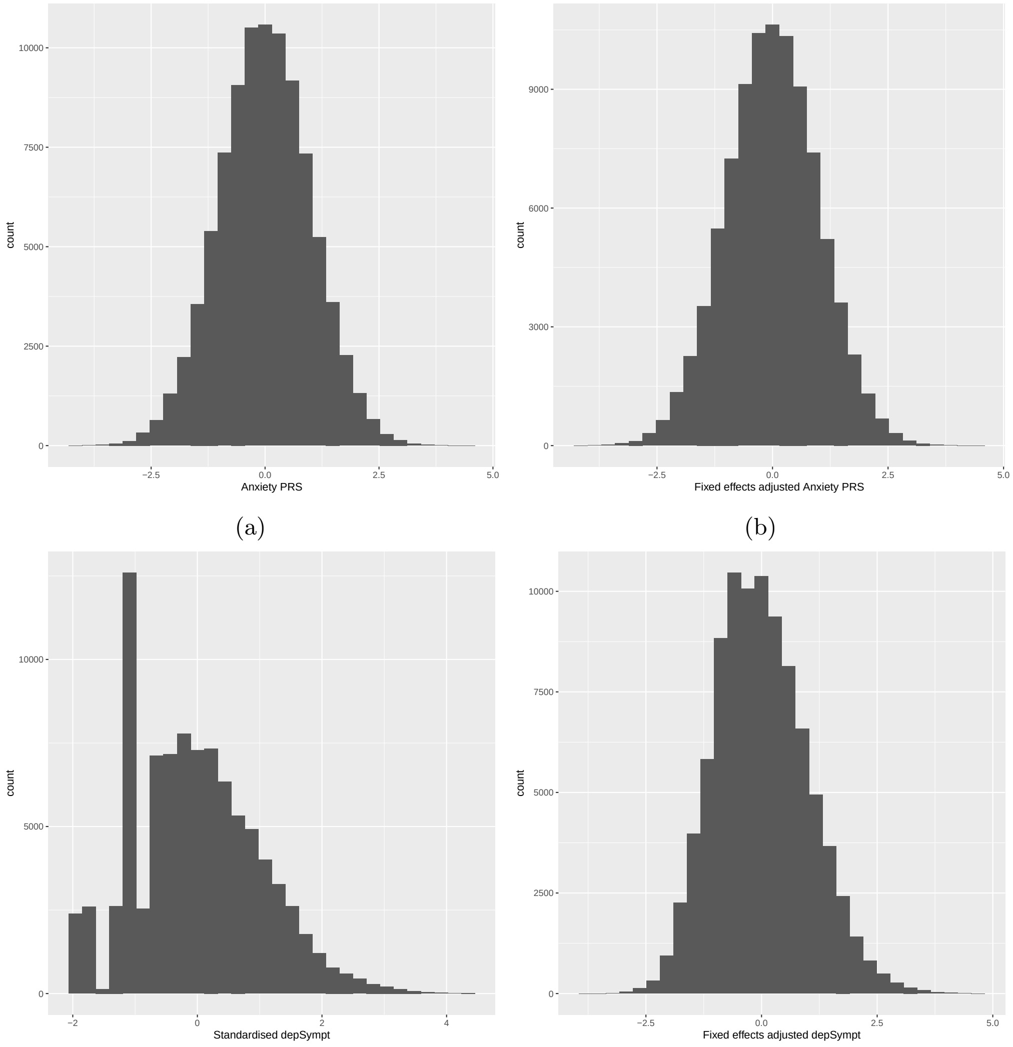

(a) (b) (c) (d)

Figure 5: Histogram of standardised: (a) anxiety PRS, (b) anxiety PRS post fixed effects adjustment, (c) depSympt (Analysis group 1) and (d) depSympt post fixed effects adjustment (for use in interaction analysis with anxiety PRS), in the available UK Biobank study population.

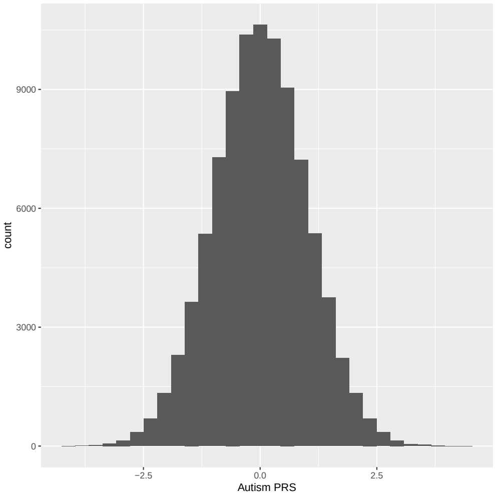

(a)

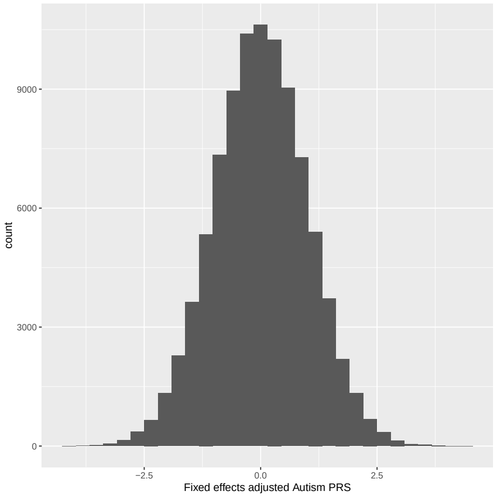

(b)

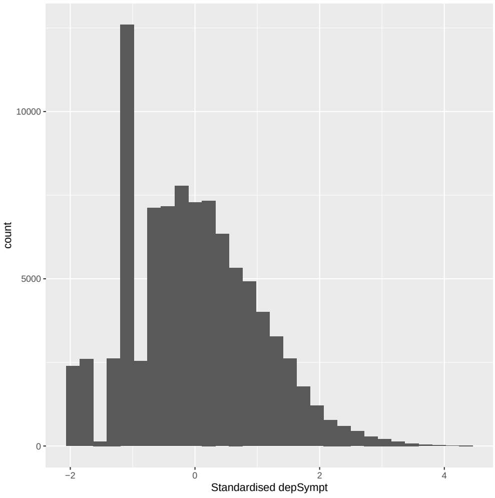

(c)

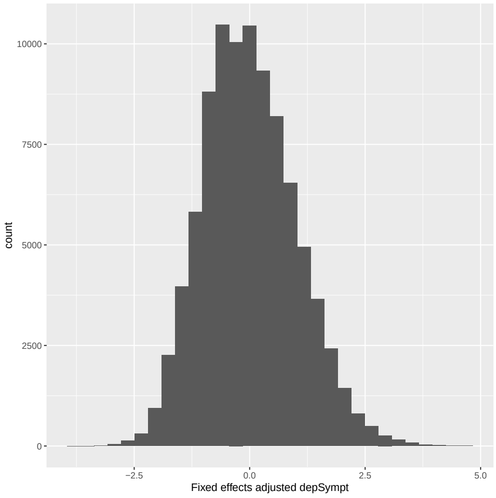

(d)

Figure 6: Histogram of standardised: (a) autism (ASD) PRS, (b) autism PRS post fixed effects adjustment, (c) depSympt (Analysis group 1) and (d) depSympt post fixed effects adjustment (for use in interaction analysis with autism PRS), in the available UK Biobank study population.

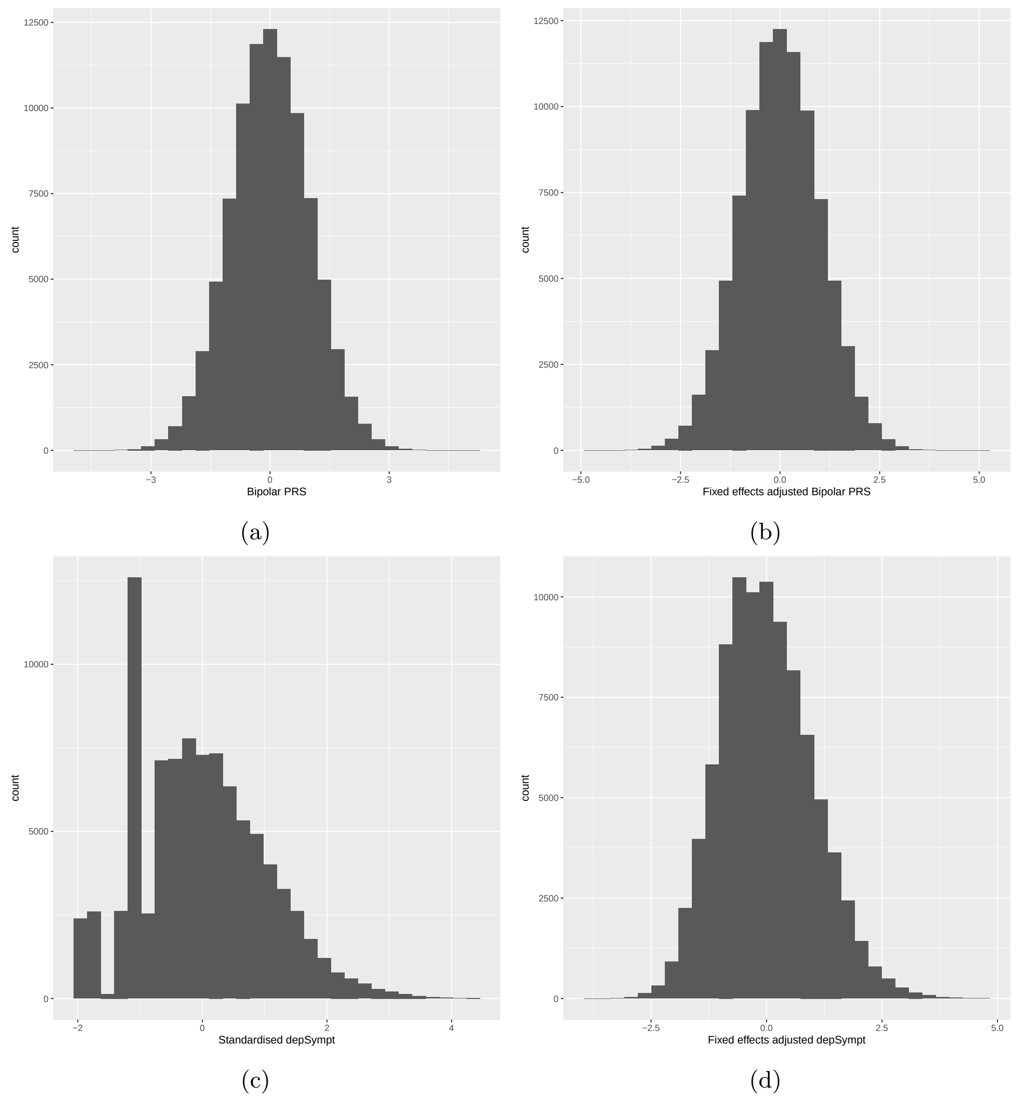

Figure 7: Histogram of standardised: (a) bipolar disorder PRS, (b) bipolar disorder PRS post fixed effects adjustment, (c) depSympt (Analysis group 1) and (d) depSympt post fixed effects adjustment (for use in interaction analysis with bipolar disorder PRS), in the available UK Biobank study population.

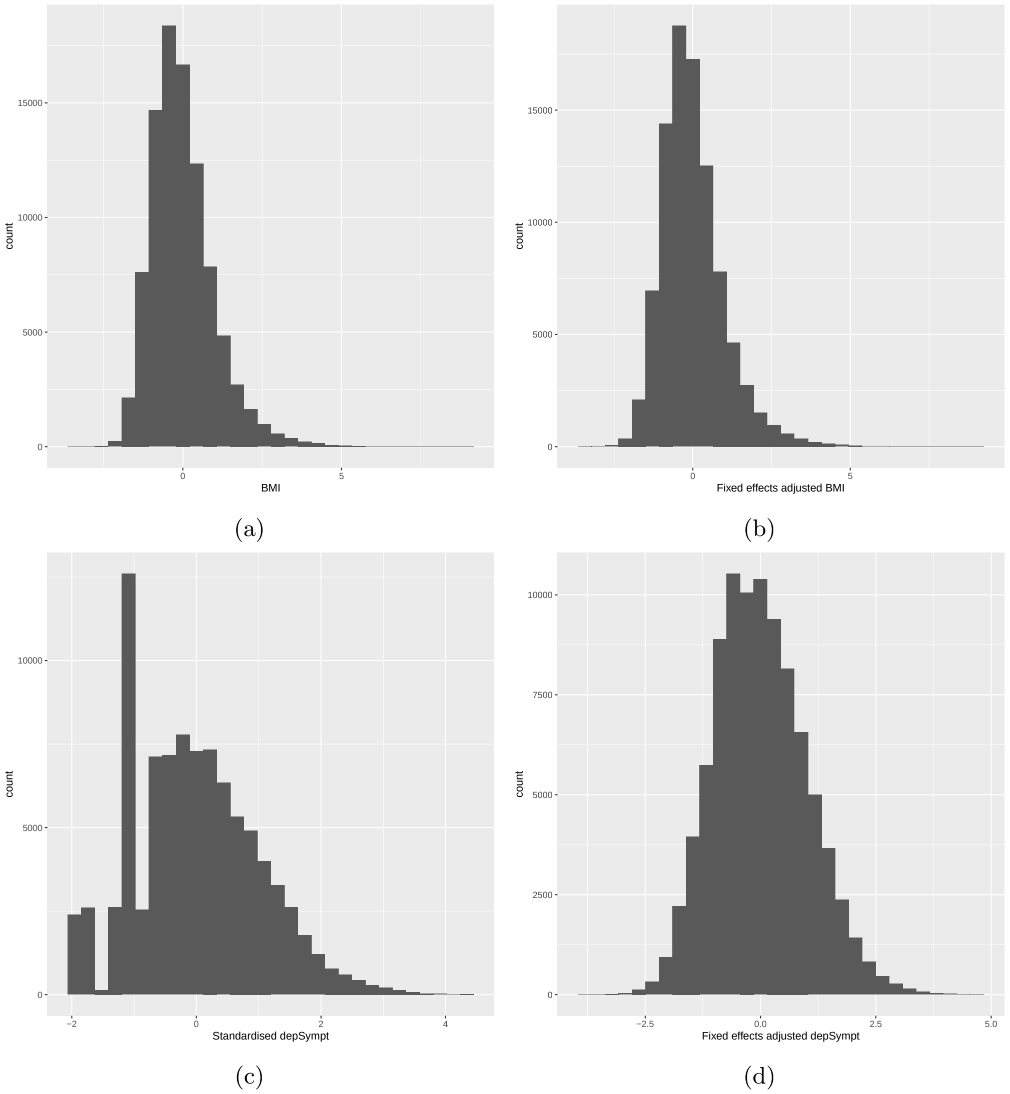

(a) (b) (c) (d)

Figure 8: Histogram of standardised: (a) BMI, (b) BMI post fixed effects adjustment, (c) depSympt (Analysis group 1) and (d) depSympt post fixed effects adjustment (for use in interaction analysis with BMI), in the available UK Biobank study population

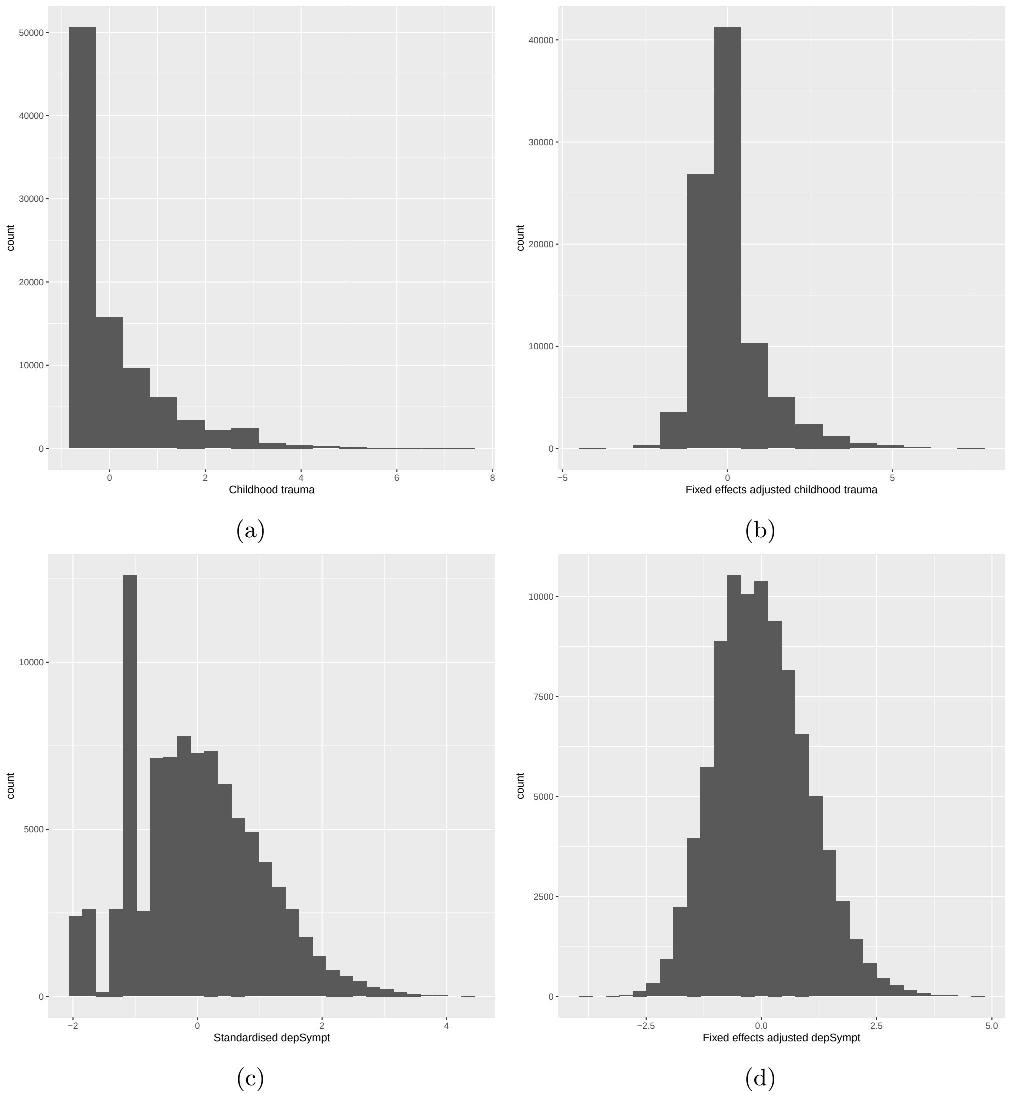

Figure 9: Histogram of standardised: (a) childhood trauma summary variable, (b) childhood trauma summary variable post fixed effects adjustment, (c) depSympt (Analysis group 1) and (d) depSympt post fixed effects adjustment (for use in interaction analysis with childhood trauma), in the available UK Biobank study population.

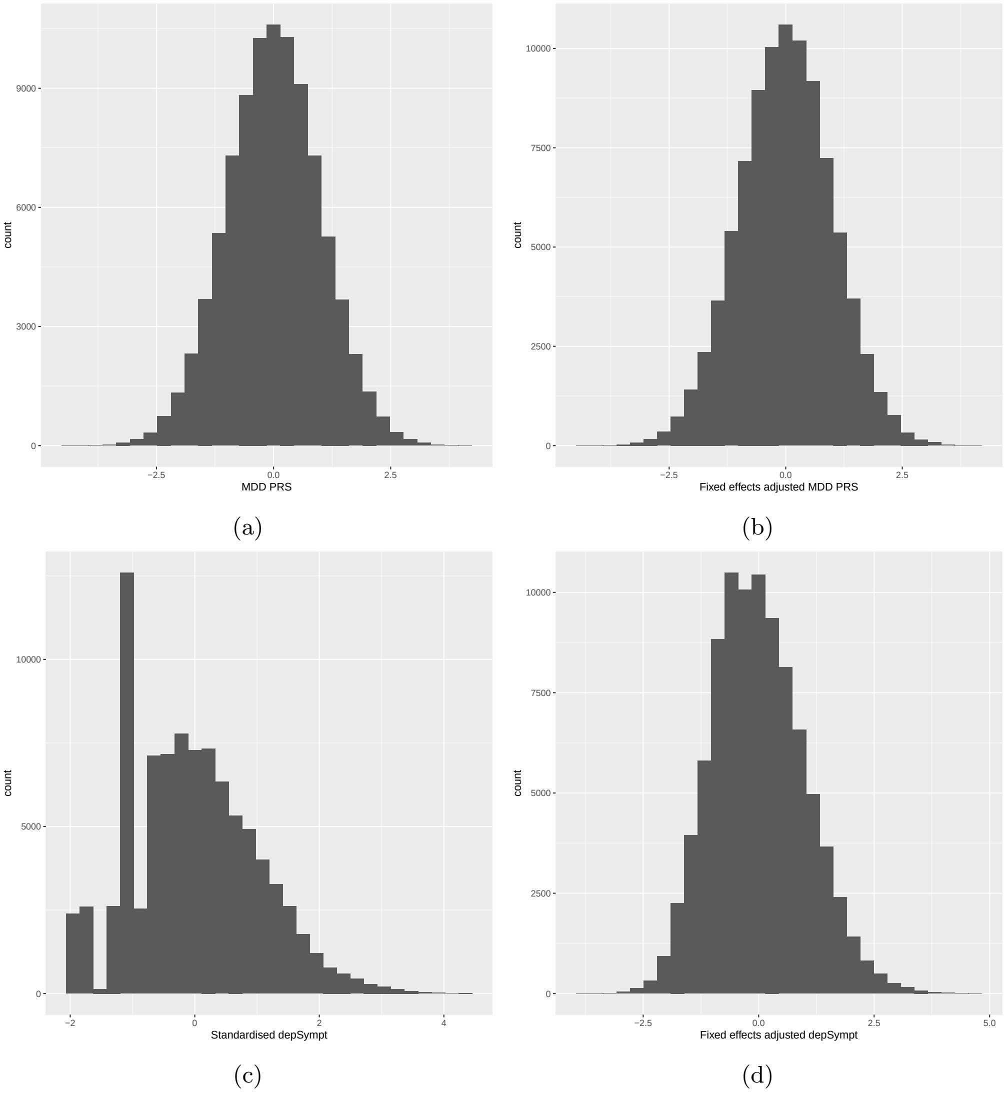

Figure 10: Histogram of standardised: (a) MDD PRS, (b) MDD PRS post fixed effects adjustment, (c) depSympt (Analysis group 1) and (d) depSympt post fixed effects adjustment (for use in interaction analysis with MDD PRS), in the available UK Biobank study population.

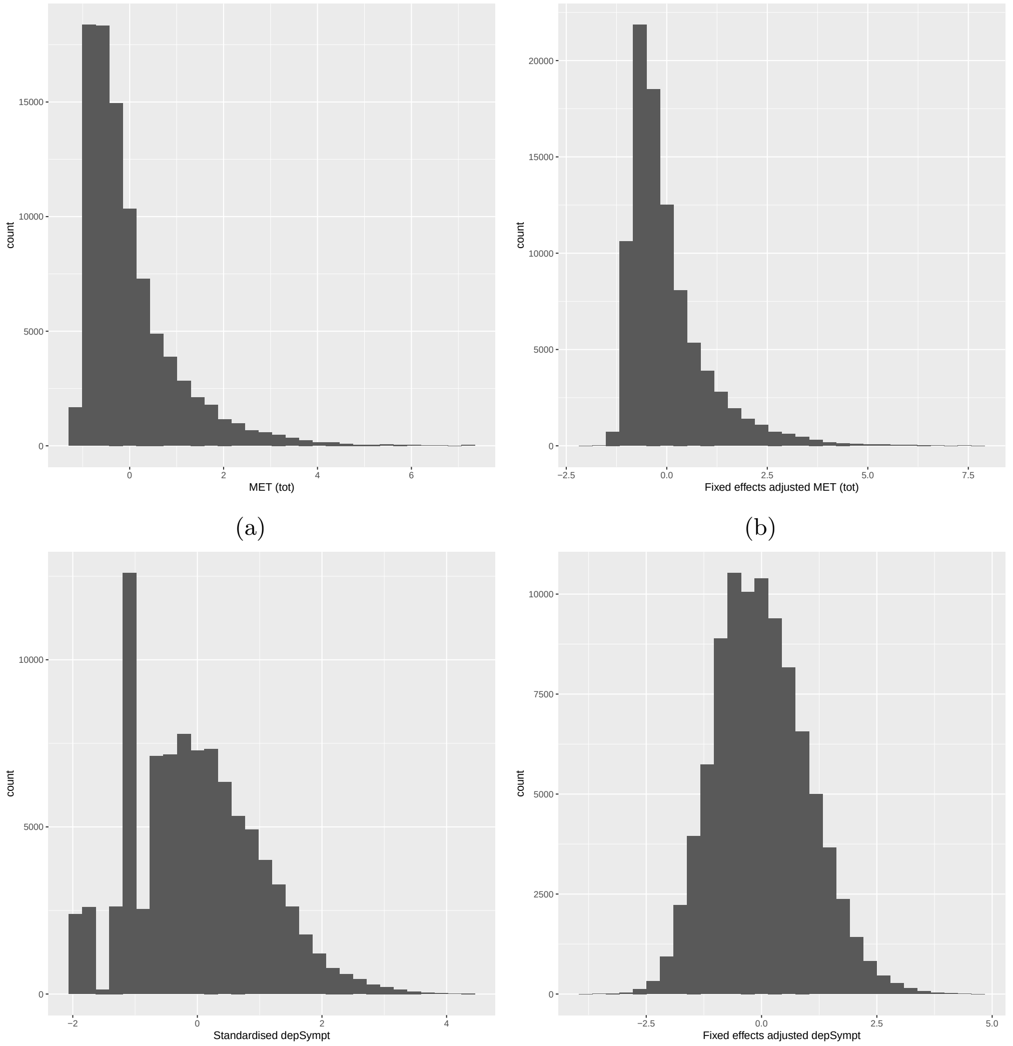

(a) (b) (c) (d)

Figure 11: Histogram of standardised: (a) MET total, (b) MET total post fixed effects adjustment, (c) depSympt (Analysis group 1) and (d) depSympt post fixed effects adjustment (for use in interaction analysis with MET total), in the available UK Biobank study population.

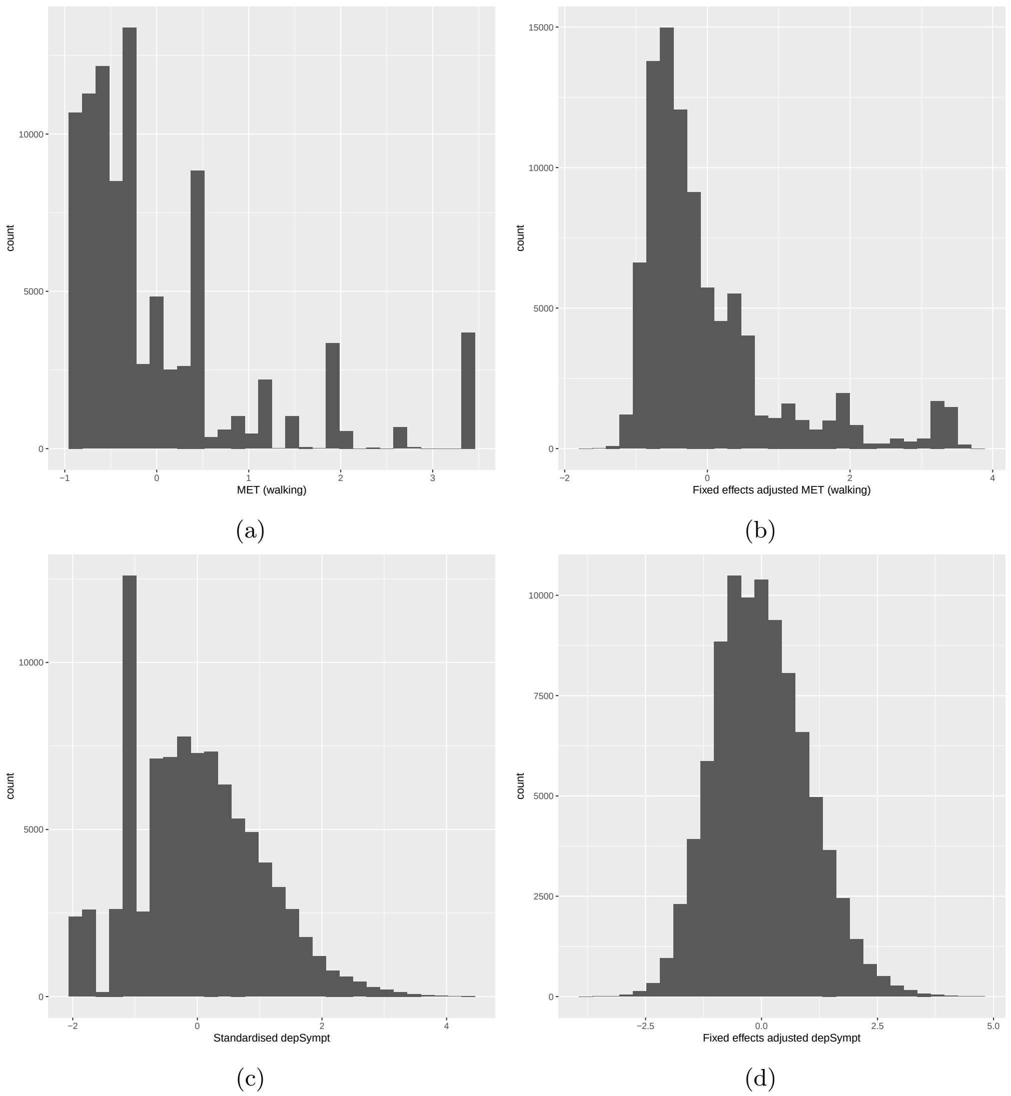

(a) (b) (c) (d)

Figure 12: Histogram of standardised: (a) MET walk, (b) MET walk post fixed effects adjustment, (c) depSympt (Analysis group 1) and (d) depSympt post fixed effects adjustment (for use in interaction analysis with MET walk), in the available UK Biobank study population.

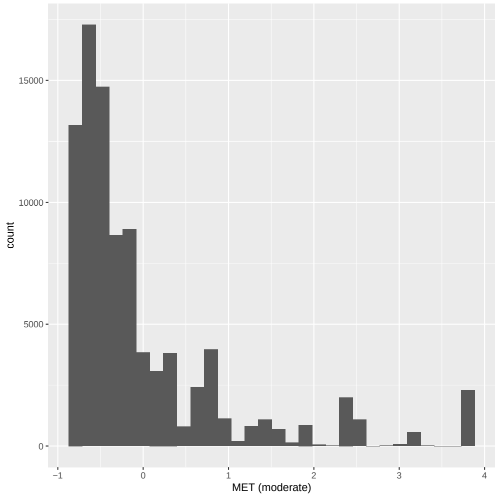

(a)

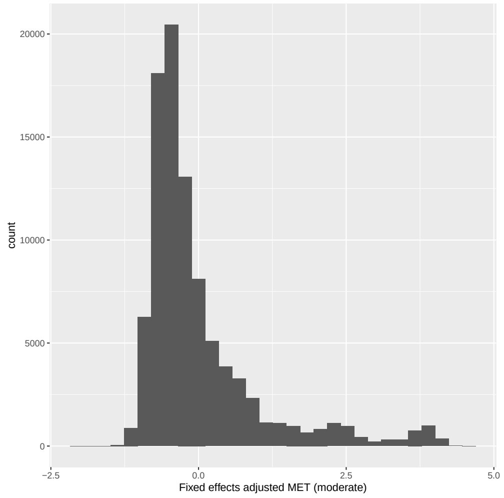

(b)

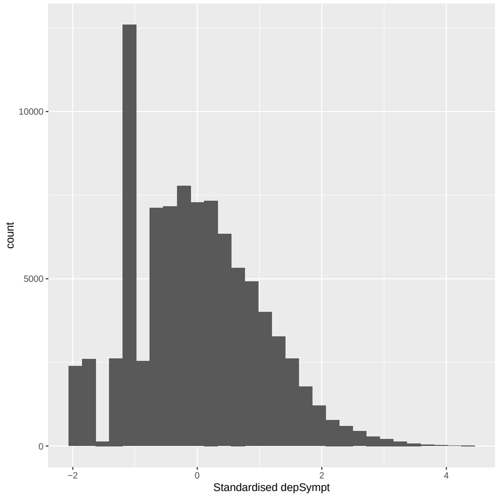

(c)

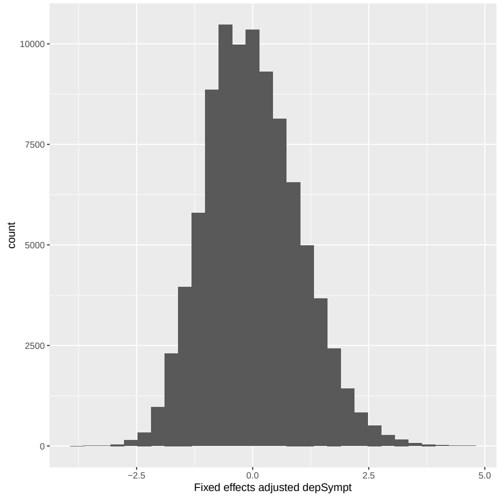

(d)

Figure 13: Histogram of standardised: (a) MET moderate, (b) MET moderate post fixed effects adjustment, (c) depSympt (Analysis group 1) and (d) depSympt post fixed effects adjustment (for use in interaction analysis with MET moderate), in the available UK Biobank study population.

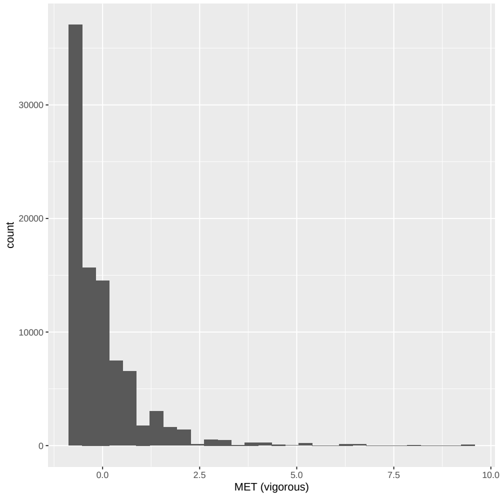

(a)

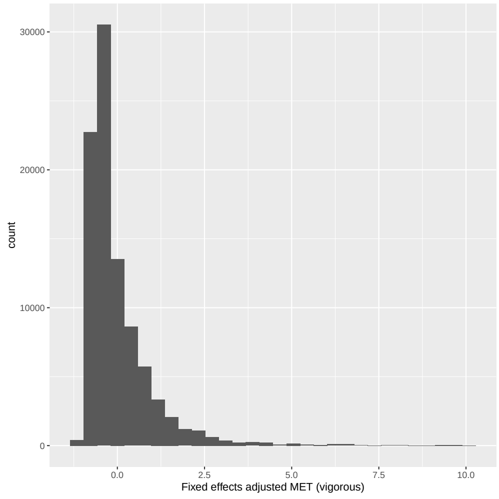

(b)

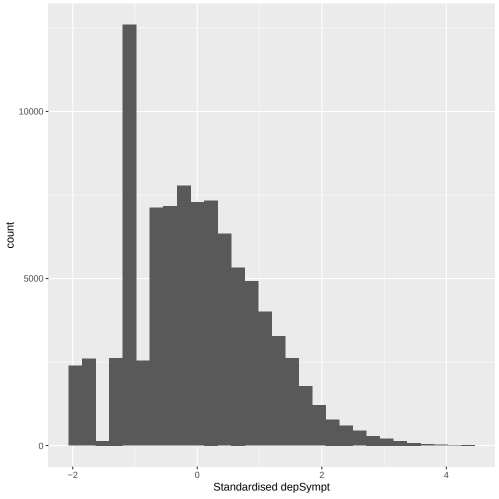

(c)

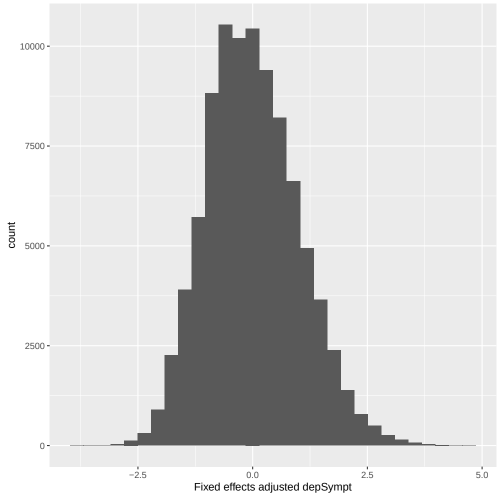

(d)

Figure 14: Histogram of standardised: (a) MET vigorous, (b) MET vigorous post fixed effects adjustment, (c) depSympt (Analysis group 1) and (d) depSympt post fixed effects adjustment (for use in interaction analysis with MET vigorous), in the available UK Biobank study population.

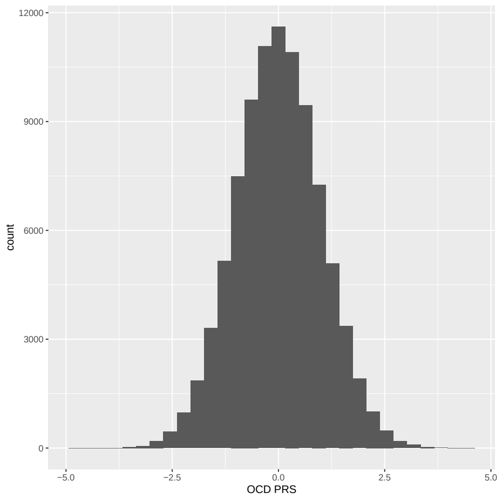

(a)

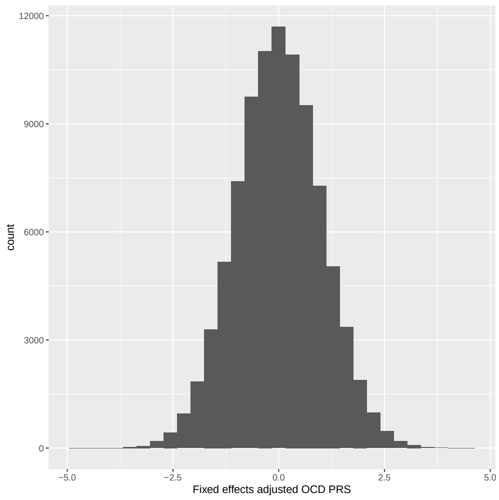

(b)

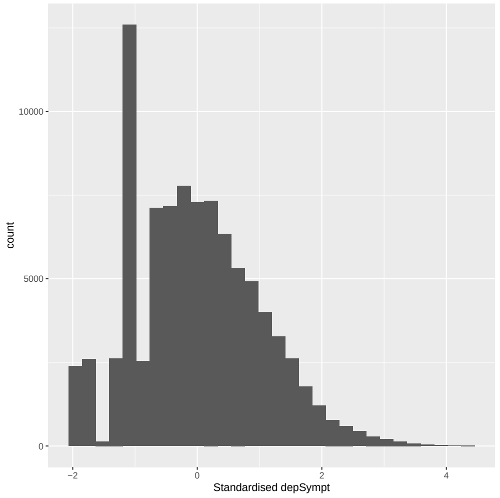

(c)

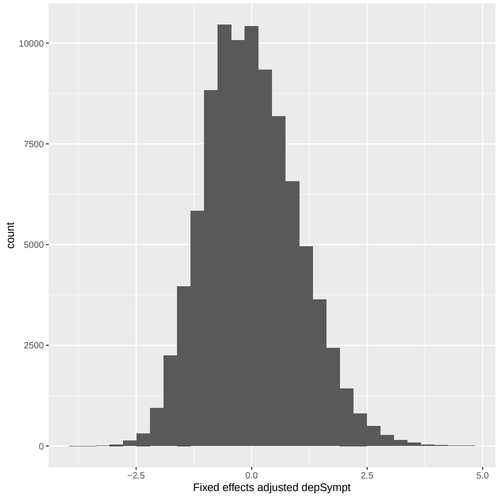

(d)

Figure 15: Histogram of standardised: (a) OCD PRS, (b) OCD PRS post fixed effects adjustment, (c) depSympt (Analysis group 1) and (d) depSympt post fixed effects adjustment (for use in interaction analysis with OCD PRS), in the available UK Biobank study population.

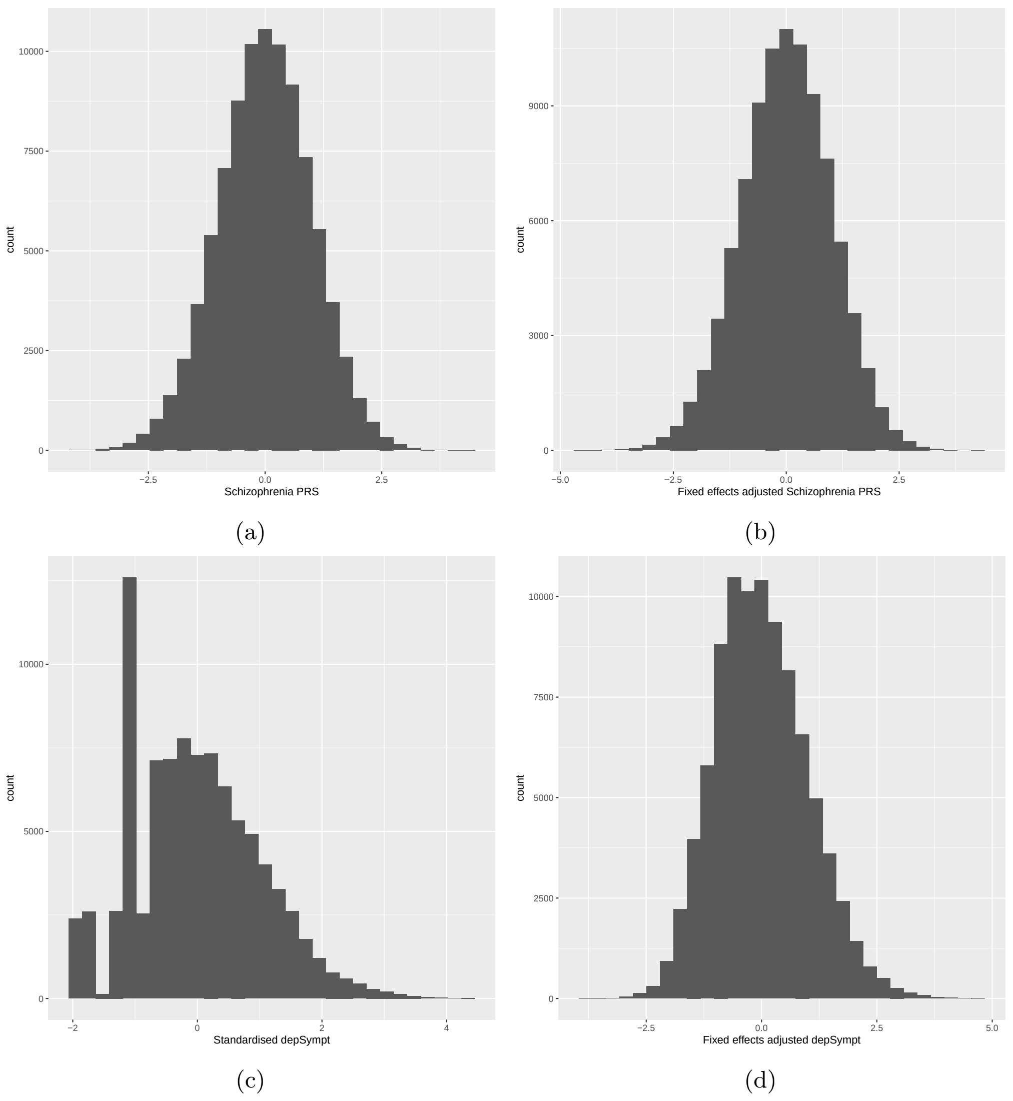

Figure 16: Histogram of standardised: (a) schizophrenia PRS, (b) schizophrenia PRS post fixed effects adjustment, (c) depSympt (Analysis group 1) and (d) depSympt post fixed effects adjustment (for use in interaction analysis with schizophrenia PRS), in the available UK Biobank study population.

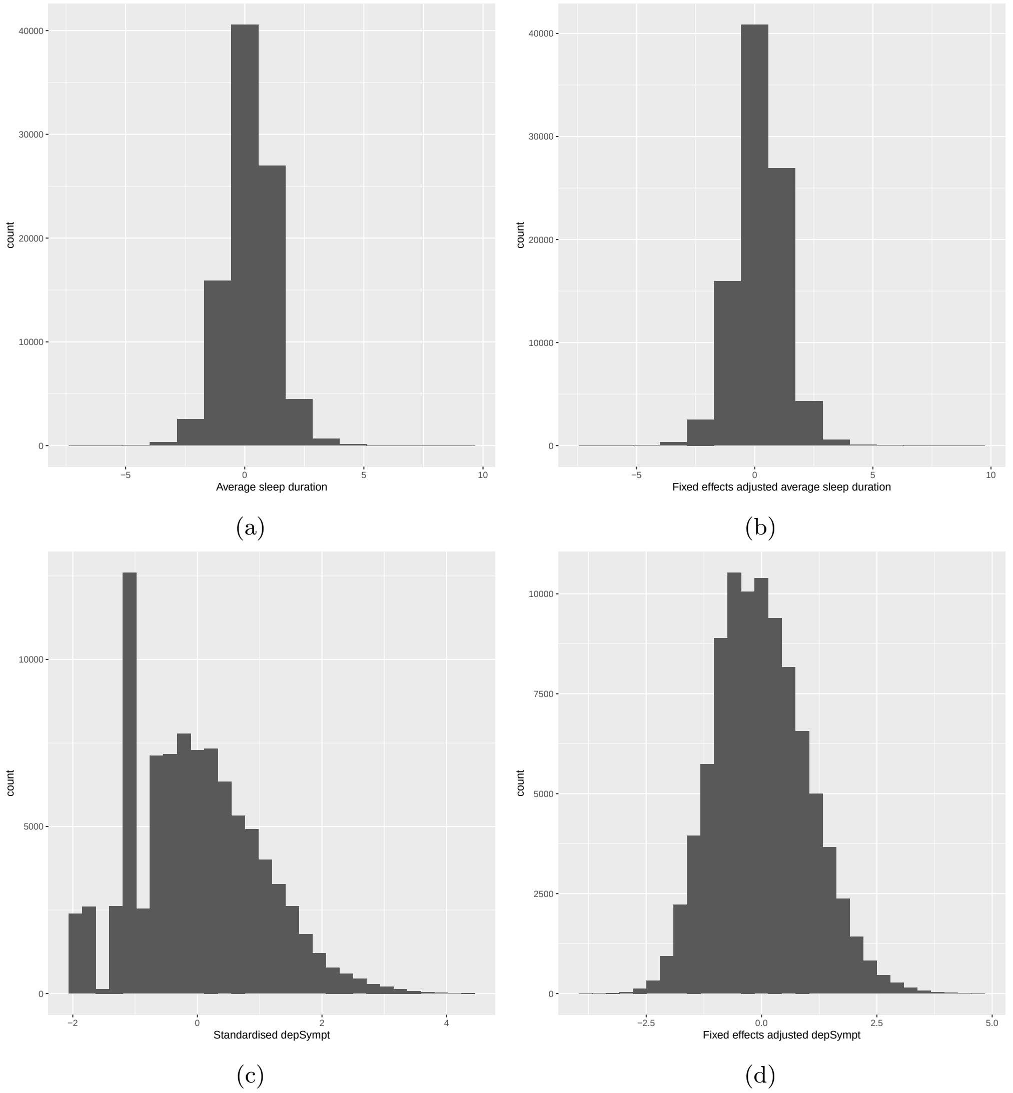

Figure 17: Histogram of standardised: (a) average sleep duration, (b) average sleep duration post fixed effects adjustment, (c) depSympt (Analysis group 1) and (d) depSympt post fixed effects adjustment (for use in interaction analysis with average sleep duration), in the available UK Biobank study population.

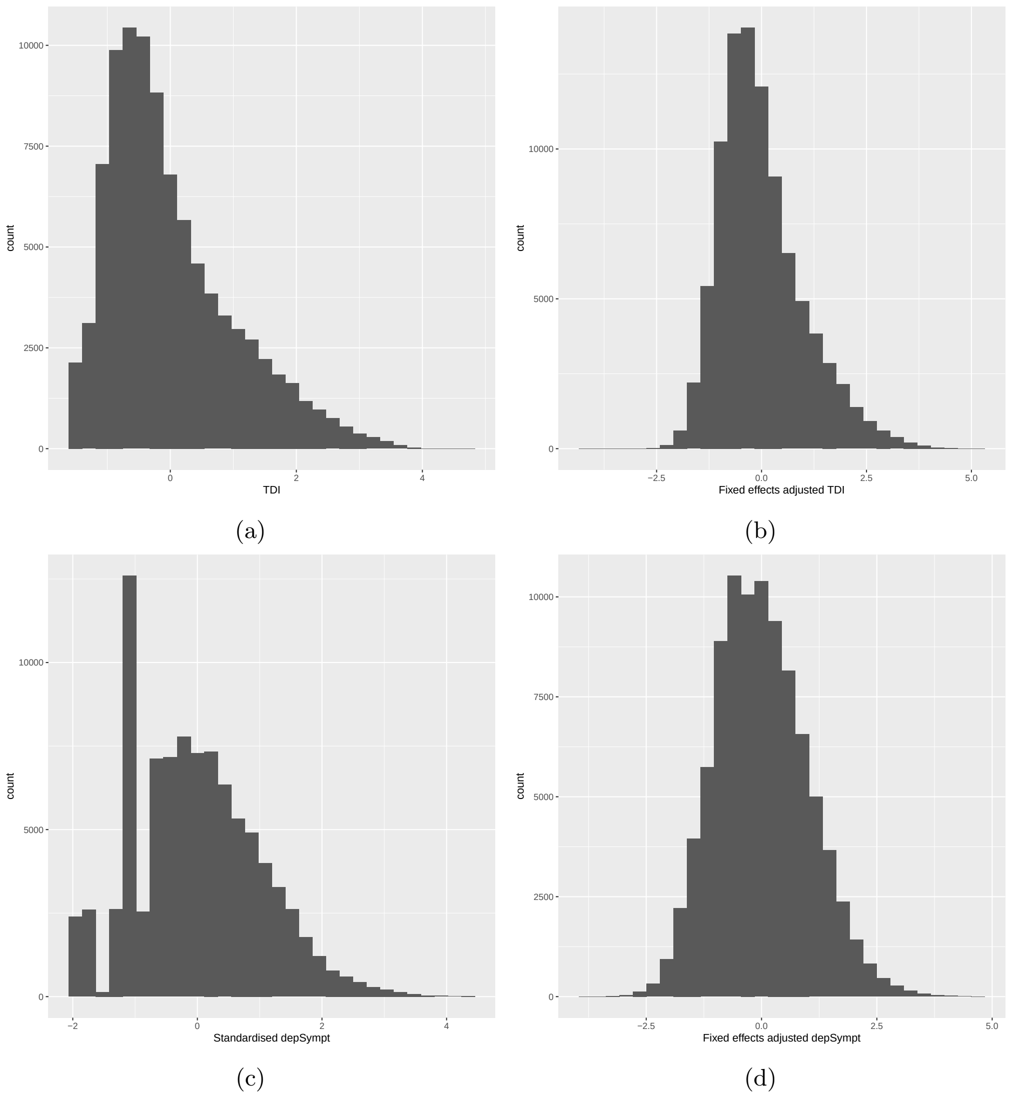

Figure 18: Histogram of standardised: (a) TDI, (b) TDI post fixed effects adjustment, (c) depSympt (Analysis group 1) and (d) depSympt post fixed effects adjustment (for use in interaction analysis with TDI), in the available UK Biobank study population.

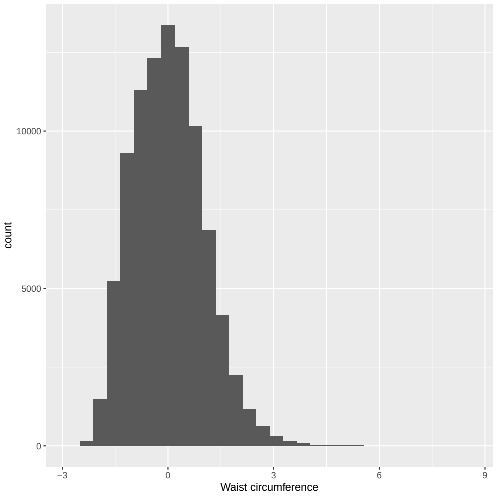

(a)

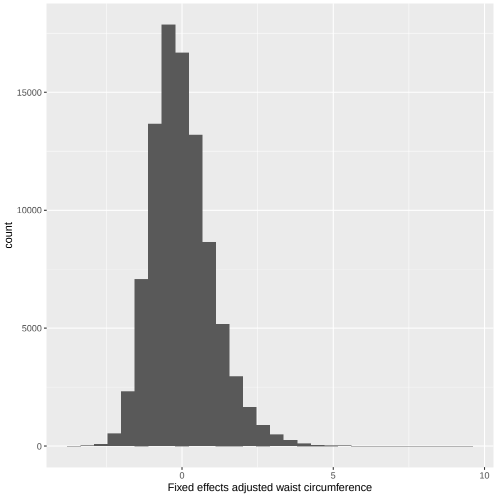

(b)

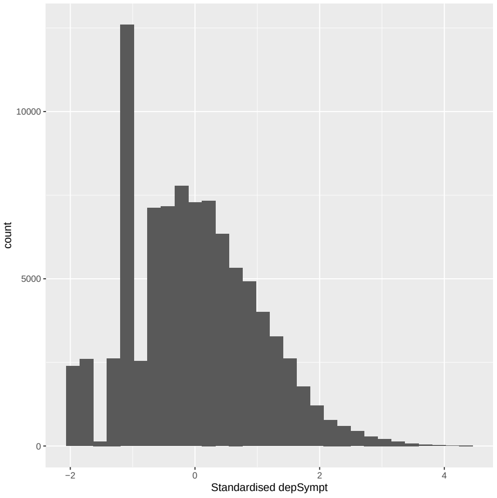

(c)

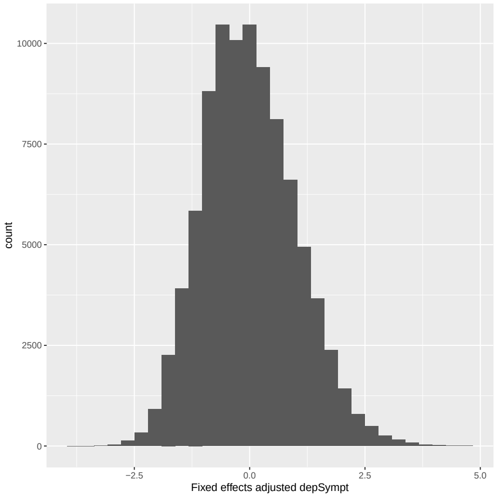

(d)

Figure 19: Histogram of standardised: (a) waist circumference, (b) waist circumference post fixed effects adjustment, (c) depSympt (Analysis group 1) and (d) depSympt post fixed effects adjustment (for use in interaction analysis with waist circumference), in the available UK Biobank study population.

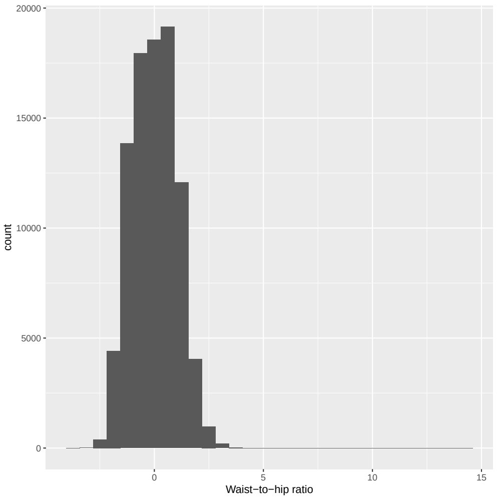

(a)

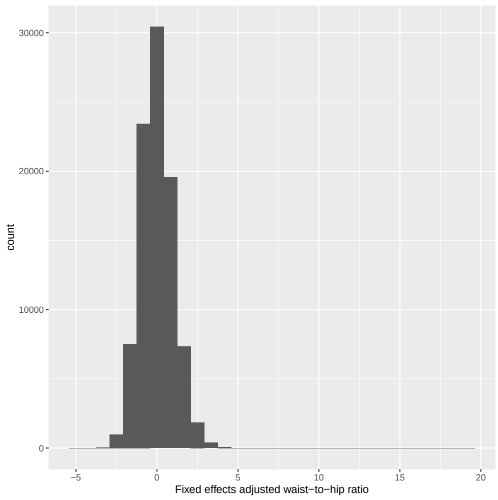

(b)

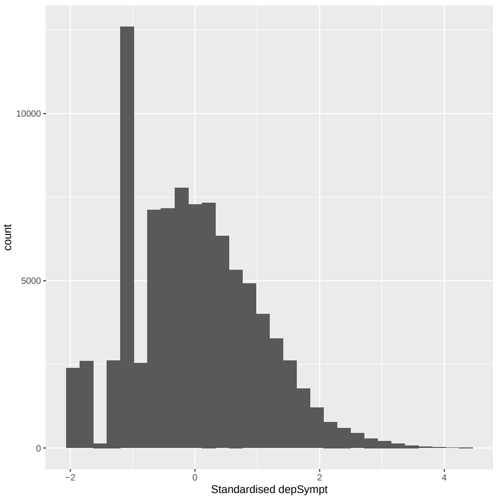

(c)

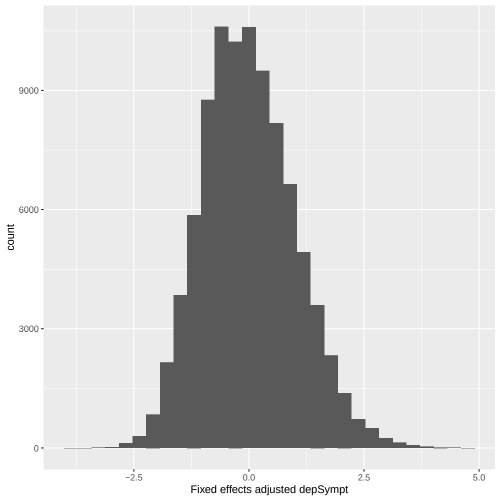

(d)

Figure 20: Histogram of standardised: (a) waist to hip ratio, (b) waist to hip ratio post fixed effects adjustment, (c) depSympt (Analysis group 1) and (d) depSympt post fixed effects adjustment (for use in interaction analysis with waist to hip ratio), in the available UK Biobank study population.

##### 3.1.2 Covariates in Analysis group 2

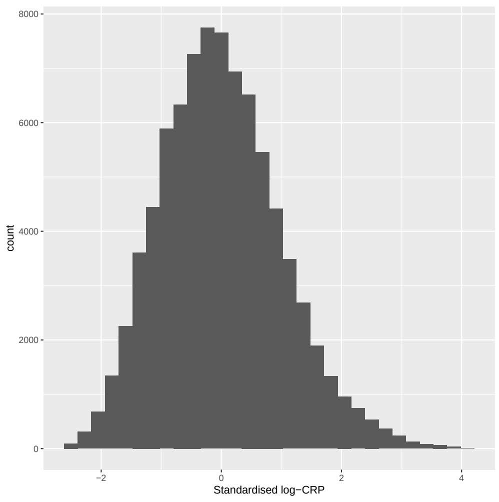

(a)

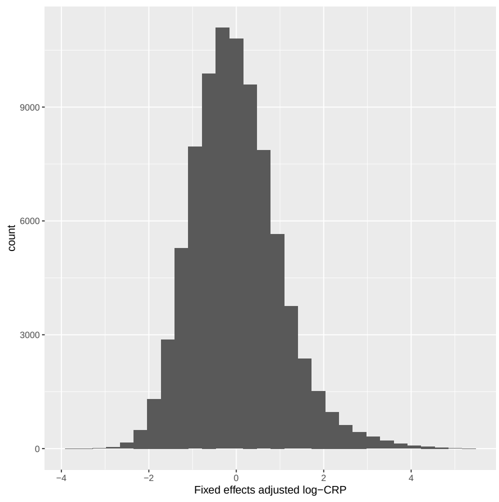

(b)

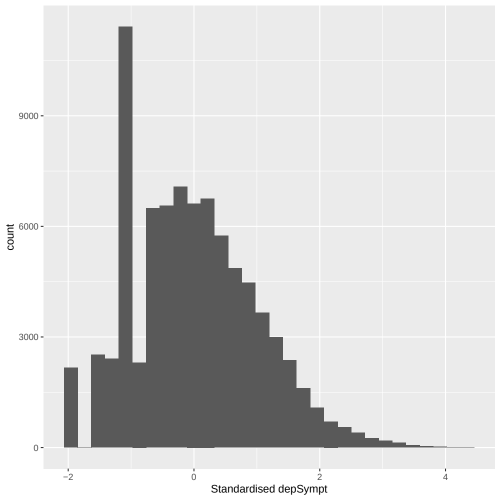

(c)

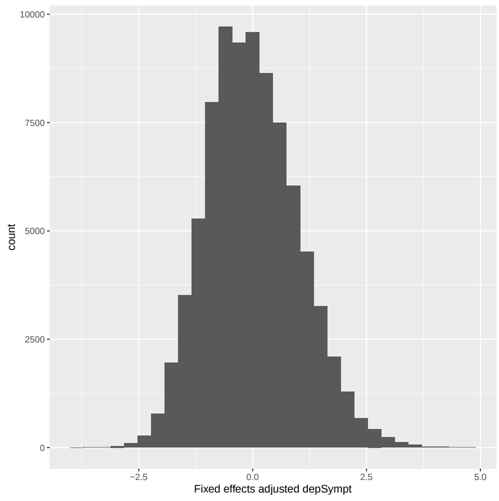

(d)

Figure 21: Histogram of standardised: (a) log-CRP, (b) log-CRP post fixed effects adjustment, (c) depSympt (Analysis group 2) and (d) depSympt post fixed effects adjustment (for use in interaction analysis with all group 2 covariates), in the available UK Biobank study population.

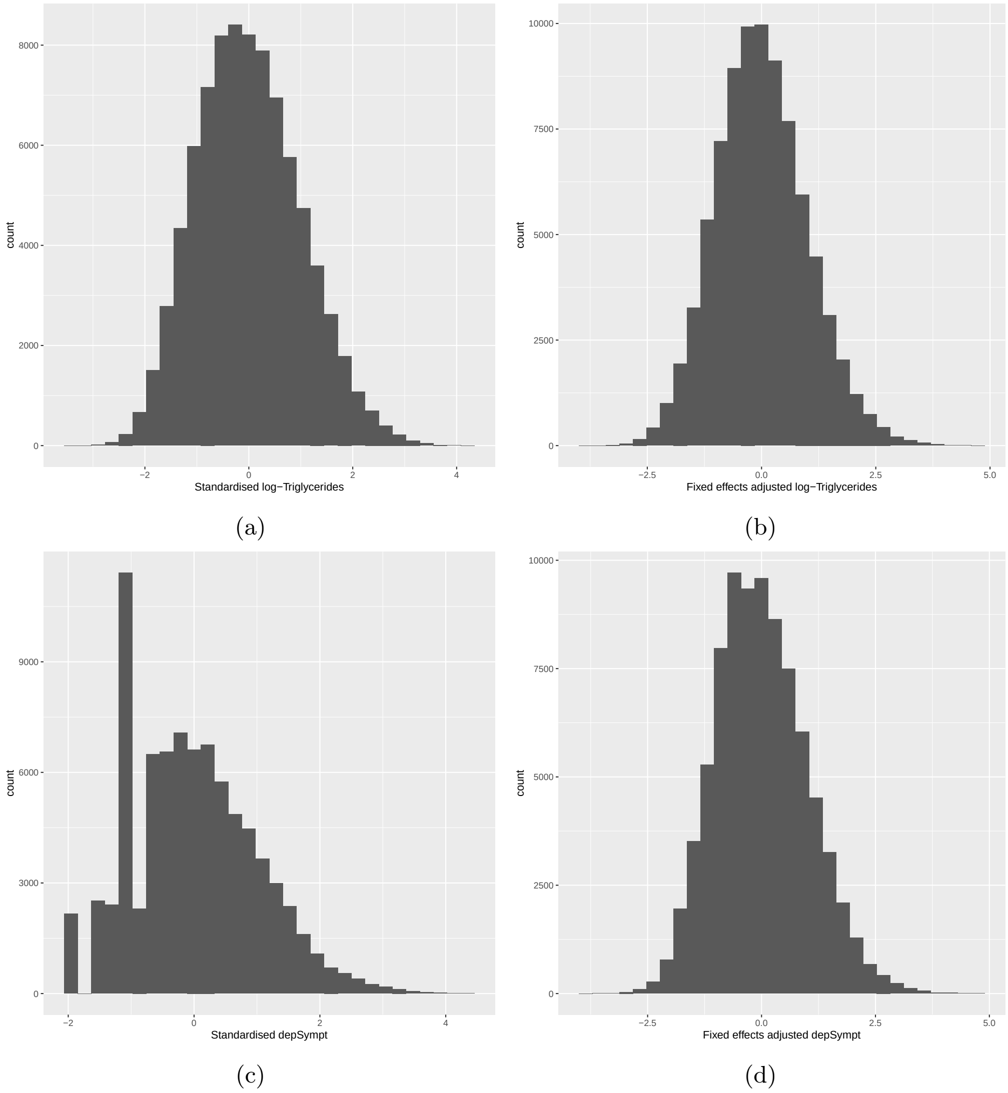

Figure 22: Histogram of standardised: (a) log-Triglycerides, (b) log-Triglycerides post fixed effects adjustment, (c) depSympt (Analysis group 2) and (d) depSympt post fixed effects adjustment (for use in interaction analysis with all group 2 covariates), in the available UK Biobank study population.

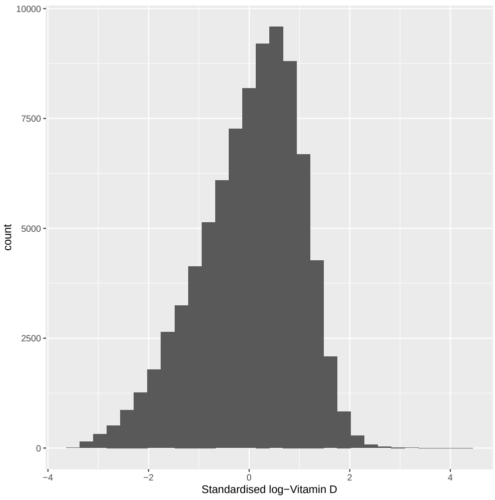

(a)

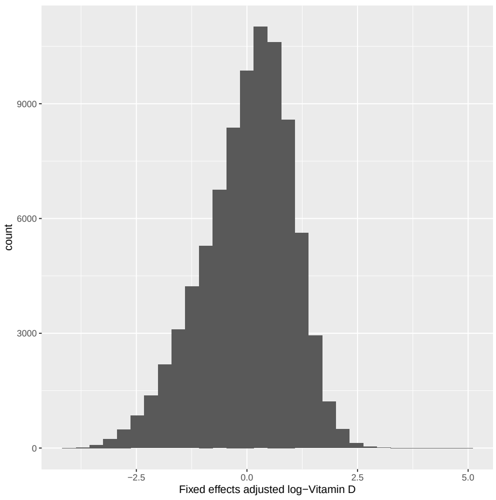

(b)

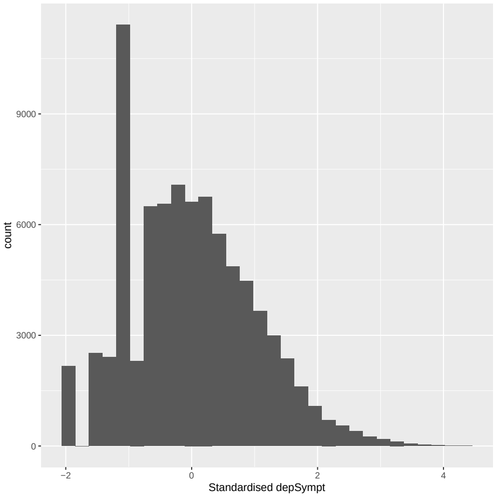

(c)

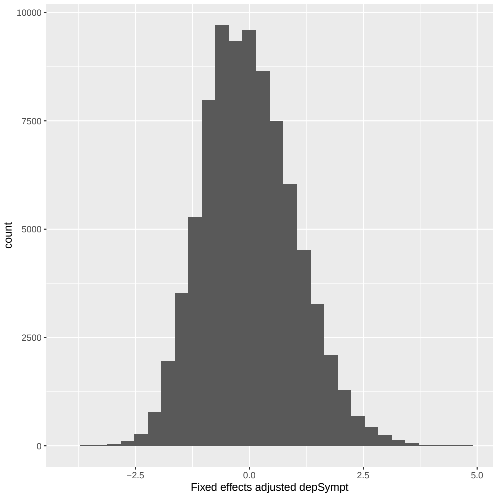

(d)

Figure 23: Histogram of standardised: (a) log-Vitamin D, (b) log-Vitamin D post fixed effects adjustment, (c) depSympt (Analysis group 2) and (d) depSympt post fixed effects adjustment (for use in interaction analysis with all group 2 covariates), in the available UK Biobank study population.

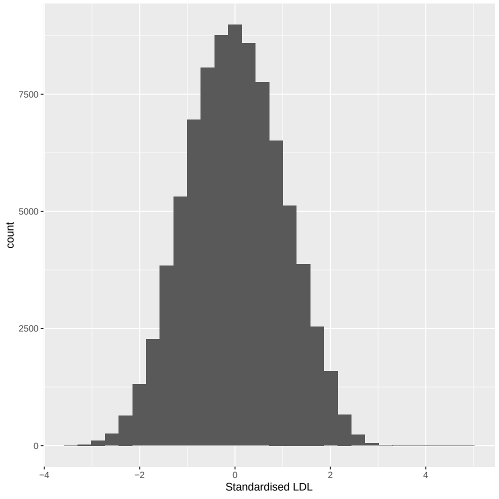

(a)

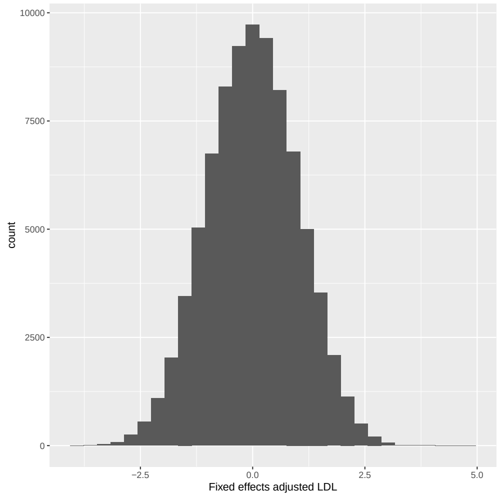

(b)

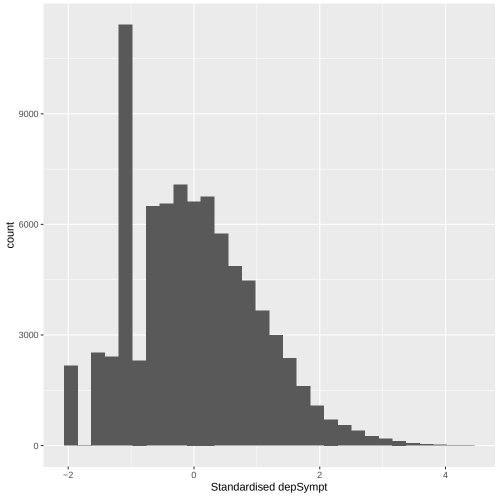

(c)

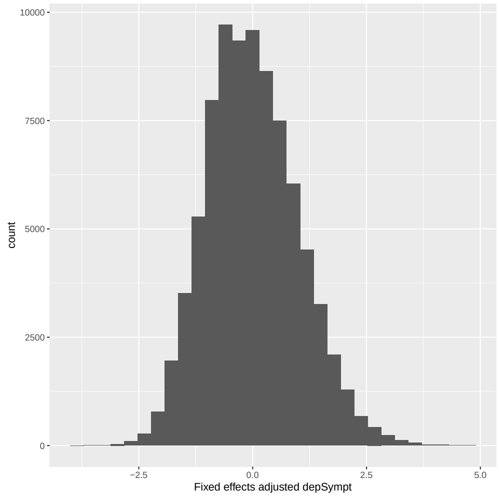

(d)

Figure 24: Histogram of standardised: (a) LDL, (b) LDL post fixed effects adjustment, (c) depSympt (Analysis group 2) and (d) depSympt post fixed effects adjustment (for use in interaction analysis with all group 2 covariates), in the available UK Biobank study population.

##### 3.1.3 Covariates in Analysis group 3

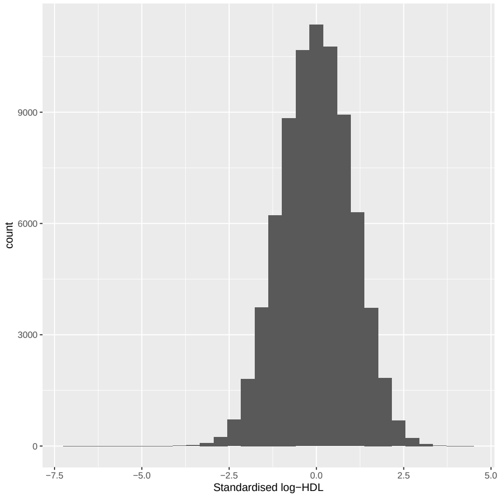

(a)

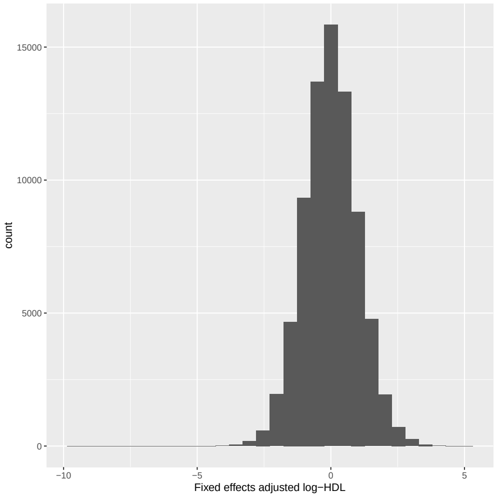

(b)

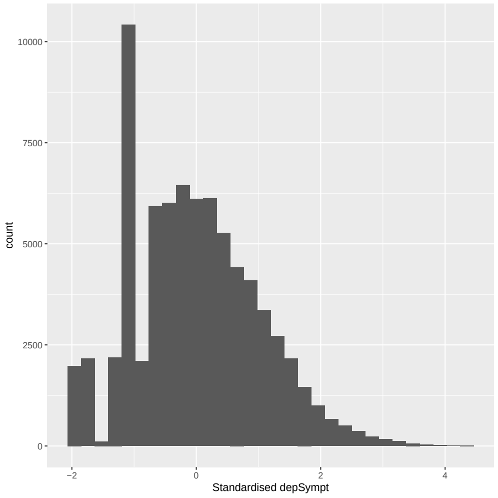

(c)

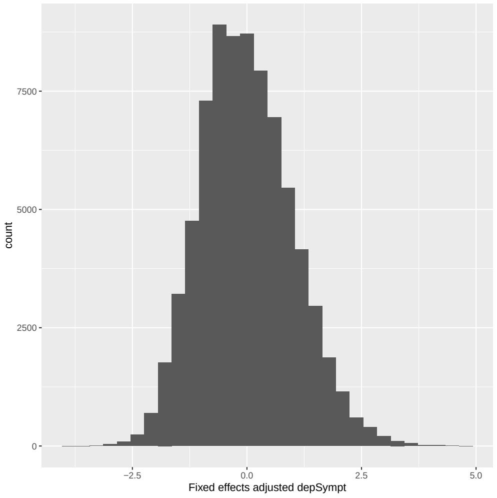

(d)

Figure 25: Histogram of standardised: (a) log-HDL, (b) log-HDL post fixed effects adjustment, (c) depSympt (Analysis group 3) and (d) depSympt post fixed effects adjustment (for use in interaction analysis with log-HDL), in the available UK Biobank study population.

##### 3.1.4 Covariates in Analysis group 4

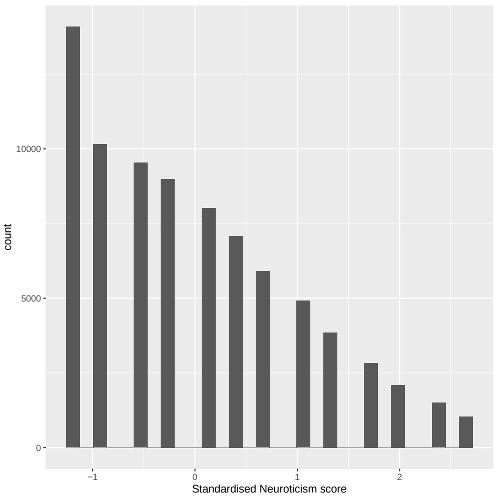

(a)

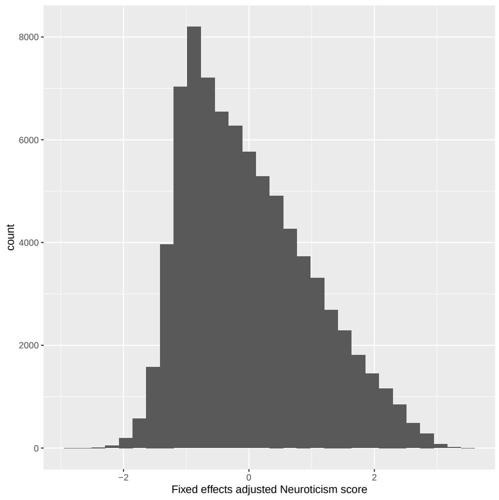

(b)

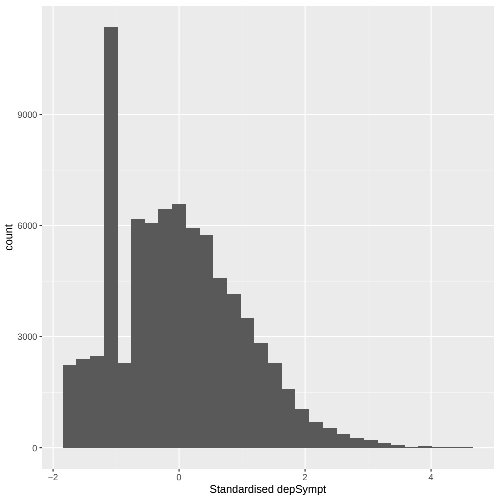

(c)

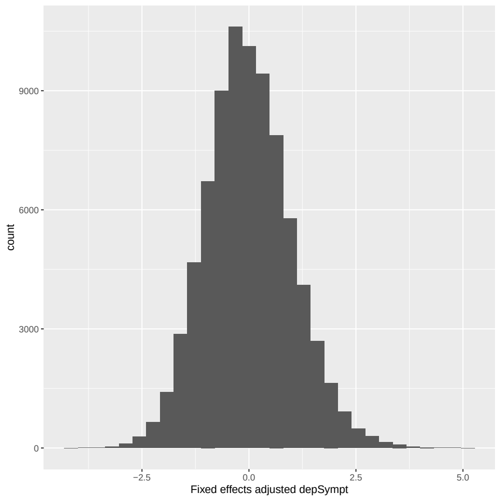

(d)

Figure 26: Histogram of standardised: (a) Neuroticism score, (b) Neuroticism score post fixed effects adjustment, (c) depSympt (Analysis group 4) and (d) depSympt post fixed effects adjustment (for use in interaction analysis with Neuroticism score), in the available UK Biobank study population.

##### 3.1.5 Covariates in Analysis group 5

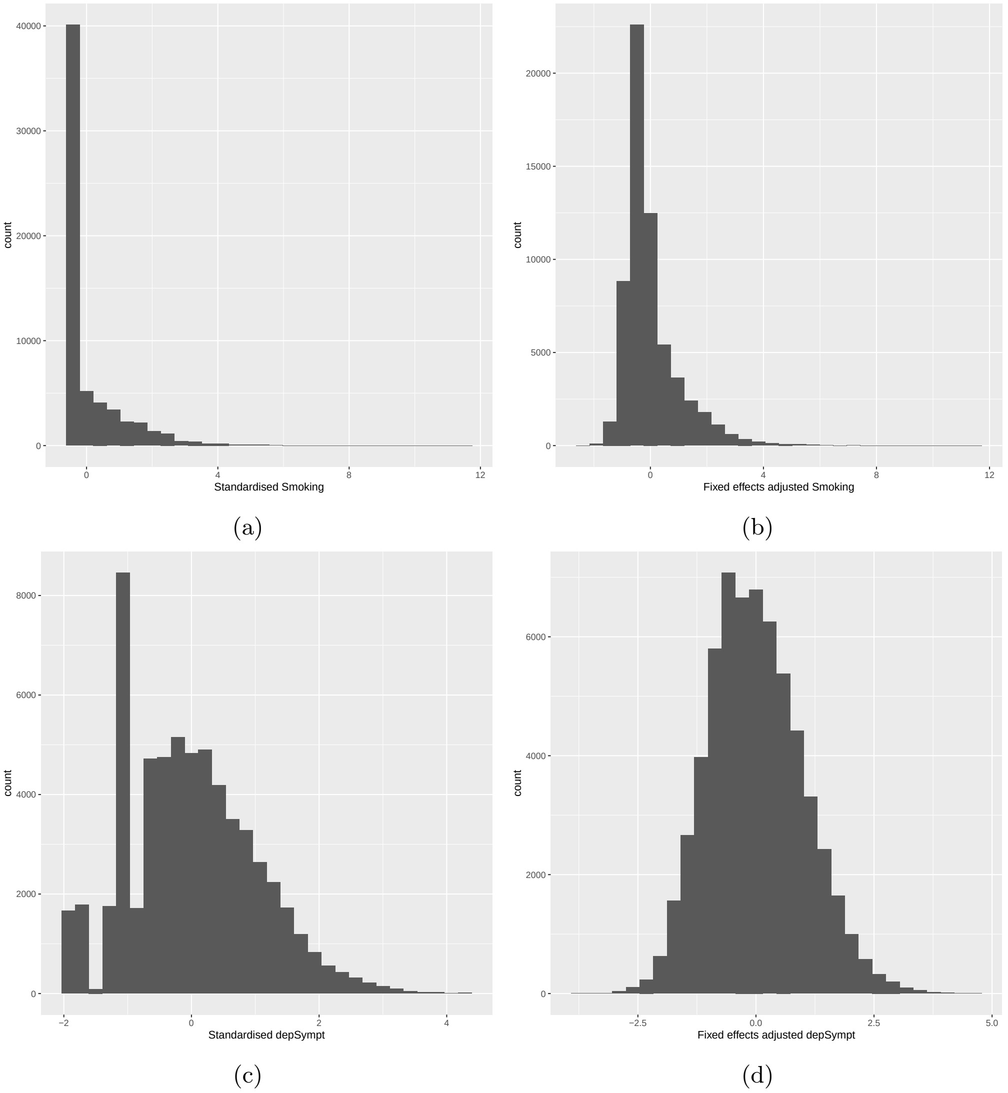

Figure 27: Histogram of standardised: (a) Smoking, (b) Smoking post fixed effects adjustment, (c) depSympt (Analysis group 5) and (d) depSympt post fixed effects adjustment (for use in interaction analysis with Smoking), in the available UK Biobank study population.

##### 3.1.6 Biomarker distribution plots: untransformed compared to log transformed

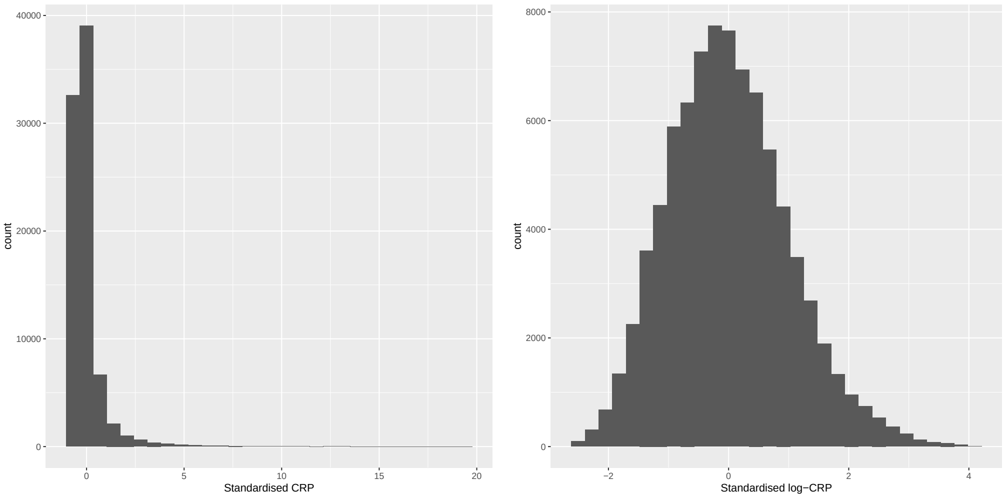

(a) (b)

Figure 28: Histogram of standardised: (a) CRP and (b) log-CRP ($N = 83,489$).

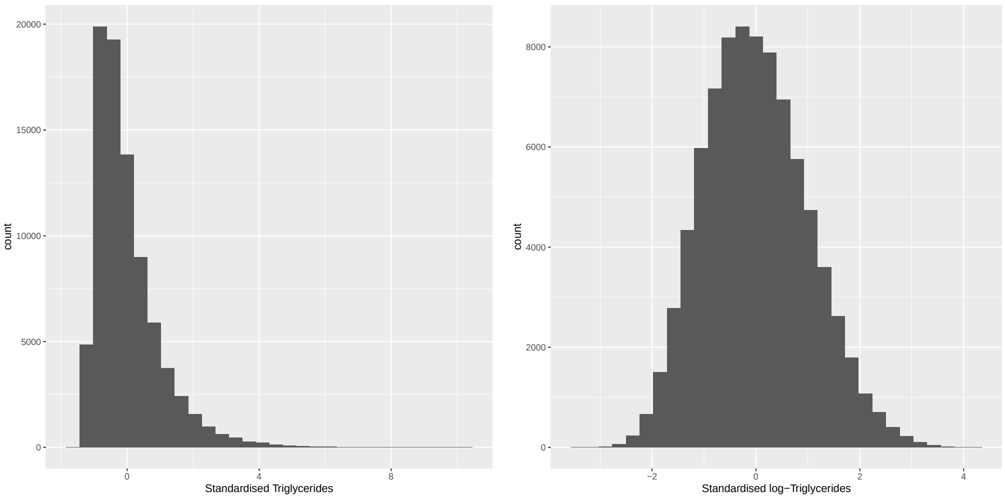

(a) (b)

Figure 29: Histogram of standardised: (a) Triglycerides and (b) log-Triglycerides ($N = 83,489$).

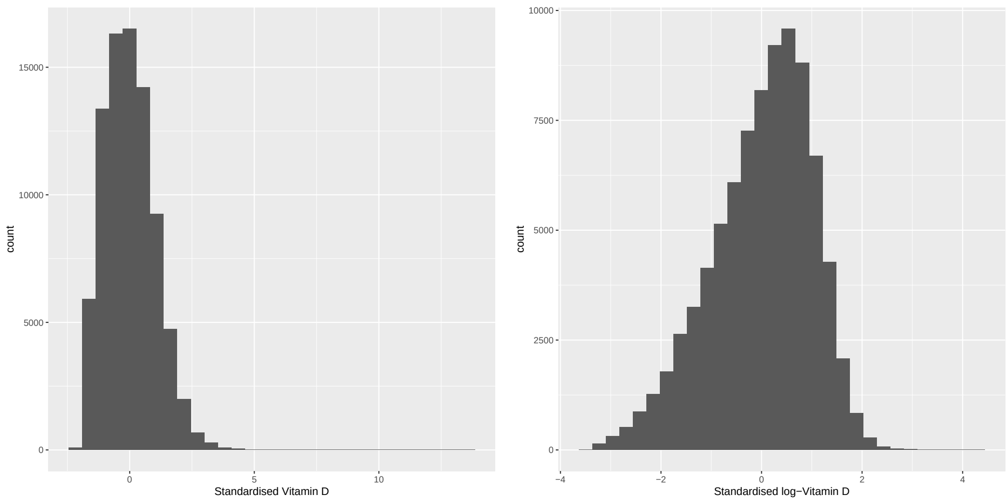

(a) (b)

Figure 30: Histogram of standardised: (a) Vitamin D and (b) log-Vitamin D ($N = 83,489$).

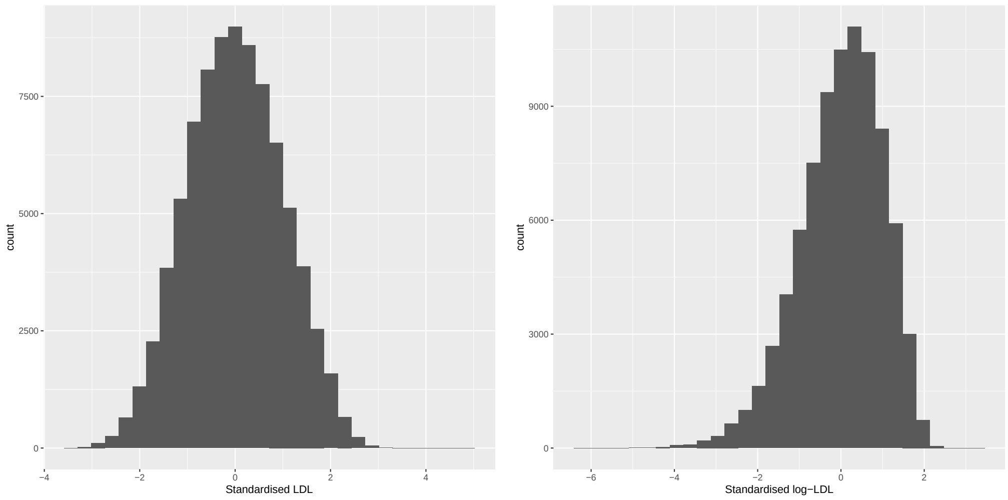

(a) (b)

Figure 31: Histogram of standardised: (a) LDL and (b) log-LDL ($N = 83,489$).

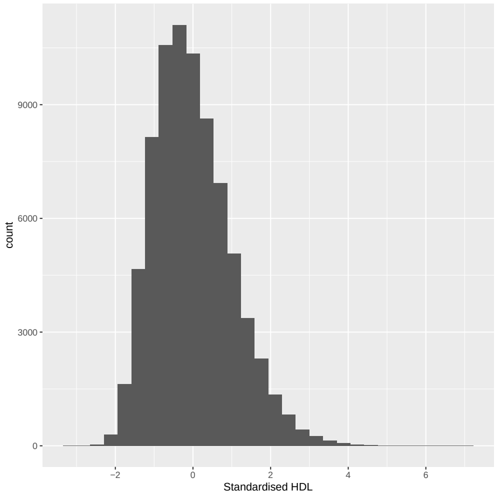

(a)

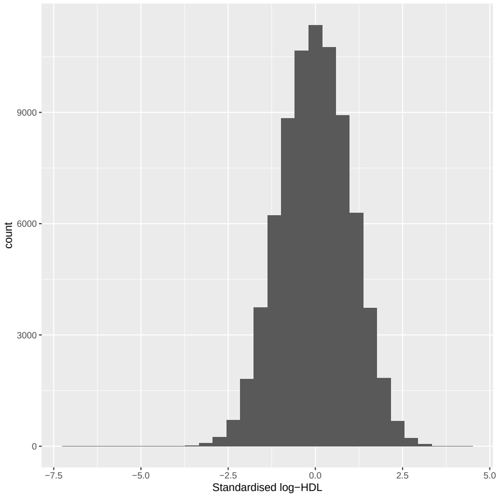

(b)

Figure 32: Histogram of standardised: (a) HDL and (b) log-HDL ($N = 76,246$).

##### 3.2 Fractional polynomial model results

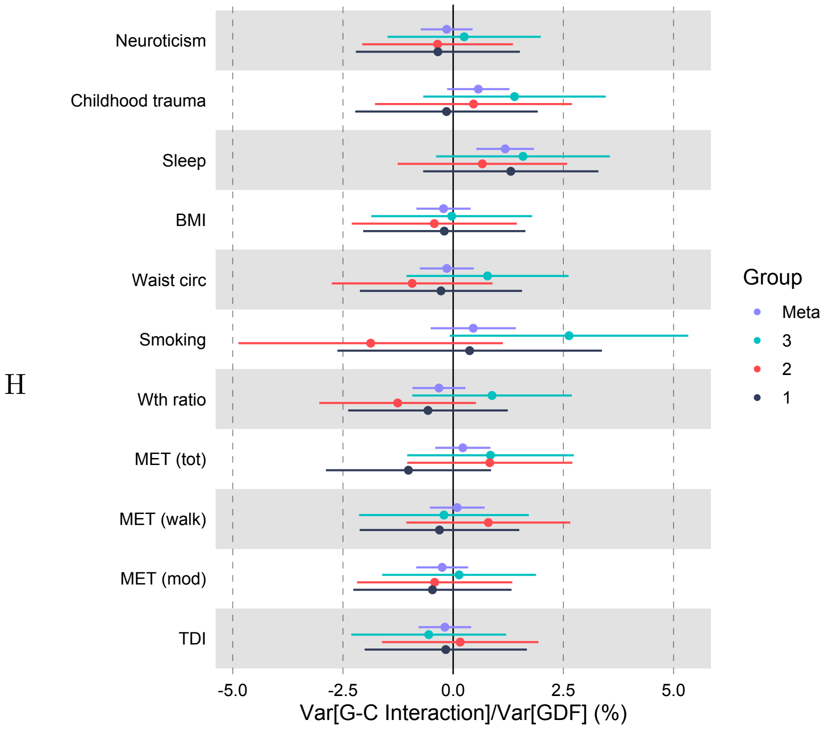

(a) Genotype-Covariate interaction

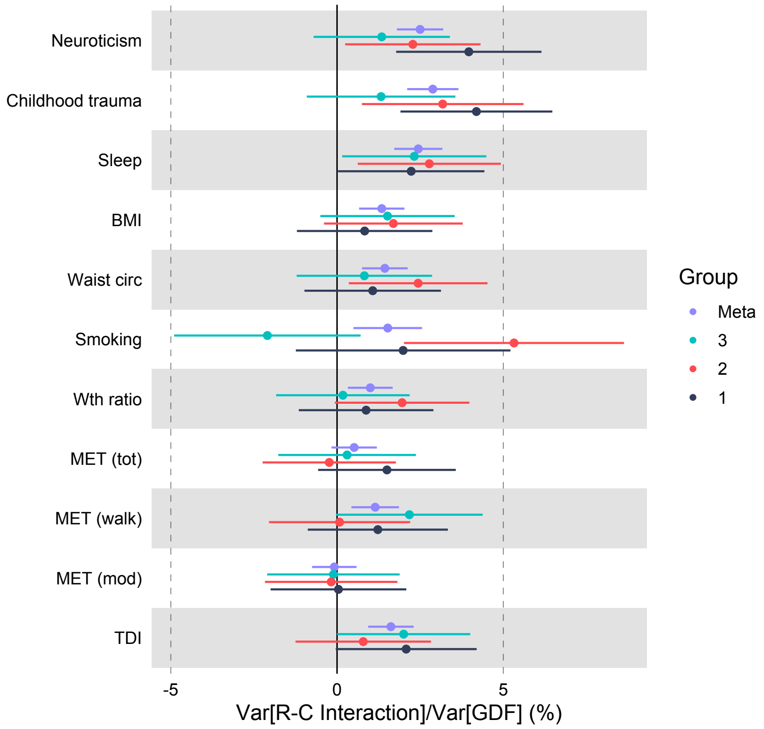

(b) Residual-Covariate interaction

Figure 33: The proportion of variation in depSympt attributable to: (a) a genotype-covariate interaction, and (b) a residual-covariate interaction, with 95% confidence intervals, for all 3 subgroups & the meta-analysis. The fixed effects component of the model is the FP model. Significant variables only.

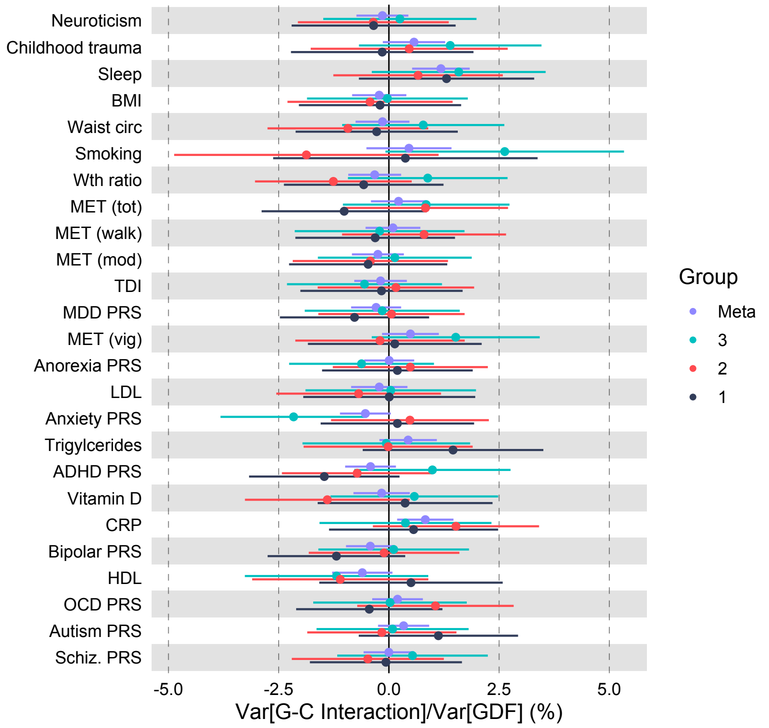

(a) Genotype-Covariate interaction

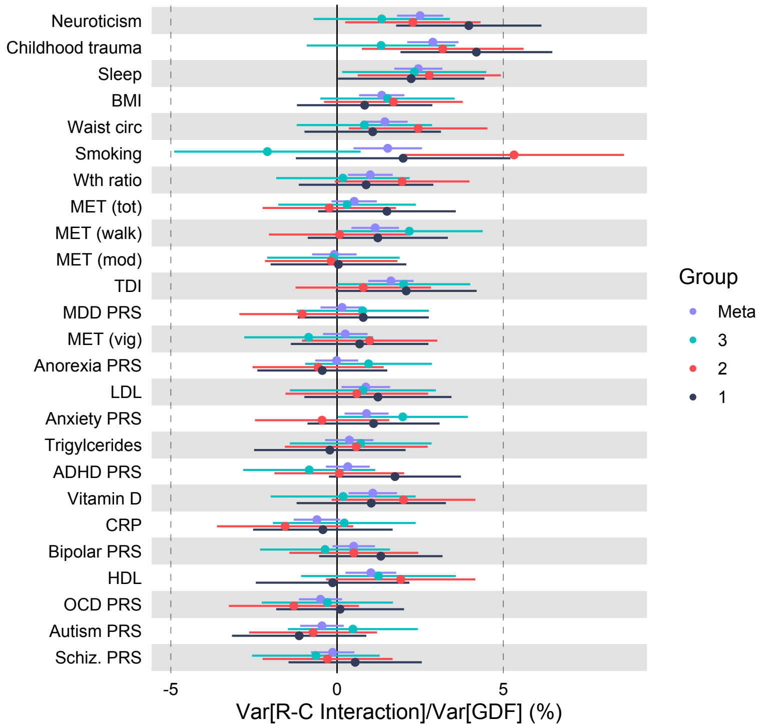

(b) Residual-Covariate interaction

Figure 34: The proportion of variation in depSympt attributable to: (a) a genotype-covariate interaction, and (b) a residual-covariate interaction, with 95% confidence intervals, for all 3 subgroups & the meta-analysis. The fixed effects component of the model is the FP model. All covariate traits.

**Plots: Genetic, residual and total variance components of $Y$ by covariate trait value**

**Significant covariates:** Neuroticism score, childhood trauma summary variable, average sleep duration, BMI, waist circumference, smoking, waist to hip ratio, total MET minutes per week (MET (tot)), walking MET minutes per week (MET (walk)), moderate MET minutes per week (MET (mod)) and TDI.

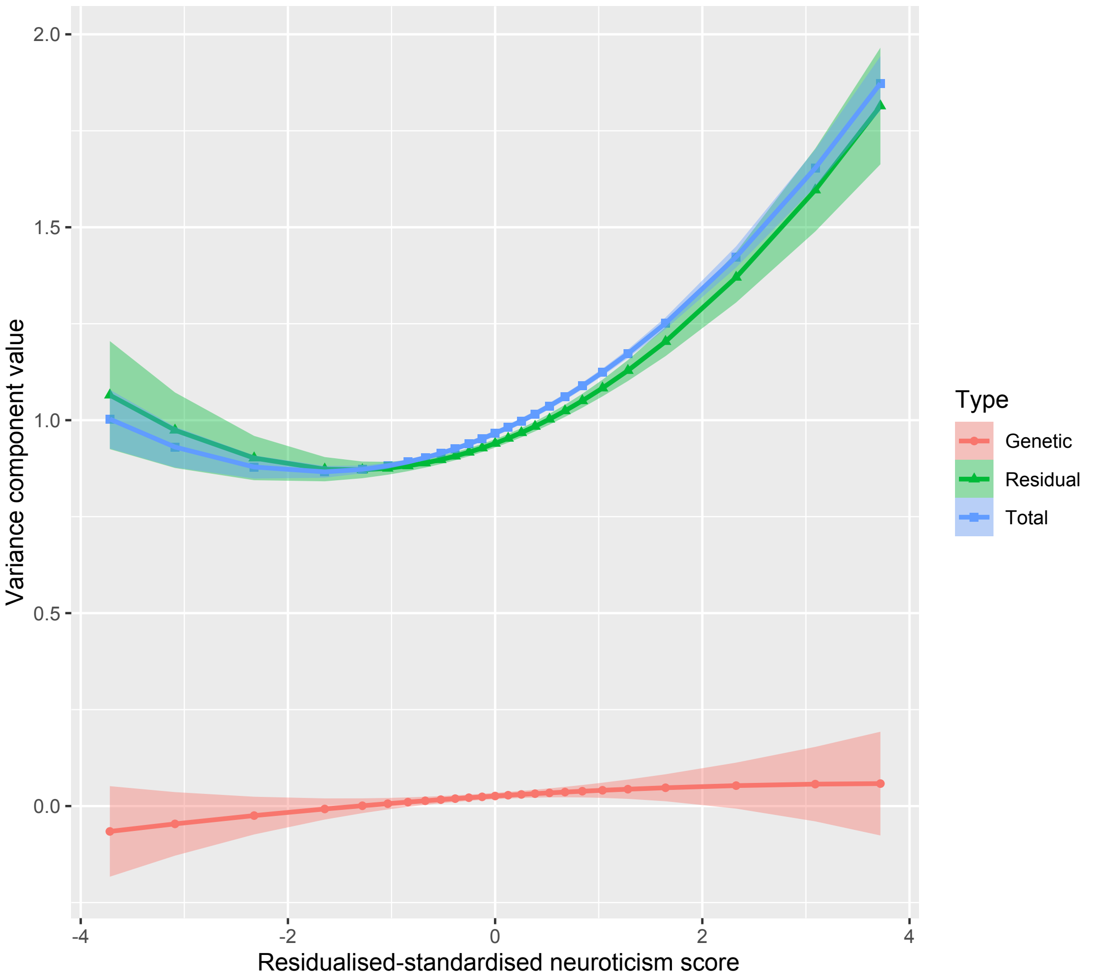

Figure 35: Variance components for $Y$ (residualised depSympt) against residualised and standardised neuroticism score, with 95% confidence intervals.

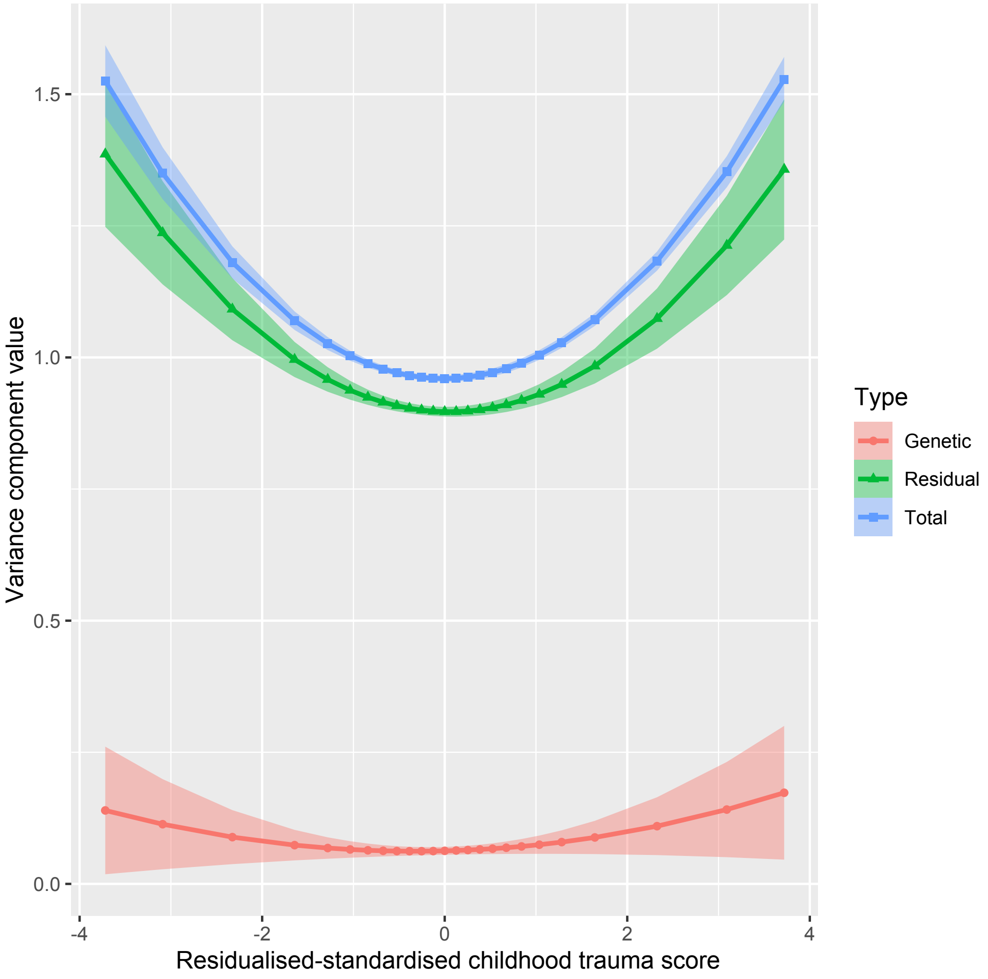

Figure 36: Variance components for $Y$ (residualised depSympt) against residualised and standardised childhood trauma summary variable, with 95% confidence intervals.

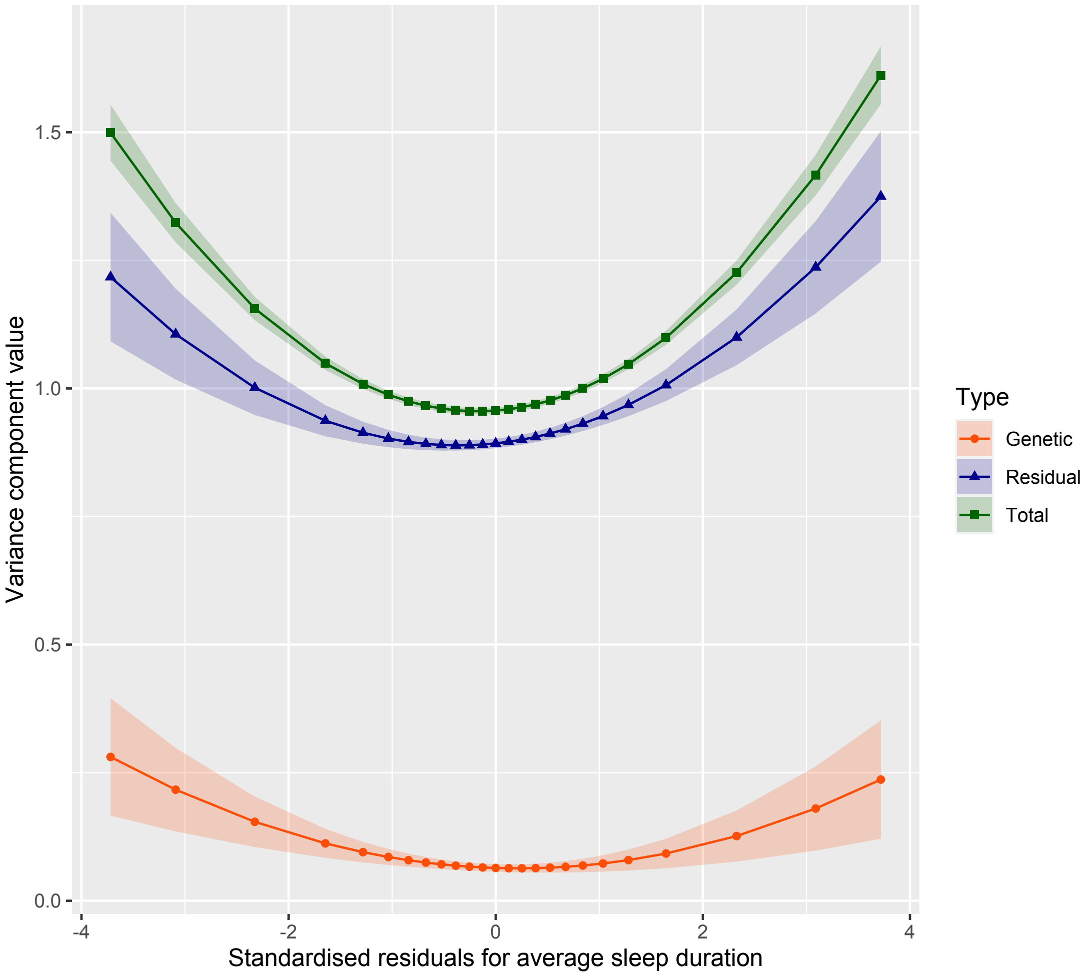

Figure 37: Variance components for $Y$ (residualised depSympt) against residualised and standardised average sleep duration, with 95% confidence intervals.

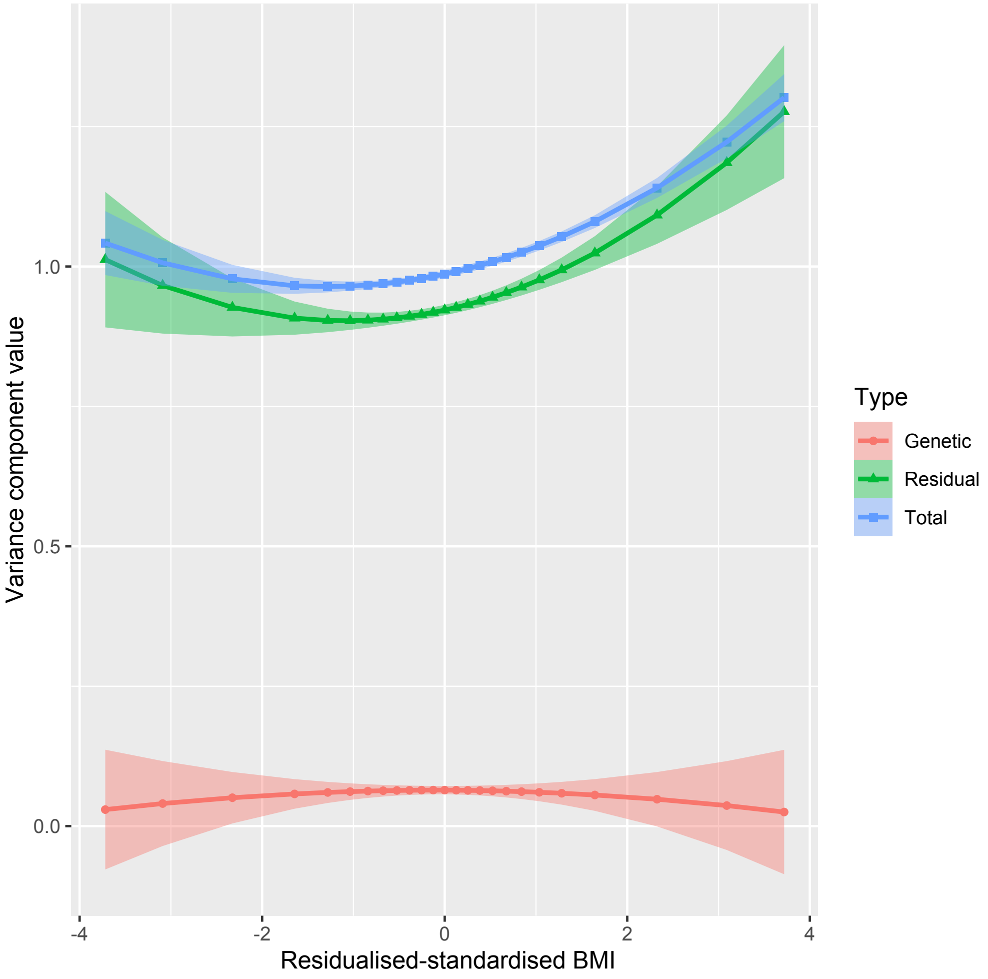

Figure 38: Variance components for $Y$ (residualised depSympt) against residualised and standardised BMI, with 95% confidence intervals.

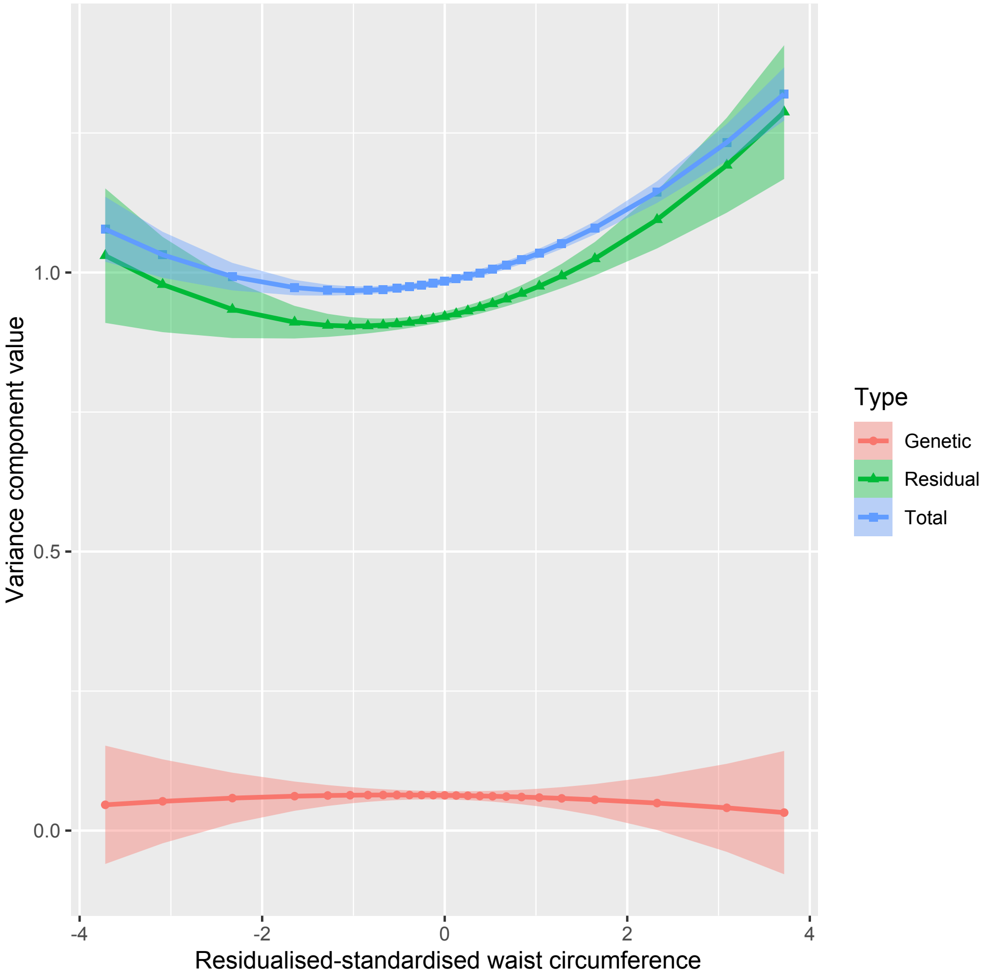

Figure 39: Variance components for $Y$ (residualised depSympt) against residualised and standardised waist circumference, with 95% confidence intervals.

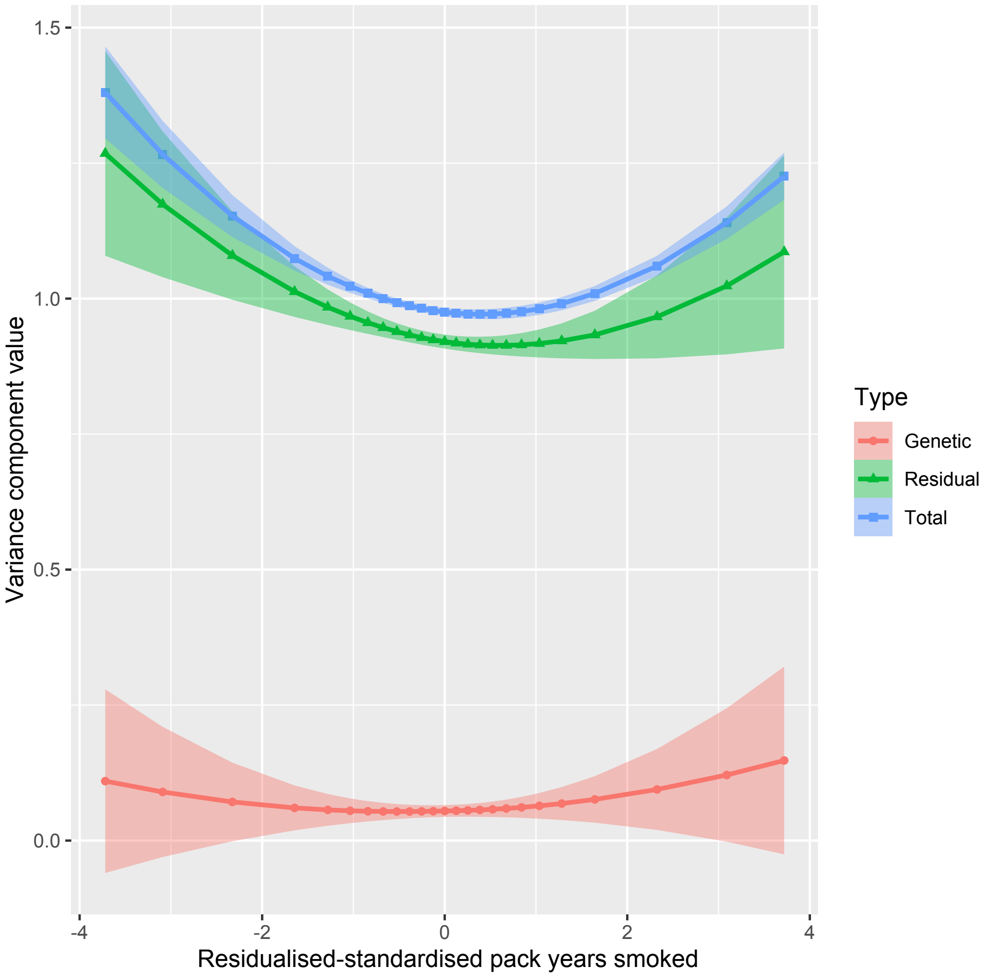

Figure 40: Variance components for $Y$ (residualised depSympt) against residualised and standardised smoking, with 95% confidence intervals.

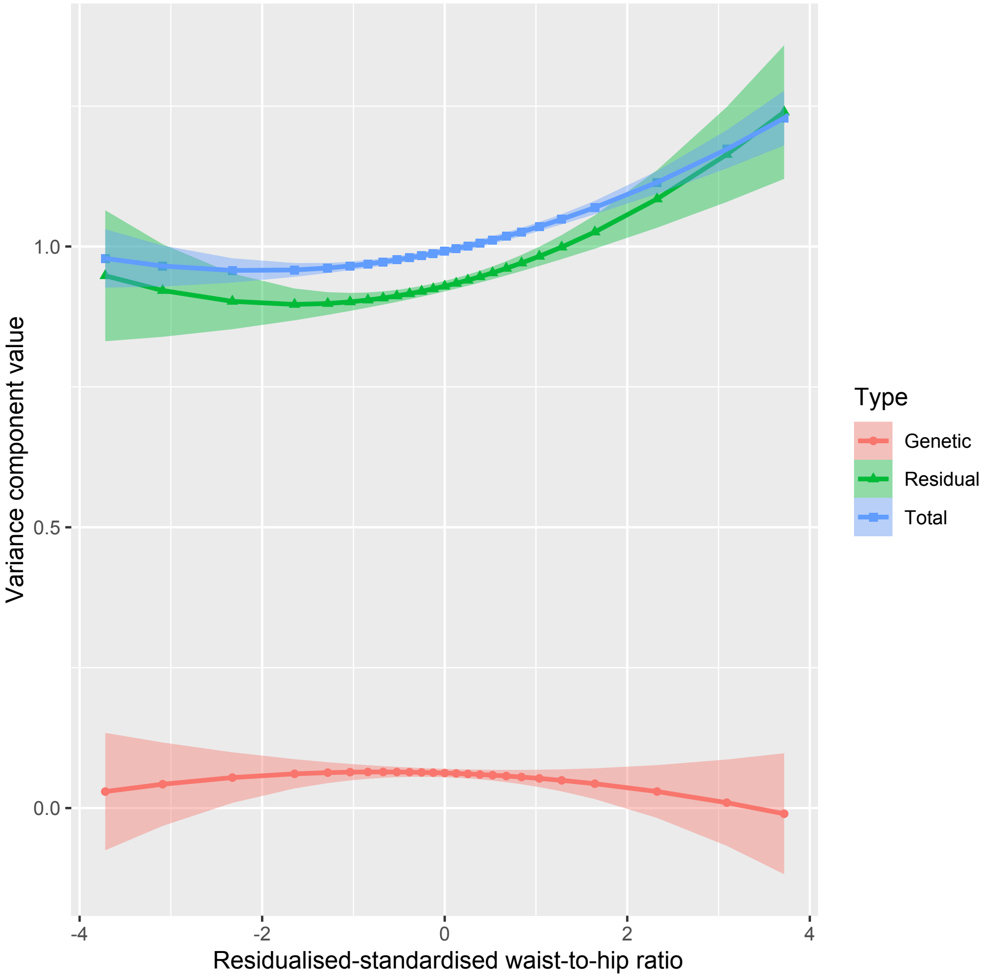

Figure 41: Variance components for $Y$ (residualised depSympt) against residualised and standardised waist to hip ratio, with 95% confidence intervals.

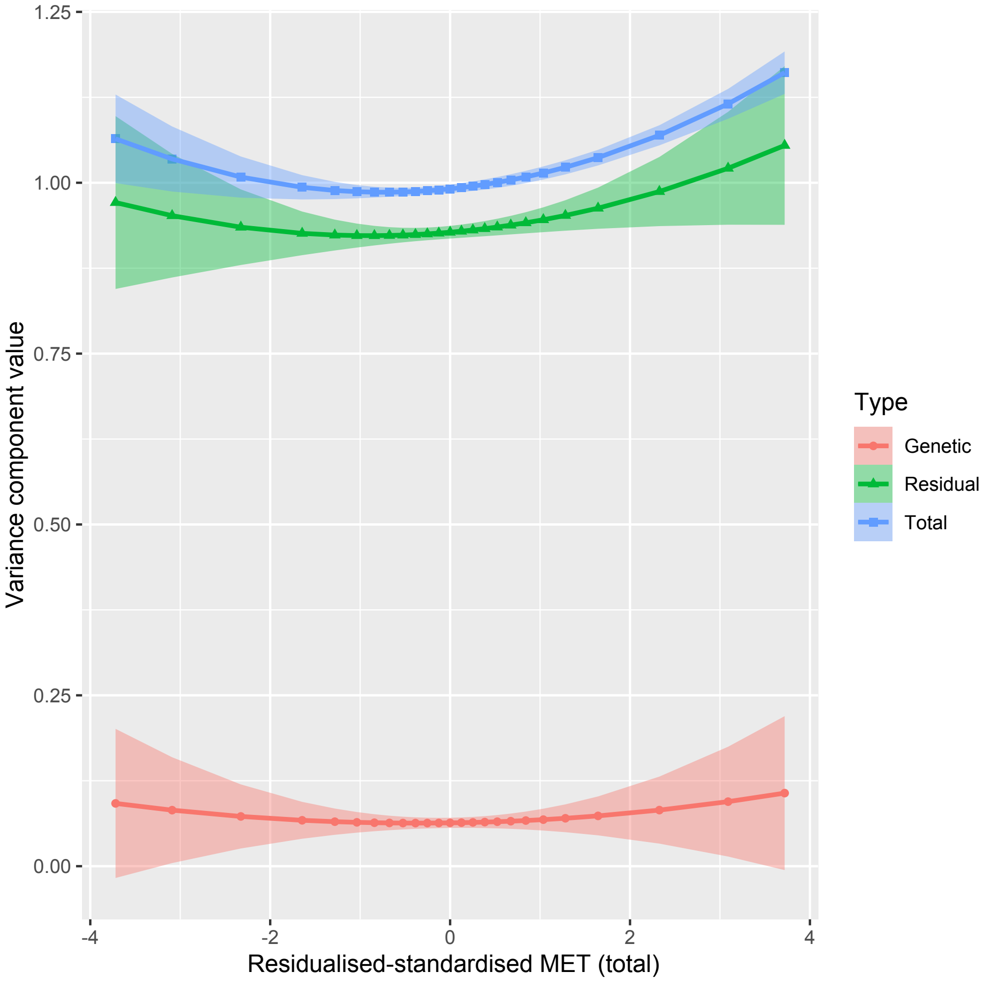

Figure 42: Variance components for $Y$ (residualised depSympt) against residualised and standardised MET (tot), with 95% confidence intervals.

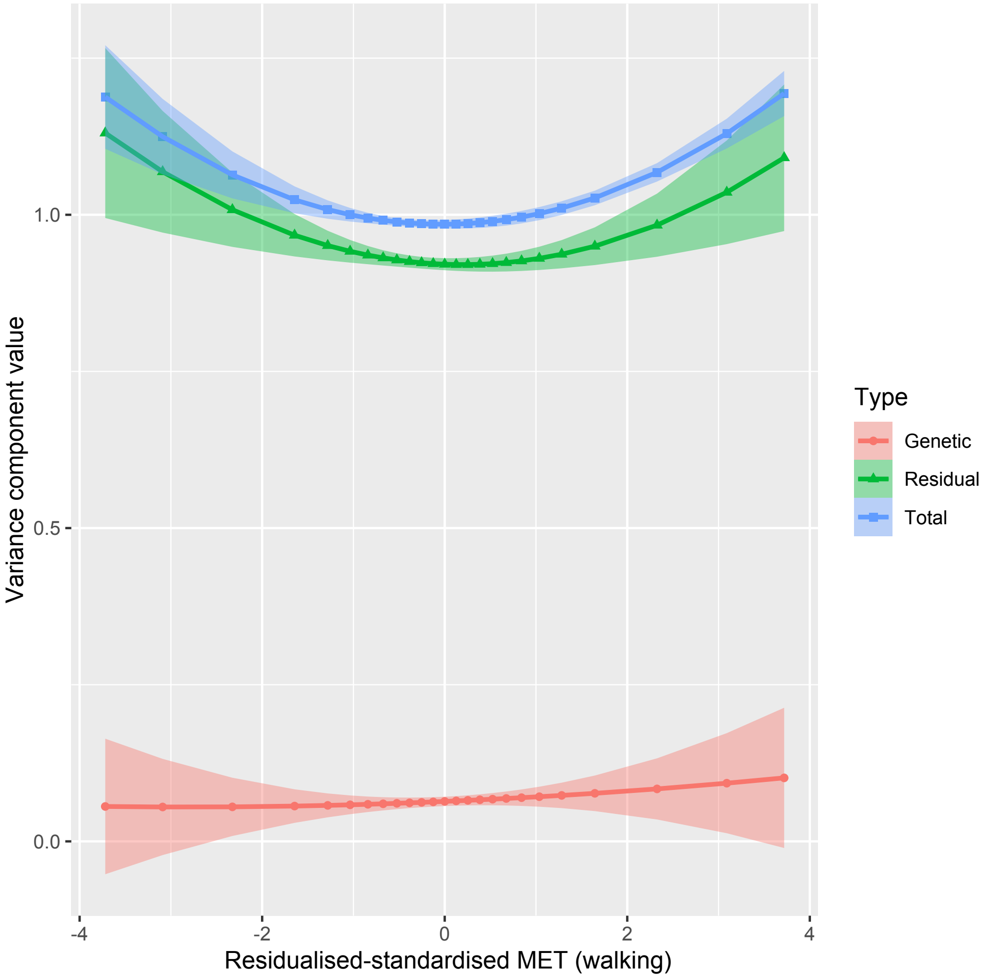

Figure 43: Variance components for $Y$ (residualised depSympt) against residualised and standardised MET (walk), with 95% confidence intervals.

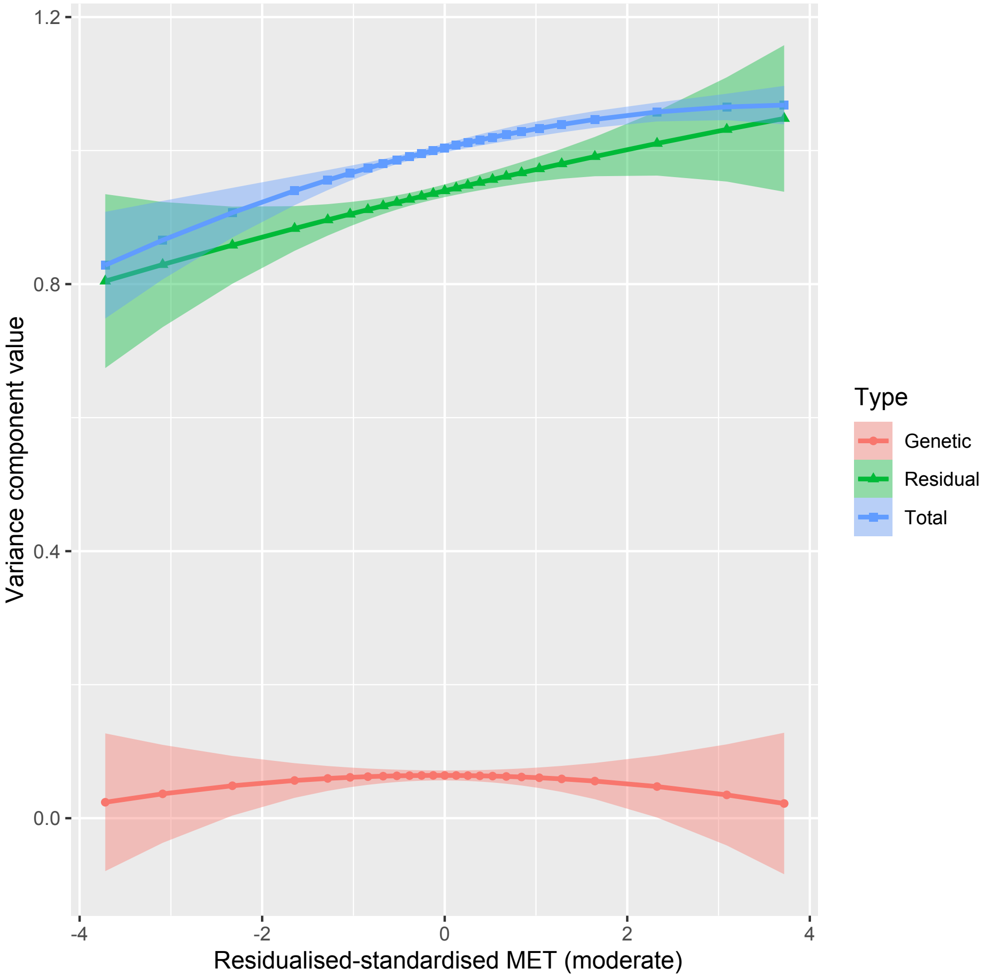

Figure 44: Variance components for $Y$ (residualised depSympt) against residualised and standardised MET (mod), with 95% confidence intervals.

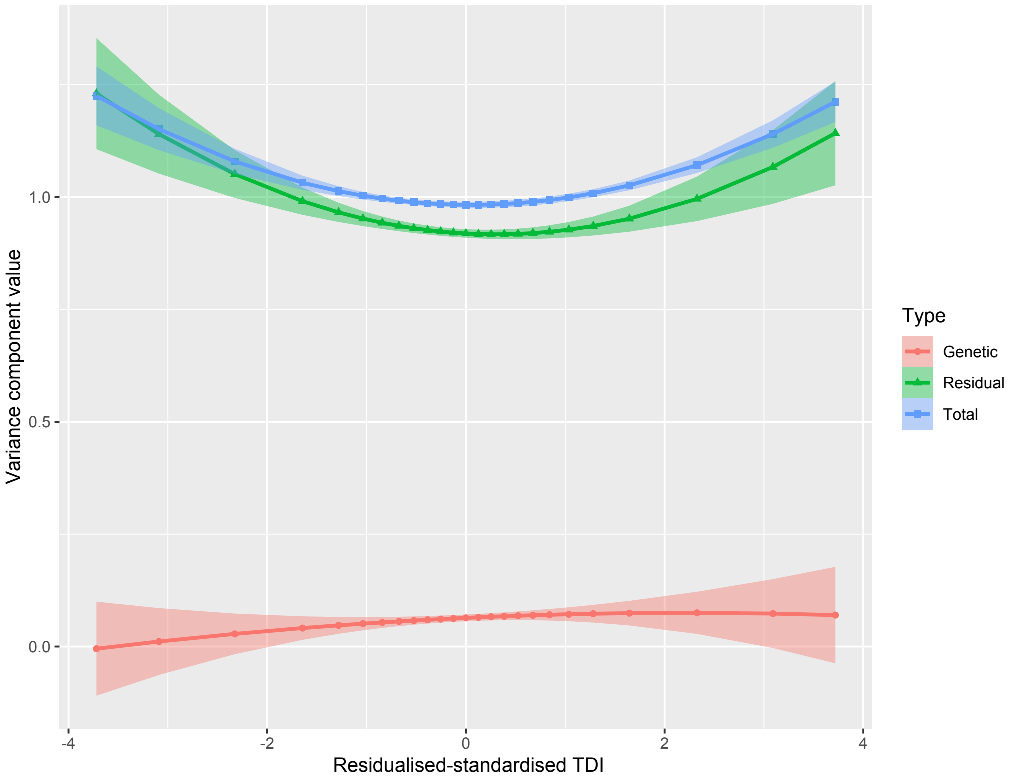

Figure 45: Variance components for $Y$ (residualised depSympt) against residualised and standardised TDI, with 95% confidence intervals.

##### Heritability plot for sleep

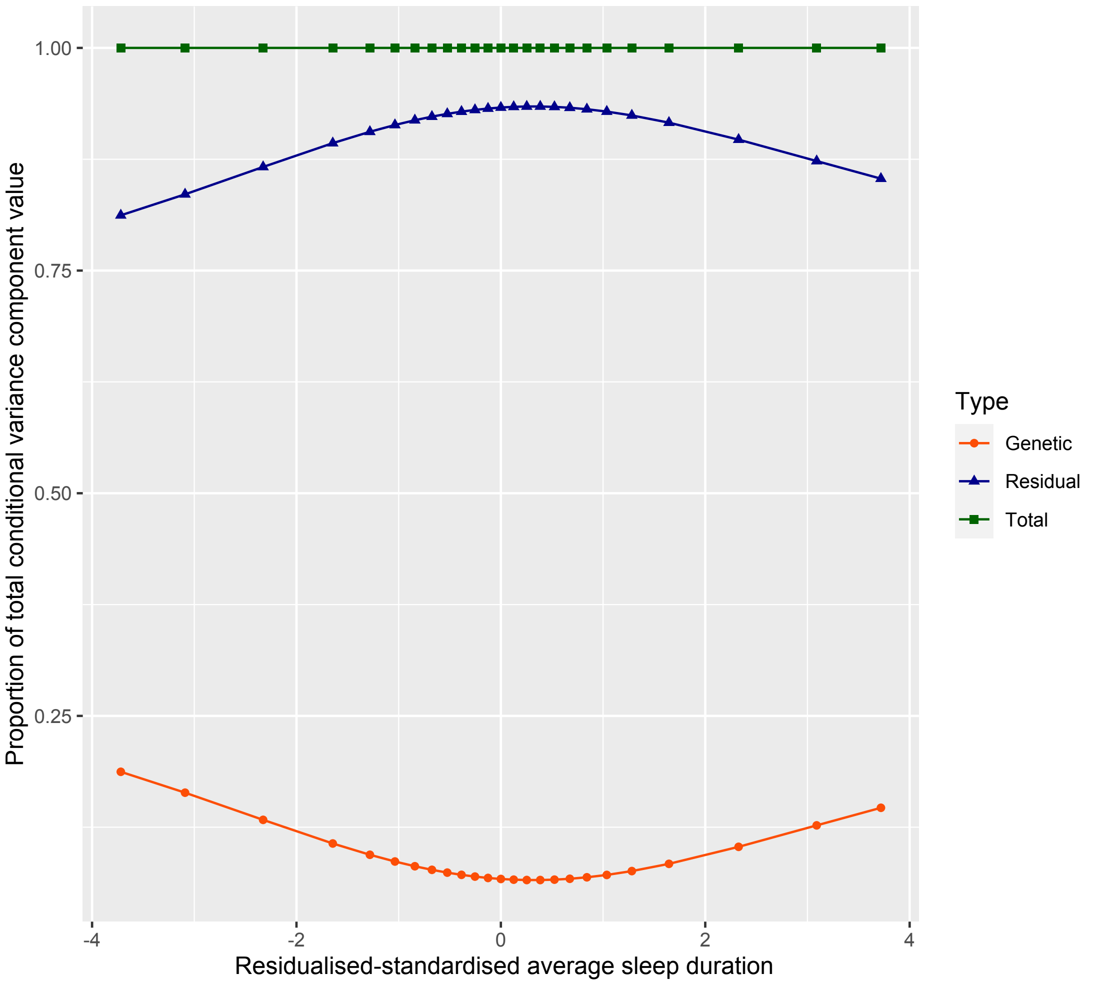

Figure 46: Proportion of the total variance component for $Y$ (residualised depSympt) attributable to the genetic component (heritability) and the residual component, as a function of residualised and standardised average sleep duration, with 95% confidence intervals.

#### Plots for MET (tot) and MET (mod) exploration

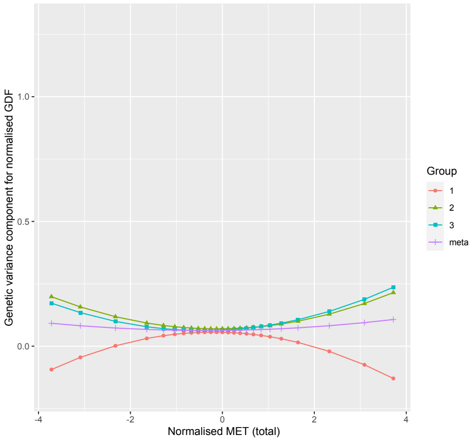

(a) Genetic variance component

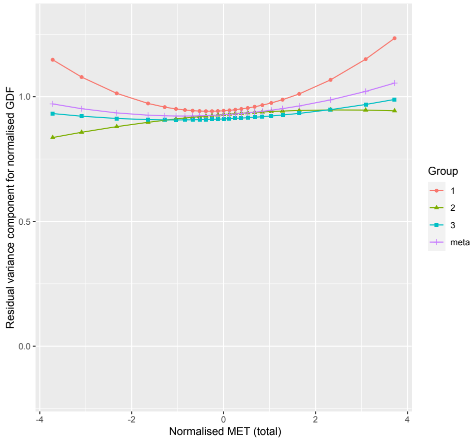

(b) Residual variance component

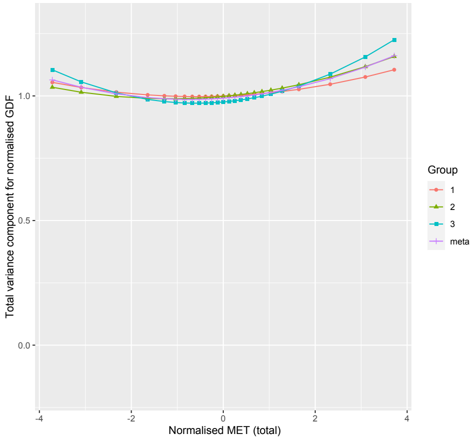

(c) Total variance component

Figure 47: Plots of the variance components for normalised depSympt across normalised MET (total)

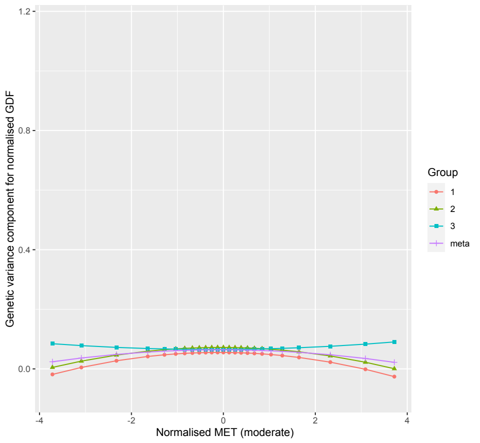

(a) Genetic variance component

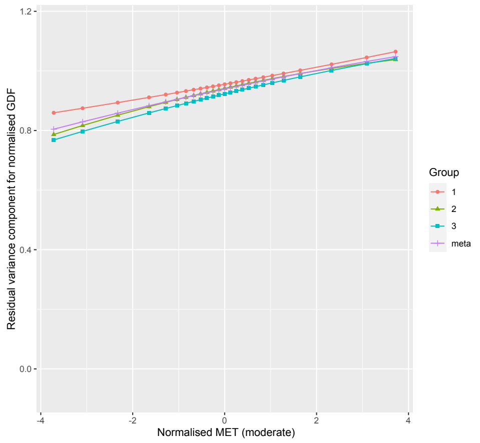

(b) Residual variance component

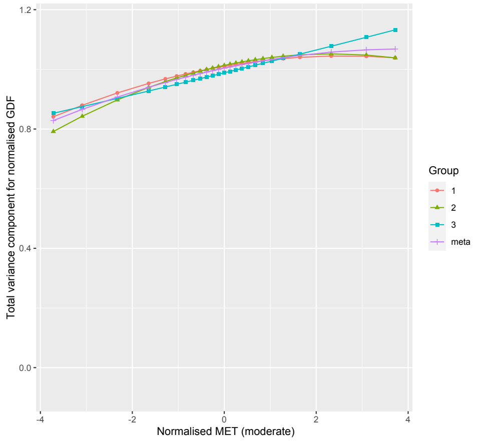

(c) Total variance component

Figure 48: Plots of the variance components for normalised depSympt across normalised MET (moderate)
